# Transition time from manic and mixed episodes to depression: risk factors and outcomes

**DOI:** 10.64898/2026.06.29.26356830

**Authors:** Chloe X Yap, Rachel Upthegrove, Fang Li, Michael Berk, Philip McGuire, Maxime Taquet

## Abstract

**Objective:** For bipolar disorder, recovery from manic or mixed episodes is frequently complicated by depression. This depression may occur within a broader episode pattern of mania-depression-euthymic interval, proposed as a subtype for which lithium is likely effective. However, the window of risk for mania/mixed-to-depression transition remains unclear. This study determines the mania/mixed-to-depression transition rate, and identifies risk factors and outcomes.

**Methods:** This retrospective cohort study includes 100,776 people (502,877 mood episodes; mean age at mood episode 41 years (SD=16), 65% female) with bipolar disorder from the NeuroBlu health record database (United States). Exposures were previous short transition time, hospitalization, clinical severity, and medications prescribed during the manic/mixed episode. The outcome was transition time from manic/mixed episodes to depression.

**Results:** 42% of manic and 29% of mixed episodes transitioned to depression within 1 month: an incidence up to 11-times higher than the overall depression rate.

Short depression transition time (≤1 month) was associated with previous short transition (RR=2.47-3.62, p<1.0e^-100^), higher manic/mixed severity (RR=1.18-1.34, p<2.7×10^-12^) and manic/mixed hospitalization (RR=1.05-1.33, p<0.012).

When prescribed during hospital-associated manic/mixed episodes, lithium (RR=0.87, p=0.0013), and antiepileptic mood stabilizers (RR=0.87-0.89, p<8.3×10^-5^) were associated with longer transition time. Combination lithium/antipsychotic and valproate/antipsychotic were more protective than antipsychotics alone.

Shorter transition time was associated with more depression-related hospital days (8-15% fewer days per month delay to depression, p<3.1×10-5).

**Conclusions:** It is important to monitor for depression soon after manic/mixed episodes. This depression may be predictable, and might be modifiable by some medications prescribed during the manic/mixed episode.

## Introduction

A substantial minority of patients with bipolar disorder rapidly transition to depression following mania. However, estimates vary widely (5-20%(1–3)) and trajectories to depression are poorly characterized. Depression onset during recovery from mania is important as it may be associated with hospitalization(4) and poorer functional recovery.(5)

Depression after a manic/mixed episode may occur within recurrent mood episode patterns. These patterns formed the basis of proposed bipolar subtypes in the 1970-80s, including the “mania-depression-euthymic interval” (MDI) subtype(6) which is one of the strongest predictors of response to lithium maintenance therapy (4-times the odds of response than those with depression-to-mania-euthymic interval pattern).(7–9) However, the transition times between mood episodes that would underpin classification were never defined, so this subtyping was not widely adopted. Instead, current treatment guidelines take a cross-sectional view of bipolar, treating each episode without regard for longitudinal patterns.

Large-scale, robust evidence on mania/mixed-to-depression transition trajectories is needed to inform monitoring and potentially prevention. Electronic health records (EHR) allow characterization of mania/mixed-to-depression transition time using real-world longitudinal data. Here, we perform the largest epidemiological study to date of mania/mixed-to-depression transition time among 100,776 individuals with bipolar disorder within a United States EHR dataset. We hypothesised that 1) depression is common soon after mania/mixed episodes; 2) clinical features and treatment within mania/mixed episodes may affect depression transition time; and 3) transition time relates to depression outcomes.

## Methods

### Data source

In this retrospective multicentre cohort study, we used the NeuroBlu(10) EHR database (version: May 2025) covering 36 million people across the United States with a psychiatric diagnosis seen in both physical and mental healthcare and both inpatient and outpatient clinics from 1999-2025 (**Appendix S1**). Neuroblu has a waiver of authorisation for analysis of deidentified health-care data from the WCG Institutional Review Board (WCG-IRB 1-1470336-1) so ethical approval and individual patient consent were not required.

### Cohort definition

We defined bipolar disorder as a lifetime ICD-9 or ICD-10 diagnosis of ≥1 manic or mixed episode plus ≥1 depressive episode (**Appendix S1**). This study followed STROBE reporting guidelines.

### Defining mania/mixed-to-depression transition time

Manic/mixed and depressive episodes were inferred from serial ICD diagnosis codes (**Appendix S1, Figure S1**) where episode start and end were defined from contiguous diagnosis dates. We quantified the time from manic/mixed episodes to depressive episodes. As bipolar depression is likely underdetected based on ICD codes (**Figure S2**), we undertook further case-finding focused on the 6 months post-manic/mixed episodes to identify new or earlier depression diagnoses using 1) Patient Health Questionnaire-9 (PHQ-9) score ≥10; 2) derived Montgomery-Asberg Depression Rating Scale (MADRS) ≥20 (a variable derived in NeuroBlu which combines MADRS score and a CGI-S proxy); 3) for manic episodes only, antidepressant prescription other than trazodone (frequently prescribed for sleep). **Appendix S1** and **Table S1** provide further detail.

We defined mania/mixed-to-depression transition time as the manic/mixed episode end to the depressive episode start. As the 1 month post-manic/mixed episode corresponded to the highest risk period, this was defined as “short mania/mixed-to-depression transition”. As 6 months corresponded to the excess risk window and to mitigate health-seeking/access biases affecting depression detection, time-to-event analyses of mania/mixed-to-depression transition time were censored to ≤6 months.

### Clinical and medication factors associated with manic/mixed-to-depression transition time

For the clinical factors analysis, we considered: hospitalization for mania/mixed episode, maximum Clinical Global Impression-Severity (CGI-S; within the first 7 days of mania/mixed episode start) and previous short mania/mixed-to-depression transition (≤1 month).

For the medications analysis, we considered psychotropic classes (**Table S2**) commonly prescribed during mania/mixed episodes (antipsychotics, antiepileptic mood stabilizers, lithium, antidepressants, first-generation antihistamines and benzodiazepines) whose prescription coincided with a manic/mixed episode. Here, the manic/mixed episode start had to be within 7 days of the visit start, and the medication had to be started within -7 to +14 days of visit start and before a diagnosis change to depression (**Figure S3**). We modeled prescription as a binary exposure for classes prescribed in ≥200 episodes.

Since people receiving psychotropic medications are significantly more unwell and likely to be hospitalised than those without (**Figure S4**), and illness severity more closely relates to prescription of some medications over others (**Figure S5**), we limited this analysis to hospital-associated episodes to mitigate confounding-by-indication (**Appendix S1**). A sensitivity analysis included all manic/mixed episodes with and without hospitalization.

We performed 3 analyses:

1. For each medication class, we modeled the covariate-adjusted association with mania/mixed-to-depression transition time.
2. Jointly modeling medication classes with covariates to account for co-prescription.
3. For Bonferroni-significant medication classes in the joint analysis, we jointly modeled individual medication effects (each prescribed in ≥200 episodes) as a sensitivity analysis.

We investigated the relative efficacy of mood stabilizers, antipsychotics, and their combination as a sensitivity analysis. We used one model per mood stabilizer (lithium, valproate, lamotrigine), plus one aggregating mood stabilizers. For the lithium/valproate/lamotrigine models, we excluded episodes where other mood stabilizers were prescribed to isolate specific drug effects.

### Depression outcomes associated with mania/mixed-to-depression transition time

For manic/mixed episodes which transitioned to depression within 6 months, we investigated the relationship between transition time and three outcomes in the 6 months following depression onset:

1. Number of depression-related hospital days (defined as inpatient or emergency visits with visit start within 7 days of a depression diagnosis);
2. Number of all-cause hospital days;
3. Maximum PHQ-9 total score.

### Statistical analysis

#### Covariates

We adjusted for age at episode, total years in EHR (from 1999 to 2025), gender, state of residence, race, education, relationship, employment (**Table 1**). For the medication analyses restricted to hospitalization, we adjusted for length of hospital stay and investigated time-to-follow-up as a mediator.

**Table 1:** Summary of demographic factors comparing those with a lifetime mania/mixed-to-depression transition time ≤1 month (MDTT) versus those without.

| **Demographic** | **Detail** | **Lifetime MDTT<1m** | **No lifetime MDTT>1m** |
|---|---|---|---|
| N | Individuals | 24984 | 75792 |
| Age | EHR entry mean (SD) | 40 (16) | 40 (16) |
|  | Mean age of 1<sup>st</sup> recorded mood diagnosis (SD) | 41 (16) | 41 (16) |
|  | Current mean (SD) | 47 (16) | 48 (16) |
| Sex | Female (%) | 16419 (65.7) | 49397 (65.2) |
| Relationship status | Married (%) | 1206 (4.8) | 3134 (4.1) |
|  | Single (%) | 4357 (17.4) | 10654 (14.1) |
|  | Other (%) | 1418 (5.7) | 4063 (5.4) |
|  | Unknown (%) | 18003 (72.1) | 57941 (76.4) |
| Employment status | Employed any (%) | 171 (0.7) | 277 (0.4) |
|  | Not working (%) | 563 (2.3) | 1368 (1.8) |
|  | Unknown (%) | 24250 (97.1) | 74147 (97.8) |
| Educational level (highest) | High school or less (%) | 484 (1.9) | 940 (1.2) |
|  | Tertiary (any) (%) | 293 (1.2) | 645 (0.9) |
|  | Unknown (%) | 24207 (96.9) | 74207 (97.9) |
| Race | American Indian or Alaskan (%) | 18 (0.1) | 61 (0.1) |
|  | Black African American (%) | 3235 (12.9) | 9932 (13.1) |
|  | Pacific Islander (%) | 56 (0.2) | 52 (0.1) |
|  | White (%) | 16811 (67.3) | 48823 (64.4) |
|  | Other (%) | 2529 (10.1) | 6922 (9.1) |
|  | Unknown (%) | 2055 (8.2) | 8925 (11.8) |
| Observation time | years (median [IQR]) | 5.4 [2.99-6.96] | 5.78 [3.63-7.01] |
| Count of episodes | Depression | 103,029 | 192,007 |
|  | Mania | 28,514 | 64,118 |
|  | Mixed | 21,036 | 94,173 |
|  | Depression per-person (median [IQR]) | 3 [2-5] | 2 [1-3] |
|  | Mania per-person (median [IQR]) | 1 [1-2] | 1 [1-2] |
|  | Mixed per-person (median [IQR]) | 1 [1-2] | 1 [1-2] |
| Count of visits | Emergency | 58,059 | 138,832 |
|  | Inpatient | 28,305 | 68,812 |
|  | Outpatient | 2,795,096 | 7,399,631 |
|  | Emergency per-person (median [IQR]) | 6 [2-14] | 6 [3-15] |
|  | Inpatient per-person (median [IQR]) | 3 [2-6] | 3 [1-6] |
|  | Outpatient per-person (median [IQR]) | 62 [27-132] | 53 [25-111] |

Where percentages do not add up to 100%, missing data make up the remaining. Breakdown of participants by state of residence is provided in **Figure S38**.

#### Per-month rate ratio of depression incidence

We calculated the rate ratio of depression for each month after manic/mixed episodes, compared to the cohort overall depression rate. Example calculation: **Appendix S1**.

#### Risk ratio of short mania/mixed-to-depression transition (≤1 month)

We calculated risk ratios, using covariate-adjusted modified Poisson regression(11) i.e., Poisson regression with a robust error variance (R lmtest::coeftest(12)) using clustered covariance matrix estimation to account for repeated within-individual measures. We performed cause-specific hazard analysis (**Appendix S1**) to account for loss to follow-up but consider this a secondary analysis due to violation of proportional hazards assumptions (**Table S3-5**). We explored the impact of adjusting for the mediator of time-to-follow-up, and performed sensitivity analyses for medication continuation versus initiation (**Appendix S1**). We also performed a target trial emulation (**Appendix S1**).

#### Associations between mania/mixed-to-depression transition time and depression severity

Covariate-adjusted generalized estimating equation Poisson regression (R geepack:: geeglm(13)) modeled depression transition time (exposure) against hospital days (outcome), where robust standard errors account for repeated measures and overdispersion.

Covariate-adjusted linear models tested associations between peak PHQ-9 total score and transition time, using robust standard errors for repeated measures.

#### Negative control outcome

We used a composite negative control outcome of upper respiratory tract infections and allergic rhinitis (**Appendix S1**).

#### Multiple testing

We report unadjusted two-sided p-values and denote associations passing Bonferroni correction (**Appendix S1**). Sensitivity analyses did not correct for multiple-testing.

## Results

### Time to depression transition after manic/mixed episodes

The total sample comprised 100,776 individuals with bipolar disorder with 502,877 mood episodes (**Figure S6**). 65% were female with a mean age at first recorded diagnosis of 41 years (SD=16), mean EHR entry age of 40 years (SD=16), and median 4.4 years of observation [IQR: 1.8-6.6] (**Table 1, Table S6**).

67% of 92,632 manic episodes (50,675 people), and 52% of 115,209 mixed episodes (55,477 people) had depression recorded as the next mood episode (**Figure S7**: episode duration distribution). 21% had another manic/mixed episode recorded next, and 20% had no following episode recorded. Mania/mixed-to-depression transition time (**Figure S1**) was usually brief (**Figure S8**): ≤1 month for 42% of all 92,632 manic episodes and 29% of all 115,209 mixed episodes, and ≤6 months for 57% of manic and 41% of mixed episodes.

Within 1 month post-manic episode, the depression episode rate was 10.9-times higher and the post-mixed rate 7.5-times higher than the overall rate of depressive episodes recorded in this cohort’s health records (0.46 depressive episodes per person-year) (**Figure 1b**, **Table S7, Appendix S1**). Depressive episodes within 1 month of mania/mixed episodes accounted for 24% (n=71,911) of all 295,036 depressive episodes.

**Figure 1:**
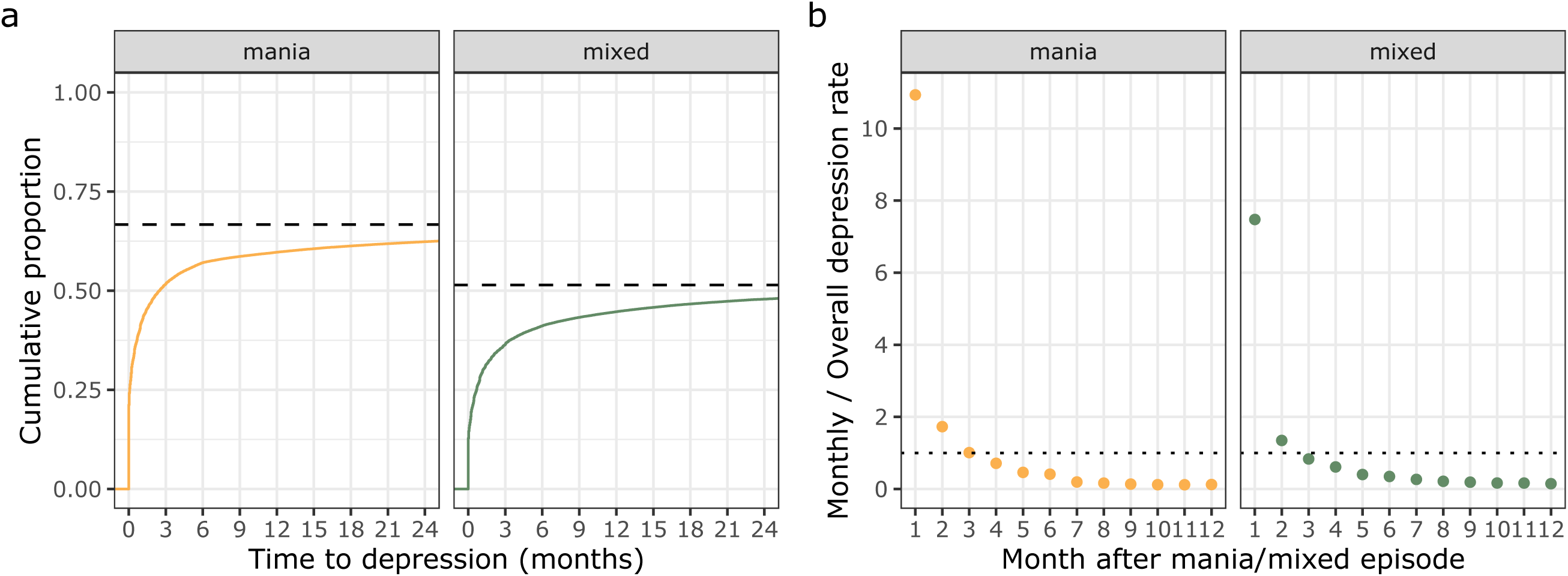
Overview of mania/mixed-to-depression transition time. a) Cumulative proportion of manic/mixed episodes transitioning to depression over time. Dashed line: proportion of all participants who transition to depression from a manic/mixed episode (without another manic/mixed episode). b) Ratio of per-month rate of depression transition, relative to the overall rate of depression in this bipolar sample. Dotted line: rate ratio=1. Data from total 208,825 manic/mixed episodes from 100,776 individuals. Source data: **Table S7**. ***Alt text***: Rates of depression transition are highest in the 1 month after a manic/mixed episode.

We hereafter define this mode of depression onset risk within 1 month post-manic/mixed episode as “short mania/mixed-to-depression transition” (demographics in **Table 1**). Severity peaks around 1.7 months (**Appendix S1, Figure S8**). After around 6 months post-manic/mixed episode, the excess transition risk stabilizes (**Figure 1a, Figure S9**), meets the all-time depression rate (**Figure 1b**), and visit frequency plateaus (**Figure S10**). We note evidence of delayed depression diagnosis: ICD diagnosis lagged self-report depression by ≥28 days in 15% of mania/mixed-to-depression transitions with matched data (**Figure S2**). Negative controls performed as expected (**Figure S11-12**).

### Clinical associations with mania/mixed-to-depression transition time

We investigated clinical factors during the manic/mixed episode associated with depression transition time (**Figure 2**; **Table S8**; **Figure S13**; overall sample: 200,558 manic/mixed episodes from 96,970 people). The risk of short depression transition time (i.e. ≤1 month) was more than doubled in those with previous short transition (post-mania RR=2.47, 95% CI: 2.39-2.56, p<1×10^-100^; post-mixed RR=3.62, 95% CI: 3.48-3.75, p<1×10^-100^) (**Figure S14**). Each 1-point increase in CGI-S within 7 days of mania start was associated with a 18% increase in short depression transitions (95% CI: 1.13-1.24, p=2.7×10^-12^) and 34% increase post-mixed episodes (95% CI: 1.27-1.42, p=8.7×10^-28^). 92% of manic/mixed episodes with CGI-S ≥5 were associated with short depression transition compared to 51% in those with CGI-S <5 (**Figure S14**). Hospitalization was associated with increased risk of shorter transitions (post-mania: RR=1.05, 95% CI: 1.01-1.10, p=1.3×10^-2^; post-mixed: RR=1.33, 95% CI: 1.27-1.40, p=1.1×10^-31^). Sensitivity analyses with no covariates, using cause-specific hazard analysis, stratifying by state and race, and negative controls supported the primary results (**Figure S15-19**).

**Figure 2:**
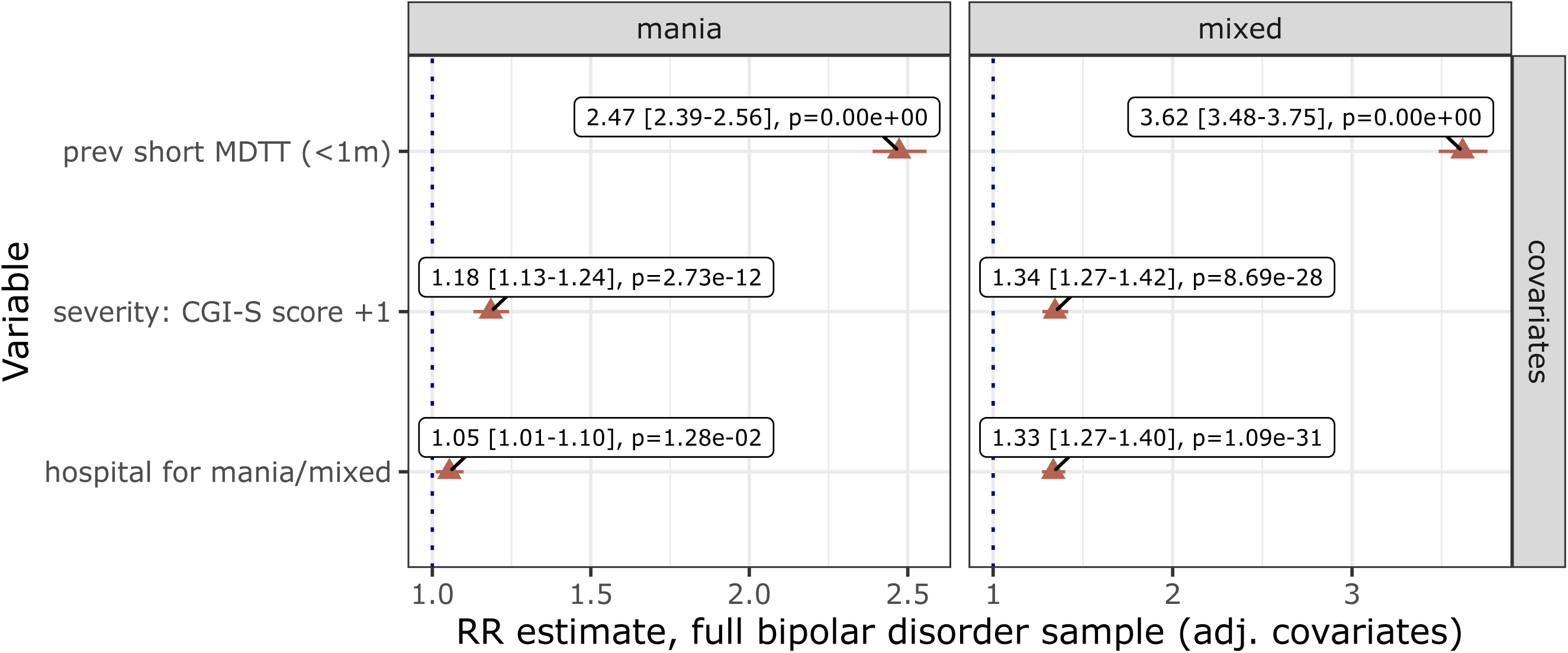
Clinical factors associated with mania/mixed-to-depression transition time within the full bipolar sample. Rate ratio coefficient plot (±95% CIs) for mania/mixed-to-depression transition time using modified Poisson regression, adjusting for covariates and with clustered standard errors to account for repeated measures. Red indicates Bonferroni-significant associations, blue indicates nominal significance (p<0.05). The hospitalization analysis included 104,663 episodes from 51,817 individuals, filtering on states with a ≥1% hospitalization rate; CGI-S analysis included 2,470 episodes from 1,619 individuals; and the previous short mania/mixed-to-depression transition time (MDTT) analysis included 108,389 episodes from 40,082 individuals. Source data: **Table S8**. ***Alt text***: Previous mania/mixed-to-depression transition time and higher severity of mania are associated with faster transition to depression.

### Medications prescribed during manic/mixed episodes associated with hospitalization

In the medication analysis we included 7,313 manic (5,562 individuals) and 5,398 mixed episodes (4,175 individuals) associated with hospitalization, accounting for covariates (**Figure S20-21**) and co-prescription (**Figure S20, 22**). Prescription of lithium (RR=0.87, 95% CI: 0.80-0.95, p=1.3×10^-3^) during mania was associated with delayed depression transition after multiple-testing correction (**Figure 3**; **Table S9**) as were antiepileptic mood stabilizers (RR=0.89, 95% CI: 0.84-0.94, p=8.3×10^-5^), with this class effect driven by valproate (**Figure S23**). Benzodiazepines were associated as a class (RR=0.93, 95% CI: 0.88-0.98, p=5.3×10^-3^) but not supported in the individual medication sensitivity analysis (**Figure S23**). Antipsychotics and antihistamines were only nominally associated after accounting for time-to-follow-up.

**Figure 3:**
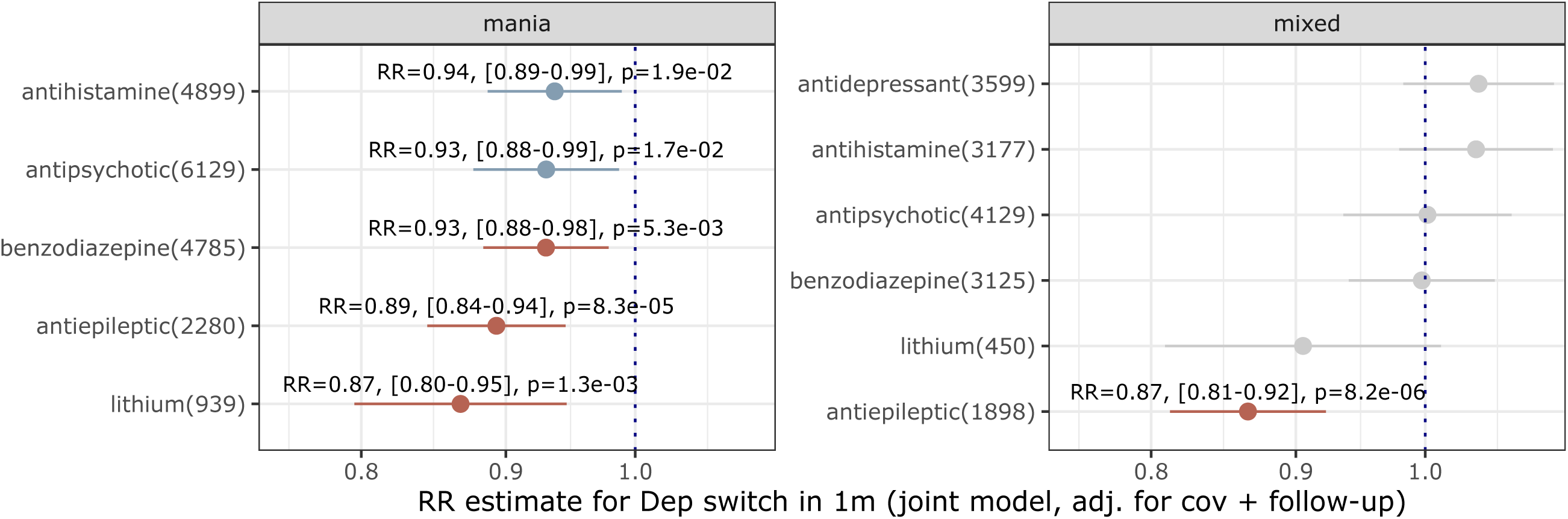
Medication classes prescribed during hospital-associated manic/mixed episodes and their relationship with short mania/mixed-to-depression transition (≤1 month; modified Poisson regression). All displayed medications modeled jointly with covariates including hospital length of stay and adjusting for mediating effects of time to follow-up. Mania analysis includes 7,313 episodes from 5,562 individuals; mixed episode analysis includes 5,398 episodes from 4,175 individuals. Red indicates statistically significant associations after Bonferroni multiple-testing correction; blue indicates nominal significance (p<0.05). Source data: **Table S9**. ***Alt text***: Lithium and valproate prescribed during mania/mixed episodes are associated with lower rates of short depression transition (≤1 month).

During mixed episodes, delayed depression transition after multiple-testing correction was associated with prescription of antiepileptics (RR=0.87, 95% CI: 0.81-0.92, p=8.2×10^-6^) and particularly valproate (**Figure 3**; **Figure S23**). Lithium showed a suggestive association (RR=0.87, 95% CI: 0.74-1.01, p=0.06), noting smaller sample size than other classes. Antipsychotics, antihistamines and benzodiazepines had little evidence of effect.

These results were supported by time-to-event analysis (**Figure S24**; noting that proportional hazard assumptions were violated; **Table S5**) and sensitivity analyses stratifying by state of residence and race; analysing the full sample without restricting to hospitalization; not adjusting for time-to-follow-up as a mediator (**Figure S21**); stratification by continued versus (re-)initiated medications; and negative controls (**Appendix S1, Figure S25-31**).

We investigated mood stabilizer/antipsychotic combinations (**Figure 4**, **Figure S32, Table S10**). Compared to antipsychotics without mood stabilizer, combination lithium/antipsychotic was associated with reduced rate of short post-mania depression transition (post-mania: RR=0.83, 95% CI: 0.71-0.96, p=8.3×10^-3^), as was combination valproate/antipsychotic (post-mania: RR=0.73, 95% CI: 0.65-0.81, p=3.9×10^-9^; post-mixed: RR=0.77, 95% CI: 0.68-0.87, p=2.0×10^-5^). Giving neither antipsychotics nor mood stabilizers was associated with faster mania-to-depression transition (p<0.05). Cause-specific hazard analyses were consistent (**Figure S33, Table S10-11**). Target trial emulation also supported combination lithium/antipsychotic and valproate/antipsychotic for mania being associated with lower depression rates at 1 month than antipsychotic-alone (**Figure S34, Appendix S1**).

**Figure 4:**
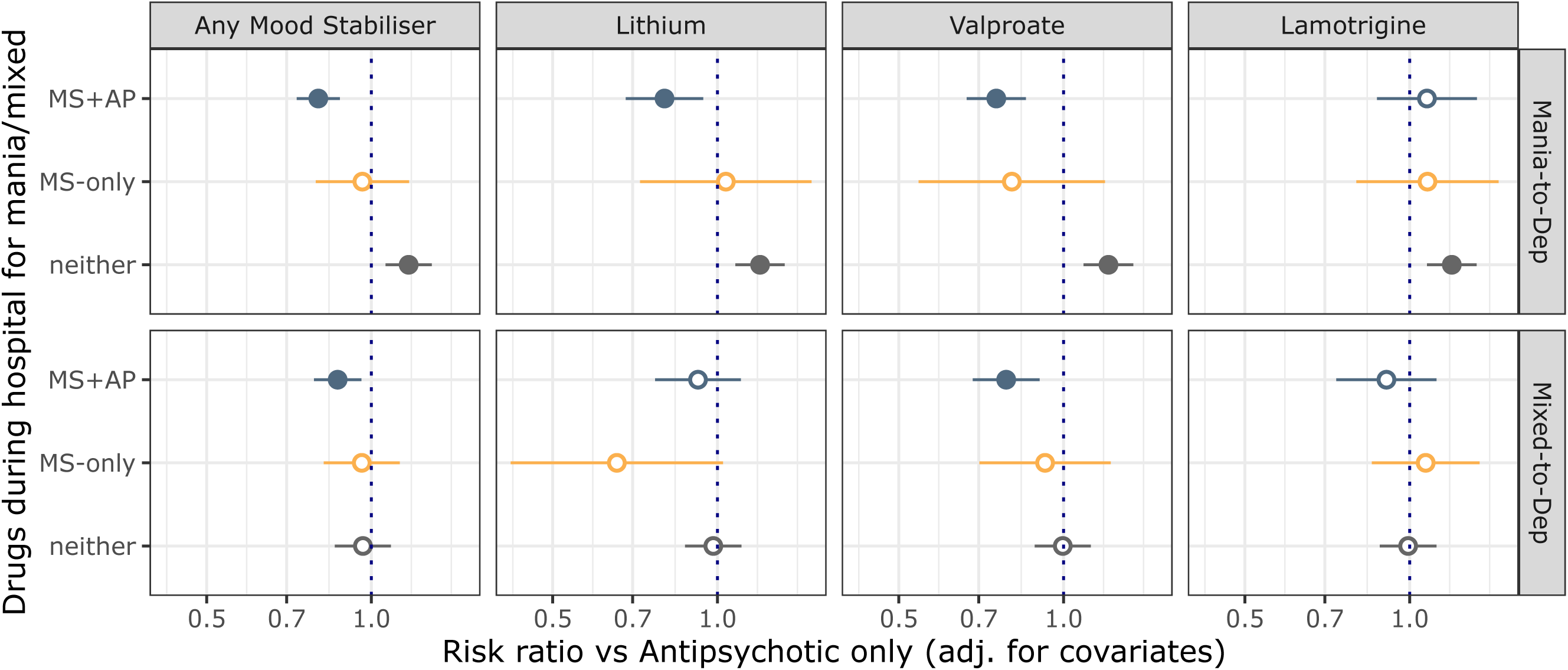
Mood stabilizer and antipsychotic combination effects. Coefficients (±95%CI from clustered standard errors to account for repeated measures) from risk ratio analysis for mood stabilizer/antipsychotic combinations (versus antipsychotic-alone) prescribed during manic/mixed episodes on the risk of short mania/mixed-to-depression transition time after mania/mixed episodes (≤1 month). Mood stabilizers are here considered aggregated as a class (“Any Mood Stabilizer”), then individually (each of lithium, valproate and lamotrigine). Filled points indicate p<0.05. “MS”: mood stabilizer; “AP”: antipsychotic. Source data: **Table S10**. ***Alt text***: Lithium and valproate on top of antipsychotics during mania/mixed episodes are associated with lower rates of short depression transition (≤1 month).

### Depression transition time and depression severity

Shorter mania-to-depression transition was associated with more depression-related hospital days (incidence rate ratio (IRR)=0.85-times, corresponding to 15% fewer days in hospital per month to depression, 95% CI: 0.80-0.89, p=1.2×10^-10^, n=53,728) and all-cause hospital days (IRR=0.91, 95% CI: 0.88-0.94, p=3.7×10^-8^) after multiple-testing. Shorter mixed episode-to-depression transition was also associated with more depression-related (IRR=0.92, 95% CI: 0.87-0.96, p=4.7×10^-5^, n=48,112) and all-cause hospital days (IRR=0.93, 95% CI: 0.90-0.96, p=3.1×10^-5^) after multiple-testing (**Figure S35-36**; **Table S15**). Shorter manic/mixed-to-depression transition was not associated with self-reported depression severity (**Figure S37**).

## Discussion

We explored manic/mixed-to-depression transition time, risk factors and outcomes in a large health record study including 100,776 individuals and 502,877 mood episodes. Depression transition time was usually short. Within 1 month, 42% of manic and 29% of mixed episodes transitioned to depression: a rate 11-times and 8-times higher than baseline, respectively. Short mania/mixed-to-depression transition was associated with severe manic/mixed episodes and previous short depression transition. Mood stabilizers prescribed near the start of manic/mixed episodes were associated with delay or prevention of depression, and combination appeared more effective than antipsychotics without mood stabilizer. Shorter depression transition was associated with more hospital days within 6 months of depression onset.

This work offers an empirically derived window after manic/mixed episodes during which depression risk is high, complementing the few and smaller longitudinal studies investigating recovery after mania.(1–3, 6, 14) We clarify an important gap in the clinically-relevant MDI construct – the time between manic and depressive episodes – which has limited its uptake. Our results highlight consideration of longitudinal episode course and how mania/mixed episodes and depression interrelate(2, 6): medications for acute mania/mixed episodes may affect risk of later depressive switch, and these differ from medications recommended for *acute* bipolar depression (lamotrigine, various antipsychotics and antidepressants(15)).

Lithium during mania/mixed episodes may be protective against depression. This result was robust across sensitivity analyses, including when lithium was (re-)initiated at the time of the manic/mixed episode (**Figure S30**), and even despite small sample size (**Table S2**) and confounding-by-indication towards more severe manic/mixed episodes associated with polypharmacy (**Figure S4-5, 13**) which would bias the effect towards the null. Our results complement the link between MDI and excellent response to lithium as maintenance therapy.(7, 8) Valproate also appeared beneficial, although these results appear more susceptible to bias based on negative control (**Figure S34**) and mediation analyses (**Table S12**). The null antipsychotic results contrast with the common attribution of post-acute depression to antipsychotic side-effects and the EMBLEM study which found associations with lower rates of depression, although prescribers in EMBLEM offered olanzapine to half of patients without randomisation which may induce confounding-by-indication.(3)

We aimed to compare medications in their potential ability to prevent short depression transition. Mood stabilizers were rarely prescribed without antipsychotics, so direct comparison of these two medication classes was not possible. However, combination lithium/antipsychotic and valproate/antipsychotic appeared to prevent short depression transition better than antipsychotics alone. These results were supported by target trial emulation (**Figure S34**), supporting causal inference.

Clinically, short depression transition may be predictable and actionable from the manic/mixed episode. It is at least as common as depression after first-episode psychosis (12-month prevalence of 39%(16) versus 60% post-mania and 45% post-mixed) where careful monitoring for depression is routine and preventative trials are underway(17). Our work supports similar monitoring, identifies high-risk groups (previous short depression transition, severe manic/mixed episodes) and provides preliminary evidence that early mood stabilizers (i.e., during the manic/mixed episode) may prevent depression to confirm in trials.

The high recurrence of short mania/mixed-to-depression transition and the association with lithium response may point to a biological subgroup. The temporal linkage of mania/mixed episodes to depression and that specific medications for mania may prevent depression suggests that depression may sometimes be a natural consequence of mania(6), supporting Kukopulos et al.’s MDI subtype(6) and an intriguing theory that lithium may prevent depression through its action upon mania.(9)

Observational, naturalistic data are susceptible to biases. Record-keeping accuracy may vary across healthcare settings: we limited the medication analysis to hospital episodes for more temporally-precise coding and to prevent circumstances where diagnosis codes could be updated after the treatment initiated for that diagnosis. Visit frequency decays over time which affects depression detection and may reflect prescribing patterns. To mitigate healthcare contact biases, we performed negative control analyses (**Figure S11, 12, 19, 31**), focused on 6-months post-manic/mixed episode, and explored mediating effects of follow-up (**Appendix S1**).

This work has limitations. First, EHR data are based on established constructs (which imperfectly map onto exploratory ones) and are confounded by patterns such as healthcare-seeking behaviours, treatment trends, recording biases, and left-censoring. Individual patient movement between sites would violate assumptions of independence in our models. If true, confidence intervals would be wider, and recurrence rate higher. Second, depression is likely under-detected (**Figure S2**) and the estimated transition times have measurement error dependent on follow-up frequency. Thus, the true peak in depression rate is likely sooner than what our data suggest. Drugs associated with higher appointment frequency (e.g., lithium, **Figure S29**) are more likely to have accurate depression timing, whereas those with less-frequent appointments are likely to have delayed depression diagnosis and appear more protective than they are. Third, we ascertained depression using clinical ICD diagnosis codes plus additional case-finding, but this may introduce demographic biases towards healthcare-seeking behaviours which we mitigated through covariate adjustment. Fourth, potential diagnostic overlap between mixed episodes and depression could inflate the depression transition rate; for example, a mixed episode with predominantly depressive features could transition to major depression when manic symptoms abate. However, we instead found that depression was less likely to be recorded after mixed episodes than mania. Fifth, generalizability: we selected ICD codes specifying polarity, which forms a minority of the total sample with bipolar disorder ICD codes, and may reflect site-related coding practices. Restricting to hospital-associated manic/mixed episodes in the medication analysis also limits generalizability. Sixth, covariate adjustment does not preclude residual confounding. For example, we were able to adjust for state of residence but not site; some sites only capture outpatient data, which may then bias hospitalization effects (**Appendix S1**). There were high rates of missing education, employment and relationship status data.

Replication is needed, ideally including jurisdictions with less pronounced confounding-by-indication. Further work should explore when medications need to be prescribed relative to manic/mixed episode start to modify depression transition time. Prospective studies are needed to closely characterize clinical features of people who go on to develop short mania/mixed-to-depression transition and a MDI pattern, as are studies of longer-term natural history and outcomes. Trials are needed to determine medication effects.

In conclusion, short mania/mixed-to-depression transition deserves closer study: as a common, morbid and under-recognised clinical entity which appears rooted in mania, is potentially prevented by lithium and valproate, and is recurrent, so may proxy a biological subtype of bipolar disorder with overt treatment implications.

## Data Availability

Data analysed in this study are available via application to NeuroBlu with paid access (https://www.neuroblu.ai/).

## Statements

### Author contributions

CXY conceptualised the project, performed all analyses and wrote the first draft. FL also performed analysis. CXY and MT directly accessed and verified the data. MT supervised the project. CXY, MT, RU, MB and PM critically reviewed the manuscript. All authors had final responsibility for the decision to submit for publication. CXY, FL and MT had full access to all the data in the study and take responsibility for the integrity of the data and the accuracy of the data analysis.

## Acknowledgements

CY and RU are funded by the National Institute of Health and Care Research (NIHR) Oxford Health Biomedical Research Centre. MB is funded by the National Health and Medical Research Council (NHMRC) Leadership 3 Investigator grant (GNT2017131). MT is funded by the NIHR. The views expressed are those of the authors and not necessarily those of the UK National Health Service, the NIHR, or the UK Department of Health and Social Care.

## Disclaimer

The views expressed are those of the authors and not necessarily those of the National Institute of Health and Care Research, UK National Health Service, or the UK Department of Health and Social Care.

## Data sharing

Code is available at GitHub https://github.com/cyap7/mdtt_neuroblu

