## Supplementary Material for "Transition time from manic and mixed episodes to depression: risk factors and outcomes"

**Contents**

**Appendix S1**

**Figure S1**: Schematic illustrating collapse of time-stamped diagnoses into episodes.

**Figure S2:** Difference in mania/mixed-to-depression transition time detection between ICD diagnosis codes and broader case-finding criteria.

**Figure S3**: Definition of hospital-associated manic/mixed episodes and relation to prescription.

**Figure S4**: Relationship between medication prescription and hospitalisation.

**Figure S5**: Distribution of CGI-S scores within 7 days of the manic/mixed episode start date in relation to medications prescribed contemporaneously.

**Figure S6**: Number of manic/mixed and depressive episodes per individual recorded in this dataset.

**Figure S7**: Durations (months) of each of the phases of mania/mixed-to-depression transition.

**Figure S8**: Density plots for peak incidence and severity of depression after mania/mixed episodes.

**Figure S9**: Plot of gradient (beta coefficient) of time to depression regressed against cumulative proportion of individuals as plotted in **Figure 1a**.

**Figure S10**: Density plot of the temporal distribution of visits following manic/mixed episodes.

**Figure S11**: Cumulative proportional incidence of negative controls after manic/mixed episodes.

**Figure S12**: Ratio of per-month rate of negative control diagnoses, relative to the overall rate of diagnoses in this bipolar sample.

**Figure S13**: Relationships between prescribing and demographic factors and prescription of major psychotropic medication classes for manic/mixed episodes, from all episodes in this sample of individuals with bipolar disorder.

**Figure S14**: Cumulative density of mania/mixed-to-depression transition time by clinical factors.

**Figure S15**: Clinical factors associated with short mania/mixed-to-depression transition time.

**Figure S16**: Coefficient plot for the effect of clinical factors available at manic/mixed episode diagnosis on the cause-specific hazard ratio for mania/mixed-to-depression transition time.

**Figure S17**: Clinical factors sensitivity analysis stratifying by state of residence.

**Figure S18**: Clinical factors sensitivity analysis stratifying by recorded racial group.

**Figure S19**: Coefficient plot for the effect of clinical factors available at manic/mixed episode diagnosis on the risk ratio of diagnosis of negative controls within 1 month.

**Figure S20**: Relationships between prescribing and demographic factors and prescription of major psychotropic medication classes for hospital-associated manic/mixed episodes.

**Figure S21**: Sensitivity analysis for rate ratio for medication classes prescribed during hospital-associated manic/mixed episodes and relationship with short mania/mixed-to-depression transition.

**Figure S22**: Upset plot for medications sets represented in the medication analysis.

**Figure S23**: Joint analysis of individual medications belonging to the medication classes significantly associated with mania/mixed-to-depression transition time, restricting to manic/mixed episodes associated with hospitalisation.

**Figure S24**: Cause-specific hazard ratio (±95% CI) from Cox regression with repeated risk intervals in medication class analyses restricting to manic/mixed episodes associated with hospitalisation.

**Figure S25**: Medication sensitivity analysis stratifying by state of residence.

**Figure S26**: Medication sensitivity analysis stratifying by recorded racial group.

**Figure S27**: Sensitivity analysis for medications classes prescribed during any manic/mixed episode (i.e., no longer restricting to hospitalisation).

**Figure S28**: Directed acyclic graphs (DAGs) for factors influencing the relationship between prescription during manic/mixed episodes (exposure) and depression transition time (outcome).

**Figure S29**: Boxplot for the distribution of (a) days from hospital visit end to first follow-up and (b) total number of visits within 180 days from hospital visit end.

**Figure S30**: Sensitivity analysis exploring recency of last prescription on the medication results.

**Figure S31**: Medications classes prescribed during hospital-associated manic/mixed episodes and their relationship with negative control outcomes.

**Figure S32:** Number of people per category for mood stabiliser/antipsychotic combination analysis.

**Figure S33:** Mood stabiliser and antipsychotic combination effects on mania/mixed-to-depression transition from cause-specific hazard analysis.

**Figure S34**: Target trial emulation results.

**Figure S35**: Scatterplot of depression-related hospital days and depression transition time.

**Figure S36**: Scatterplot of all-cause hospital days and mania/mixed-to-depression transition time.

**Figure S37**: Scatterplot of time to depression and maximum PHQ-9 score.

**Figure S38**: Percentage of individuals with medication and clinical scale data availability per-state.

### Appendix S1

**Detail on the Neuroblu dataset**

The release code for this dataset is 25R2, created 16 April 2025. It is a real-world de-identified dataset aggregated from EHRs specifically tuned for neuropsychiatric research. The data in NeuroBlu have been collected during provision of routine healthcare across multiple specialty institutions (n=32) in the US throughout the years 1999-2025 (latest follow-up is 15 June 2025). Patients were included into the dataset if they were diagnosed with at least one mental or behavioral disorder. For the identified patients, all corresponding records available in the EHR system are collected and de-identified.

The provider types differ in how much of the care they record is neurological and psychiatric care (behavioral health broadly). Of the diagnosis records coming from the specialty psychiatric network, 75.7% are mental or behavioural health diagnoses. The same measure is 25.4% at the urban public and safety-net systems, 15.9% at the large ambulatory network, and 12.2% at the multi-state integrated delivery networks. Each percentage describes only that provider’s own records.

Number and type of contributing sites

The dataset includes data from 32 provider organisations:

- A national ambulatory EHR network (30.9M patients with an encounter). Community-based outpatient practices across the US. The care delivered here is general ambulatory medicine, delivered to a behavioural health cohort.
- Two multi-state integrated delivery networks (3.8M). Settings include general acute care hospitals, ambulatory medical specialty and primary care, emergency departments, pharmacy, ambulatory surgery, home health, hospice and nursing facilities. These contribute the inpatient and emergency record.
- Six urban public and safety-net systems (1.1M). County mental health authorities and academic safety-net providers. Settings are distinctively public-sector behavioural health: community mental health centres, psychiatric hospitals, inpatient psychiatric facilities, school-based services, early intervention agencies, residential substance-abuse treatment, homeless shelters and outpatient rehabilitation.
- Twenty-five sites of a legacy specialty psychiatric research network (539K). Almost entirely community mental health centres. This is the specialty behavioural-health core and the source of the deepest historical record, back to 1992.

Across all contributors, 66 distinct place-of-service types are represented, including 997 general acute care hospitals, 1,825 ambulatory specialty clinics, 398 ambulatory primary care clinics, 171 emergency departments, 24 community mental health centres, 8 inpatient psychiatric facilities and 3 psychiatric hospitals.

Individual contributing organisations are not identified by name. Holmusk does not disclose data-provider identities in publications or external documentation, and describes contributors by type and care setting instead.

Distribution of data across sites and states

The ambulatory network accounts for 84.9% of patients; the other 31 organisations share the remaining 15%.

State of residence is recorded for 34,784,547 patients (95.9%) and spans all 50 states, DC, Puerto Rico and Guam. The states with the most patients are California (3.58M), Texas (2.58M), Florida (2.53M), Ohio (2.40M) and New York (1.62M).

Note that state reflects patient residence rather than site location; for the national ambulatory network the two diverge, since its patients span every state from a single mapped care-site record

Completeness of medication capture (psychiatric medications)

NeuroBlu data captures all medication records entered at the participating sites. These records reflect prescriptions as documented in the EHR; consequently, they indicate that a medication was ordered but not that the prescription was filled at a pharmacy, nor that it was taken by the patient. The exception is medication administered directly on the provider’s premises, for example long-acting injectable antipsychotics, for which administration can be confirmed. Because the participating institutions predominantly deliver psychiatric care, medication capture is most complete for psychiatric agents. Prescriptions for non-psychiatric conditions, for example chronic diseases typically managed in primary care by clinicians outside the psychiatric team, are expected to be under-reported, as they may be prescribed and recorded elsewhere. The degree of this under-reporting cannot be established with confidence and should be considered when interpreting non-psychiatric medication data.

Specifically related to the bipolar disorder cohort, 25R2 holds 1,985,502 patients carrying at least one bipolar disorder diagnosis, of which 1,700,992 (85.7%) have at least one psychiatric medication record.

Completeness of outcome capture

NeuroBlu data includes structured clinical assessments (for example, the PHQ-9) where these were administered and recorded. However, measurement-based care is not applied consistently across all settings, and in routine practice structured assessments are not administered on a standardized cadence or in a standardized volume. As a result, the number and frequency of recorded assessments vary between patients. Two patients with the same diagnosis (for example, major depressive disorder) may have different numbers of recorded measurements, and some may have none at all.

Because NeuroBlu data reflects data generated during routine care, coverage and completeness track real-world clinical documentation practices rather than a prospective protocol. Given these properties, it is valuable for studying care as it is actually delivered, while also meaning that completeness varies by data type and setting.

Specifically related to the bipolar disorder cohort, the following coverage is observed:

- ICD diagnoses: 1,985,502 patients (100%), 32,907,433 records, through June 2025.
- PHQ-9 total score: 499,094 patients (25.1%), 1,998,723 records, 1996 to 2025.
- CGI-S: 69,534 patients (3.5%), 2,972,122 records, 1992 to 2021.
- Inpatient encounter: 180,262 patients (9.1%), 663,171 records, through June 2025.

Note that in analysis, we analysed data from a subset of the bipolar disorder cohort with ICD codes specifying episode polarity (see section: **ICD codes**), reducing the cohort to 100,776 patients (see section: **Cohort definition – further detail**).

**ICD codes**

- Mania/hypomania codes were from ICD-9 (296, 296.0, 296.00, 296.01, 296.02, 296.03, 296.04, 296.05, 296.4, 296.40, 296.41, 296.42, 296.43, 296.44, 295.45) and ICD-10 (F30, F30.0, F30.1, F30.10, F30.11, F30.12, F30.13, F30.2, F30.8, F31.0, F31.0, F31.10, F31.1, F31.10, F31.11, F31.12, F31.13, F31.2, F31.71, F31.73).
- Mixed codes were from ICD-9 (296.6, 296.60, 296.61, 296.62, 296.63, 296.64, 296.65) and ICD-10 (F30.6, F31.60, F31.61, F31.62, F31.63, F31.64, F31.77).
- Depression codes (unipolar or bipolar) were from ICD-9 (296.50, 296.51, 296.52, 296.53, 296.54, 296.55, 296.20, 296.21, 296.22, 296.23, 296.24, 296.25, 296.30, 296.31, 296.32, 296.33, 296.34, 296.35) and ICD-10 (F31.30, F31.31, F31.32, F31.4, F31.5, F31.75, F32.0, F32.1, F32.2, F32.3, F32.4, F32.8, F32.9, F33.0, F33.1, F33.2, F33.3, F33.4, F33.8, F33.9).

**Cohort definition – further detail**

As mood episode polarity switching was key to our analysis, we filtered for individuals with diagnosis codes with both poles (mania/mixed and depression), leaving a cohort of 100,776. Notably, of the 1,985,502 people with bipolar disorder in Neuroblu, 835,363 were assigned ICD codes without polarity specifiers; 711,899 patients had depression polarity diagnosis codes without mania/mixed codes; 189,367 patients had mania/mixed polarity codes but no depression specifier codes.

As additional quality control filters, we excluded 474 individuals with >50 mood episodes recorded in the health record as outliers, noting that for these individuals, apparent switches in polarity occurred every couple of days.

**Inferring mood episodes from ICD codes**

Consecutive ICD codes were collapsed into episodes (manic/mixed versus depressive) where they occurred within 90 days of each other. In cases where a manic/mixed/hypomanic and depressive diagnosis episode had coinciding dates, the order was determined by minimising the overall number of switches between manic/mixed/hypomanic states and depressive states throughout that individual’s EHR. Where there were no nearby (<90 days) preceding or subsequent mood diagnosis codes to “anchor” the order of the switch, these episodes were excluded due to it being impossible to infer episode sequence, leading to the exclusion of 14,095 episodes. We also excluded 4,351 manic/mixed episodes with duration >365 days as outliers. This left 755,568 mood episodes from n= 133,811 individuals with evidence of both manic/mixed and depressive episodes.

**Definition of depression following manic/mixed episodes including broad case-finding**

We elected for a broad definition of depression following manic/mixed episodes because depression diagnosis codes tended to lag behind other proxies of depression which indicates under-diagnosis (**Figure S1**). Notably, this was also true for antidepressant prescribing which indicates that this lag includes at least a degree of under-recording (rather than under-diagnosis). We did not perform similar case detection for more distant depression episodes as we assume that they would be correctly coded where the presenting complaint was depression.

Broadening the case-finding definition beyond ICD codes identified an additional 17,572 instances of mania/mixed-to-depression transition ≤6 months, and also resulted in an earlier start date for depressive episodes following 12,442 manic/mixed episodes (**Table S1**).

After these filters, we were left with 524,351 mood episodes from 100,776 patients.

As we included antidepressant prescription start date as a proxy for depression diagnosis, we determined whether this definition was specific to depression and not just capturing episodes of anxiety. Of the depression episodes identified (17,572) or with start date brought earlier (12,442 episodes) using broad case-finding, only a small minority (2.3%) had a diagnosis of anxiety occurring in the previous 1 year (**Table S1**) – a rate that was very similar to depression as defined by ICD codes and broad-case finding based on clinical scales (**Table S1**). We therefore conclude that our antidepressant prescription definition captures depression rather than anxiety.

**Overlap of medication and clinical scale data with the identified bipolar cohort**

Within this bipolar sample, n=96,366 had medication data (95.6%), n= 49,545 individuals had a PHQ-9 score (49.2%), n= 2,432 individuals had at least one CGI-S score (2.4%), and n=2,431 had a derived MADRS score which is derived from CGI-S (2.4%). The percentage of individuals with availability of each data type per state is shown in **Figure S38**. This indicates that medication data is generally complete, but that conclusions drawn using some of the clinical scales (e.g., CGI-S) may be less generalisable than others (e.g, PHQ-9).

**Example calculation of per-month rate ratio of depression**

The overall equation is:

$$Incidence rate ratio= postMania incidence / overall incidence$$

$$=\left( \frac{\#episodes_{Mania} transitioned to Dep}{total observation time postMania} \right)/\left( \frac{\#episodes_{Dep}total}{total observation time} \right)$$

We first estimated an ***incidence rate for depression after manic/mixed episodes***.

To do this, we estimated the conditional probability P(depression in the following 1 month window|manic or mixed episode).

Here we provide an example calculation for the rate ratio of depression within 1 month of mania.

The denominator of the post-mania depression incidence rate on a per-month basis is the summed 1 month follow-up over all manic/mixed episodes. We note that the detection of depression depends on individuals receiving follow-up; further, follow-up becomes less likely over time as patients are discharged; thus, the depression incidence rate calculation should be confined to episodes receiving ongoing follow-up. We therefore exclude from the denominator episodes belonging to individuals who do not receive any follow-up within that 1-month bin:

92,271 manic episodes followed up for 1-month ~ 92,271 post-episode person-months

In person-year units:

92,271 person-months / 12 months per year = 7689.25 person-years

For the numerator of this incidence rate:

38,854 manic episodes transitioned to depression within 0 to 1 months.

Thus the depression incidence rate post-mania is:

38,854 episodes / 7689.25 person-years = 5.05 episodes/person-year.

We repeated these calculations for discrete bins of depression transition time (i.e., 0-1 months, 1-2 months, etc.), and updated the denominator within each bin to exclude episodes lost to follow-up.

To estimate the incidence rate ratio, we compared the mania-to-depression rate to the ***overall depression rate*** (per person-year) captured within the health record.

The overall depression rate is calculated as follows:

The denominator is the total summed years of observation time for individuals who experience at least 1 manic/mixed episode and at least 1 depressive episode. This equals 638,373 years.

The numerator is the total number of depressive episodes (295,036).

Thus the overall depression rate for those who experience at least 1 episode of mania is 295,036/638,373 = 0.46 episodes/person-year.

The incidence rate ratio in the 1 month after mania equals the incidence rate for depression after manic episodes (5.05) divided by the overall depression rate (0.45): an incidence rate ratio of 10.9.

Using the same process to estimate the incidence rate ratio in the 1 month after mixed episodes, gave 7.5 (3.46/0.46).

The result is then interpretable as: *for those with active health records, what is the rate of depression diagnosis per-month after manic/mixed episodes, relative to the rate of episodes detected within a healthcare setting*?

**Time to peak severity of depression after manic/mixed episodes**

We identified n=14,925 manic/mixed episodes followed by a PHQ-9 measurement within 6 months and at least 1 additional (total ≥2) PHQ-9 measurement within 12 months, then calculated the time to the maximum PHQ-9 total score over a threshold ≥10 (moderate severity depression). We took the date corresponding to the highest PHQ-9 score as the peak severity for that episode, and took the median time to peak severity across all episodes. Where there were tied peak PHQ-9 scores, we randomly selected one mania/mixed-depression transition time.

We determined how the time to peak severity compared to chance by generating an empirical null distribution randomly shuffling for each episode the relationship between PHQ-9 scores and mania/mixed-depression transition times for 1000 permutations. We took as the p-value the rank of the observed result relative to the empirical distribution. Statistical significance was set to p<0.05 in a two-sided test.

We found that depression severity peaked sooner than chance for both manic (median=1.8 months, permutation p<0.001) and mixed episodes (median=1.6 months, permutation p<0.001) (**Figure S8**).

**Confounders and mediators of medication prescription and depression transition time**

Directed acyclic graph (DAG) to guide identification of confounders and mediators

We constructed (using Daggity^1^) a directed acyclic graph (DAG) to depict our causal model and assist in designing our analysis (**Figure S28**). We were particularly interested in the circumstances in which earlier follow-up after hospital discharge could confound or mediate the relationship between prescription choices made during manic/mixed episodes (exposure; **Figure S29**) and depression transition time (outcome). An example of this is prescription of lithium which we show in our data to be associated with earlier follow-up (**Figure S26**) which is likely related to therapeutic monitoring and also initial illness severity (**Figure S4-5**), and is more likely to be co-prescribed with other drugs (**Figure S20**). The interactive DAG is available at: <https://dagitty.net/mNVYSb9h9>.

Under the assumptions used to construct **Figure S28**, adjusting for the following 3 confounders are sufficient to ensure that confounding-by-indication (that is, confounding of the relationship between prescription of a given drug (“drug X”) during a hospital-associated manic/mixed episode and time to depression transition) is controlled for:

- Hospitalisation; or when the dataset is stratified by hospitalisation, length of hospital stay;
- Medication co-prescription; and
- Demographics.

DAGs are helpful as they show explicitly the assumptions underlying confounder selection. The following model modifications may lead to biased estimates of the effect of our exposure on outcome of interest:

- If severity of hospital-associated manic/mixed episodes directly causes earlier follow-up independent of duration of hospital stay.
- If illicit substance use directly causes earlier follow-up / rapid depression switch, independent of duration of hospital stay.
- If the data we have used to adjust for confounders are systematically biased in some way. Duration of hospital stay is likely to be accurate. Drug Y co-prescription during manic/mixed episodes is also likely to be accurately recorded as we restricted to hospital-associated episodes. Demographic data may not be missing at random, so this remains a potential source of residual confounding.

Validating confounders

We first aimed to confirm confounding-by-indication in the full bipolar dataset. For this, we used modified Poisson regression^2^ to test for associations between hospitalisation and medication class prescription during mania/mixed episodes (**Figure S4**). Based on these results, we restricted medication-based analyses to manic/mixed episodes associated with hospitalisation, and also because health records relating to hospital are likely to be more accurately coded.

We also confirmed that prescription is associated with manic/mixed severity (**Figure S5**). Severity data is only available for a proportion of hospital-associated episodes, but hospital length of stay (which proxies manic/mixed severity) is available for all episodes. Thus, we adjusted for hospital length of stay as a confounder in the medication analyses.

Investigating mediators

From **Figure S28**, earlier follow-up is a potential mediator of the relationship between medication prescription and depression transition time among manic/mixed episodes associated with hospitalisation. In particular, lithium is associated with sooner follow-up after the hospital visit.

***We compared the following two analyses***:

1. An analysis of the *overall* relationship between prescription of each broad drug class during manic/mixed episodes 🡪 rapid depression transition (**Figure 3, Figure S21c** repeated).
2. A sensitivity analysis adjusting for the mediating effect of time to first follow-up appointment after the hospital visit (**Figure S21d**) on the medication prescription 🡪 depression transition time relationship.

The discrepancy in results between **Figure S21c-d** indicate that the results are in part explainable by time to follow-up after the hospital visit.

***We also performed a formal mediation analysis***. The limitation of this approach is that we were unable to use the clustered standard errors approach that is required for modified Poisson regression and to account for repeated measures. We therefore simplified the input models to the following, acknowledging that confidence intervals will appear narrower than they truly are:

- Effect of medication 🡪 short depression transition while accounting for log-transformed time until follow-up: logistic regression (previously used modified Poisson regression)
- Effect of medication 🡪 log-transformed time until follow-up: linear regression

Mediation analysis results are tabulated in **Table S12**. Notably, there was evidence of substantial mediation effects, indicating that time to follow-up after discharge meaningfully affects the relationship between medication prescription for manic/mixed episodes and short depression transition time. Antipsychotics (>55%) and antiepileptics (37-71%) were particularly strongly affected. Lithium was the least affected (26-31%). We therefore decided to adjust for time to follow-up in the primary analysis but also retain plots without accounting for time to follow-up for comparison (**Figure S21**).

Note that these analyses assume that earlier follow-up is due to drug prescription and monitoring rather than being a direct consequence of rapid depression switch; otherwise, adjusting for time to follow-up would induce a collider bias. Our model is supported by evidence for differences in follow-up between medications in expected patterns (**Figure S29**; lithium associated with shorter time to follow-up) and also evidence that bipolar depression is underdiagnosed in this cohort (**Figure S2**).

Interpreting medication effects in the context of differential follow-up and depression detection

It is unlikely that differential follow-up is “masking” a beneficial effect of medications other than lithium:

- Lithium was associated with earlier follow-up than other medication classes. This would bias towards earlier detection of depression and artificially increase the rate ratio for rapid depression transition (i.e., a false negative for a “protective” effect). This explains why adjusting for time to first follow-up accentuates the rate ratio versus when not adjusting (**Figure 3, Figure S21c-d**).
- Other medications are associated with a longer time from hospital discharge to the first follow-up appointment. Assuming that patients wait until their scheduled follow-up appointment to seek help for depressive symptoms, this would create a bias towards later detection of depression and artificially reduces the rate ratio for rapid depression transition (i.e., these may be false positives for a “protective” effect).
  - Thus, it is safe to rule out a “protective” effect for medications lacking association with rapid depression transition. If these medication effects incorporate a bias towards later detection of depression, the rate ratio would be still higher.
  - Medications associated with longer time to follow-up and which demonstrate a “protective” signal (i.e,. benzodiazepines, antihistamines) could potentially be false positives, which is supported by the mediation analysis (**Figure 3**, **Figure S21c-d**).

**Cause-specific hazard analysis**

We used multivariable, covariate-adjusted, cause-specific hazard analysis of recurrent risk intervals (cause: depression), accounting for repeated observations within individuals (R package survival::cluster()^3^) and including covariates (R package tidycmprsk::coxph^4^; competing event: mania/mixed recurrence). We censored to a 6-month post-mania/mixed episode window as this was the period within which the depression transition rate plateaued (**Figure 1**, **Figure S9**) as did the visit frequency (**Figure S10**). We checked proportional hazards assumptions using the Schoenfeld residual approach (**eTables 3-5**).

For medications, we used joint models to account for co-prescription and stratified (R package survival::strata()^3^) by 1) hospitalisation for the manic/mixed episode and 2) history of previous mania/mixed-to-depression transition ≤1 month, as this better satisfied proportional hazards assumptions, tested using the Schoenfeld residual approach.

We overall found widespread violation of proportional hazard assumptions including when using stratification approaches. Due to challenges with interpretation, we present risk ratios as the primary analysis.

**Target trial emulation**

A TTE framework supports comparisons between treatments. This framework mitigates confounding-by-indication by using inverse probability of treatment weighting to balance groups by measured baseline covariates, and with clearly defined time-zero to avoid immortal time bias.

We designed a TTE to answer the following two questions:

- Is lithium + antipsychotic therapy superior to antipsychotic alone?
- Is valproate + antipsychotic therapy superior to antipsychotic alone?

We compared combination therapy to antipsychotic-alone, as there were insufficient numbers of individuals with either lithium or valproate without antipsychotic prescription. Thus, this analysis is most analogous to the mood stabiliser/antipsychotic analysis whose results are depicted in **Figure 4**.

Design

- Population:
  - Inclusion criteria:
    - The first manic episode within the period of 2010-2025 (i.e., 1 episode per person).
    - Hospitalisation for the manic episode.
  - Exclusion criteria:
    - Hospital length of stay >30 days (as there is the potential for the depressive episode to have started within this window).
    - Severe renal disease
    - Severe hepatic disease
    - Pregnancy
    - Patients were excluded if lithium or valproate had been prescribed at any point in the 180 days before admission, thereby reflecting medication (re-)initiation.
- Exposure: Groups were assigned based on what medications were prescribed during the hospitalisation stay for the manic (only manic) episode.
  - Antipsychotics + lithium: n=1,191 vs antipsychotics-alone (specifically no valproate): n=3,437
  - Antipsychotics + valproate: n=2,469 vs antipsychotics-alone (specifically no lithium): n=3,437
- Covariates to balance by – all data available at the start of hospitalisation for the manic episode:
  - Age at index
  - Gender
  - Race
  - Index year
  - Region
  - Education
  - Relationship status
  - Employment status
  - The number of years of recorded history
  - Diagnosis of schizophrenia
  - Diagnosis of schizoaffective disorder
  - Diagnosis of any other psychotic disorder
- Baseline definition: Start of hospitalisation for mania. Eligibility, treatment group and all covariates are determined as of this point.
- Interval between baseline and time zero: the hospital stay itself. Treatment group is decided by what is prescribed during the stay, and patients in whom a depressive episode is recorded before discharge are excluded, so this interval contributes neither events nor person-time to the analysis.
- Start of follow-up (time zero): The end of hospitalisation
- Follow-up period: from discharge until 30 days after
- Outcome: Depression diagnosis within 30 days of the end of hospitalisation, as defined by ICD diagnoses, PHQ-9 ≥10 and derived MADRS ≥20
- Negative controls: allergic rhinitis and upper respiratory tract infection

Analysis:

Baseline covariates for antipsychotics alone, antipsychotics plus lithium, and antipsychotics plus valproate are presented as means and standard deviations or frequencies and proportions, as appropriate. We used stabilised inverse probability of treatment weighting to construct a pseudo-population in which the measured baseline covariates were independent of the assigned treatment strategy. Balance was assessed using standardised mean differences between treatment strategies before and after weighting, with an absolute standardised mean difference below 0.1 considered adequate (**Table S13**).

In this pseudo-population, we estimated, for each treatment strategy, the risk of a depression diagnosis by day *t* under the counterfactual scenario in which all eligible patients followed that strategy from discharge onward. For each arm, the risk was obtained from a weighted Kaplan–Meier curve and calculated as one minus the survival probability. The effect measure at 30 days was the risk ratio, with antipsychotic monotherapy without lithium or valproate as the reference. This estimator does not require the proportional hazards assumption. Confidence intervals and two-sided P values were obtained from 2,000 bootstrap resamples of the cohort, with the weights re-estimated in each resample.

Distinction between the primary analysis (marginal effects) and the current TTE

- Goal: Our primary analysis indicates the effect of 1 medication class on the outcome, while holding all other variables constant, which allows us to test a broader number of medication classes in a single model. In contrast, the TTE provides evidence for whether one intervention is superior in comparison to another, by balancing covariates between exposure groups based on baseline data.
- Inclusion/exclusion criteria: The primary analysis did not have inclusion and exclusion criteria as that analysis sought to understand the marginal effects of each medication. The TTE framework has inclusion and exclusion criteria, which are used to ensure that the groups are exchangeable (i.e., both groups have to be equally able to be prescribed lithium vs lithium + antipsychotic).
- Accounting for severity of the manic episode: Severity ratings at the start of the manic episode have high rates of missing data which cannot be adjusted for. Length of hospital stay has complete data and so was adjusted for in the primary analysis. However, the TTE framework strictly requires covariate data to be available at baseline (i.e., start of hospitalisation) to simulate “randomisation”. We therefore performed a post-hoc analysis comparing length of hospital stay between the arms to explore the relationship between length of hospital stay and prescribing.
- Accounting for co-prescribed medications: The primary analysis accounted for other medications that are prescribed to treat mania within +/-7 days of hospitalisation. In the TTE framework, it was not possible to account for medications prescribed before the baseline timepoint (i.e., hospitalisation). Note that polypharmacy is highest in association with lithium, followed by antiepileptics, whereas antipsychotics are least likely to be prescribed alongside other psychotropic medications (**Figure S20**).
- Interpretation as continuation versus (re-)initiation: The primary analysis binarised the exposure based on whether there was any prescription of a given medication within +/-7 days of the start of hospitalisation. This was followed by a sensitivity analysis stratifying the exposure by evidence of medication continuation versus a (re-)initiation (**Figure S30**). In contrast, the TTE only considered cases of (re-)initiation as this gives a cleaner design.
- Start of follow-up period: In the primary analysis, follow-up was from the end of the manic episode as the ideal construct to investigate the depression transition from, although then the end of the manic/mixed episode may become dependent on frequency of outpatient appointments (e.g., some people are discharged with ongoing mania for further management in the community). In contrast for the TTE, follow-up was from the end of hospitalisation (filtering for hospital length of stay <30 days).
- Total sample: The primary analysis was limited to individuals with both manic/mixed and depressive episodes for reasons explained in the **Supplementary Note**. The TTE considered all manic episodes.

Result

Compared to antipsychotics alone, lithium+antipsychotic during hospitalisation for mania was associated with a significantly lower rate of depression within 30 days of hospitalisation (RR=0.77, 95% CI: 0.59-0.99, p=0.043).

Compared to antipsychotics alone, valproate+antipsychotic during hospitalisation for mania was also associated with a significantly lower rate of depression within 30 days after hospitalisation (RR=0.71, 95% CI: 0.58-0.86, p<0.001).

Our post-hoc comparison of length of hospital stay showed that the combination therapy group had significantly longer hospital stays than the antipsychotic-alone groups. The groups had similar CGI-S scores within 7 days of admission, albeit noting that a minority of episodes (n=616 in the lithium arm; n=761 in the valproate arm) had a CGI-S score available. This could imply some residual confounding that is difficult to account for within the TTE framework where only covariates available at baseline can be accounted for (i.e., not hospital length of stay), but where there is also limited data available to quantify baseline severity (i.e., limited CGI-S data availability).

Most negative control analyses did not identify statistically significant associations, although confidence intervals were generally wide: lithium+antipsychotic on upper respiratory tract infections: RR=1.33, 95%CI: 0.43-2.94, p=0.56; lithium+antipsychotic on allergic rhinitis: RR=1.03, 95%CI: 1.01-3.50; valproate+antipsychotic on allergic rhinitis: RR=0.75, 95%CI=0.10-2.66. The exception was valproate+antipsychotic on upper respiratory tract infections: RR=1.86, 95%CI=1.03-3.66.

Overall, these results (**Table S14**) are consistent with the primary results displayed in **Figure 4**. We note also that the lithium group had less than half the sample size compared to the valproate group, which contributes to the wider confidence intervals observed.

**Negative control outcomes**

We defined a composite negative control outcome and repeated all analyses to compare how these behave to depression. We chose two common physical health conditions within this composite:

1. Upper respiratory tract infections (URTI).
2. Allergic rhinitis. This outcome is helpful to assess specificity, as it should be a positive control for antihistamines and a negative control for the other medications.

Cumulative incidence (**Figure S11**; compare to **Figure 1a**): Incidence of URTIs and allergic rhinitis is much more gradual after mania/mixed episodes than the incidence of depression (**Figure S33**).

Excess rate (**Figure S12**; compare to **Figure 1b**): These results suggest little relation between rates of diagnosing these physical health conditions and time soon after manic and mixed episodes.

Association with clinical features (**Figure S19**; compare to **Figure 2**): After mania, there was a nominal association with previous short depression transition time, and after mixed episodes, there was a significant association after multiple testing correction, but the magnitude of this effect was substantially less than in the primary analysis. This may reflect increased health-seeking behaviour.

Association with medications for manic/mixed episodes (**Figure S31**; compare to **Figure 3**): There were no associations between medications prescribed within the first 7 days of manic/mixed episodes and the composite negative control outcome in the 1 month from mania/mixed episode end. However, the error bars were wide.

Thus, this negative control analysis indicates that our results relating to short depression time may in part be related to patterns in healthcare contacts, but that this is unlikely to be driving the results in entirety.

**Sensitivity analysis exploring recency of last prescription on the medication results**

We defined a “manic/mixed start period”, as the 7 days before the coded manic/mixed episode. This was chosen to be consistent with our medication exposure definition (within +/- 7 days of manic/mixed episode start), and acknowledges that coding may be delayed from true onset. Looking backwards in time from this manic/mixed start period, we quantified the days since the last medication prescription.

First, we plotted per drug/class the distribution of days since the last prescription in the 90 days before the manic/mixed start period. Note that prescriptions that spanned the manic/mixed start period were assigned to time=0. In general, prescription rates in the 90 days preceding the manic/mixed start period were low. Lithium was particularly unlikely to be prescribed in the 90 days before the manic/mixed start period compared to other anti-manic medications. In contrast, lamotrigine was relatively more likely to be prescribed (**Figure S30a**).

Second, we performed a sensitivity analysis for the medication models. We modelled medication exposures as a three-level factor:

1. Prescription within +/- 7 days of the mania/mixed episode, but with another prescription within the 90 days before the manic/mixed start period
2. Prescription within +/- 7 days of the mania/mixed episode, but with no prescription recorded within the 90 days of the manic/mixed start period (indicating re-starting at the time of the manic/mixed episode)
3. No prescription

We considered this a sensitivity analysis to assess consistent direction of effect. This is because this analysis essentially splits the medication sub-groups, and so has lower power than the primary analysis. For this reason, we did not include multiple testing correction.

These results indicate that for lithium, valproate and antipsychotics, there were consistent effects whether the medication was prescribed prior to the manic/mixed episode or started contemporaneously (**Figure S30b**).

**Multiple testing**

Here we delineate the multiple testing burden used in Bonferroni correction for each analysis. Broadly, we applied multiple testing within each major analysis and considered manic and mixed episodes separately:

- Clinical factors analysis: Number of tests = 3 (hospitalisation for the manic/mixed episode, severity, past history of short mania/mixed-to-depression transition ≤1 month).
- Medication class analysis: Number of tests = 6 (antipsychotics, antiepileptic mood stabilisers, lithium, antidepressants, antihistamines and benzodiazepines).
- Depression severity analysis: 3 per analysis (depression-related hospitalisation, all-cause hospitalisation, PHQ-9 total score).

A per-analysis multiple testing approach is appropriate as outlined previously^5^.

### **Supplementary Figures**

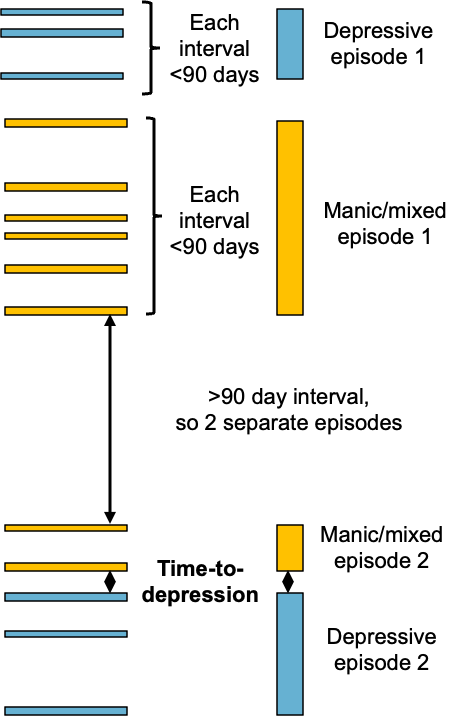

**Figure S1**: Schematic illustrating collapse of time-stamped diagnoses into episodes.

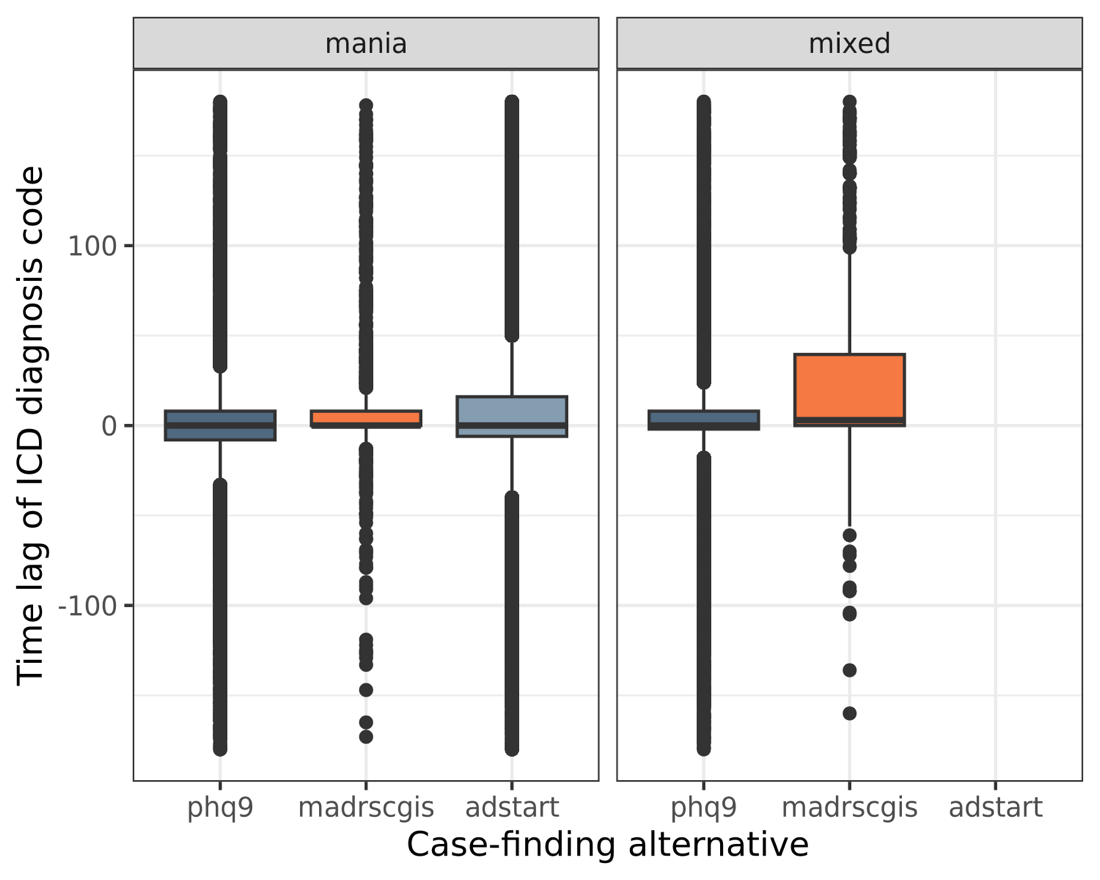

**Figure S2:** Difference in mania/mixed-to-depression transition time detection between ICD diagnosis codes and broader case-finding criteria in days. Additional case-finding measures shown here include PHQ-9 (patient-report questionnaire), MADRS/CGI-S (derived variable from the Neuroblu database inferring MADRS score from CGI-S) and antidepressant start date (mania only, as antidepressants may be prescribed during a mixed episode, so should not be used to mark the start of depression).

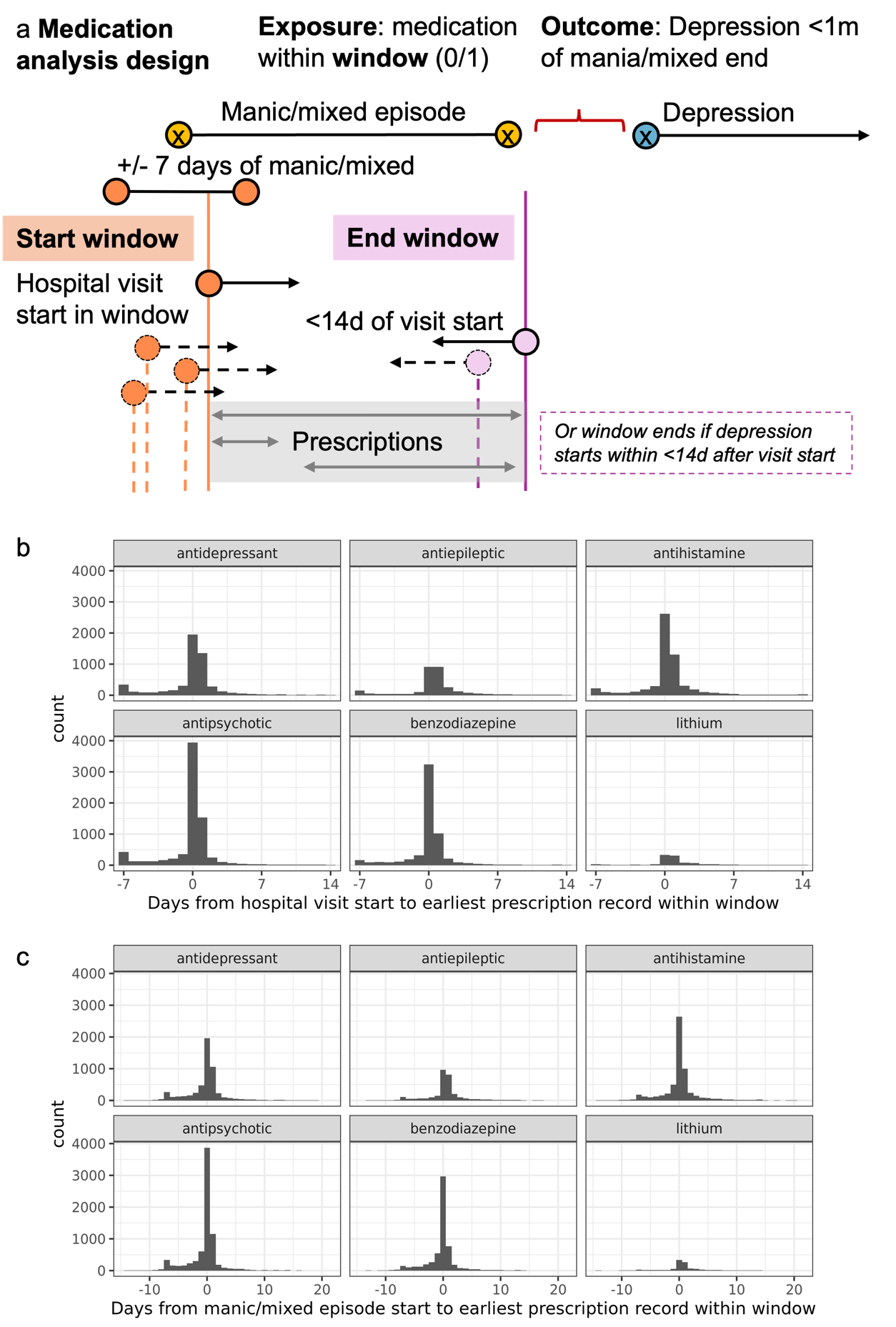

**Figure S3**: Definition of hospital-associated manic/mixed episodes and relation to prescription. a) Diagram of exposure window studied in the analysis of medications prescribed during manic/mixed episodes associated with a visit (e.g., hospital), and their relationship with the outcome mania/mixed-to-depression transition time. b) Histogram of days from hospital visit start to the earliest record of prescription for each drug broad class within the window. c) Histogram of days from manic/mixed episode start to the medication prescription.

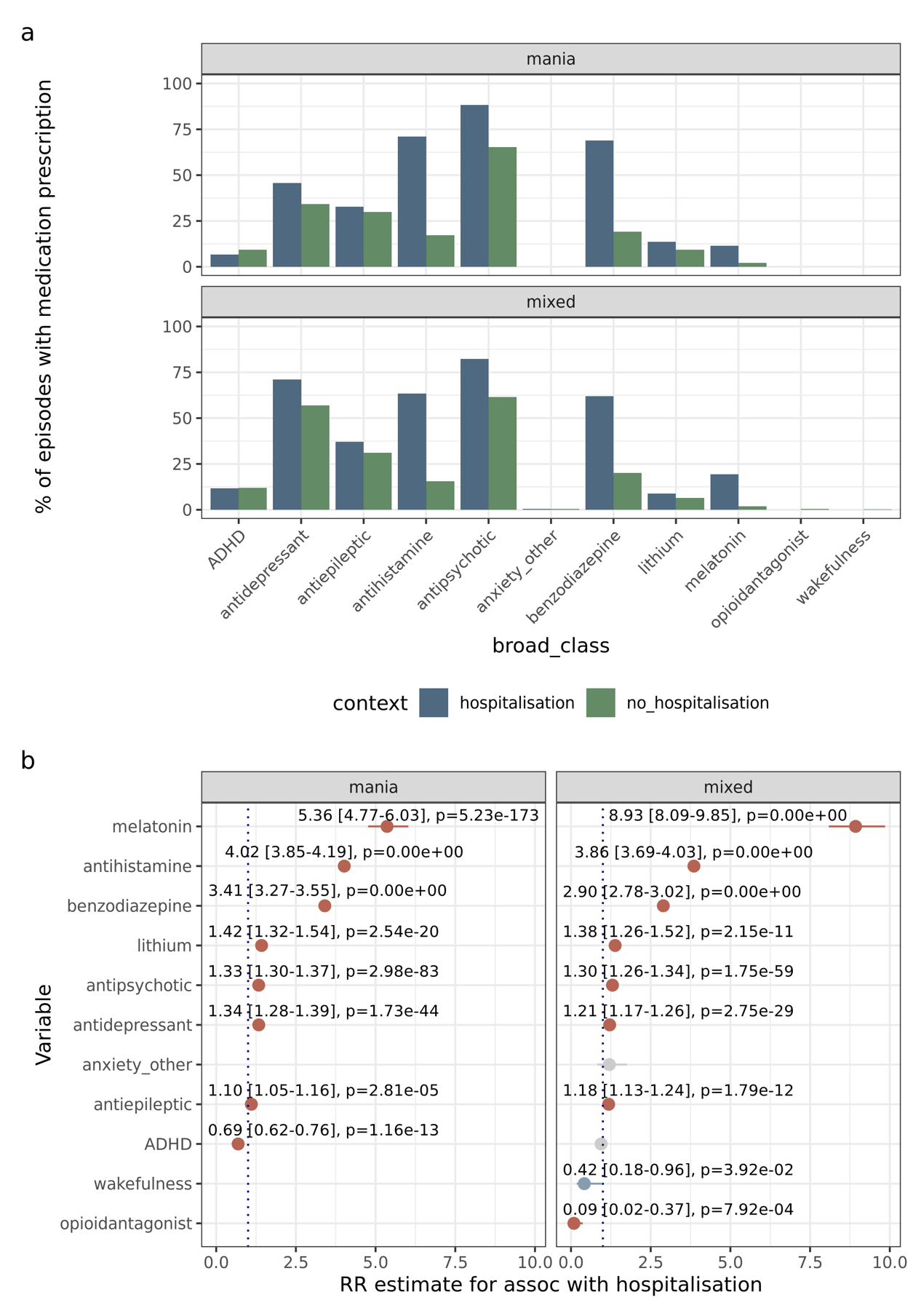

**Figure S4**: Relationship between medication prescription and hospitalisation. a) Barchart for percentage of episodes with each broad medication class prescribed for manic/mixed episodes associated with hospital (blue) versus not associated with hospital (green). b) Coefficients from modified Poisson regression for association between broad class medication prescription and hospitalisation. Plotted here is the overlapping subset of 65,038 episodes from 36,425 individuals with both medication and visit data, and for states with hospitalisation rate ≥1%.

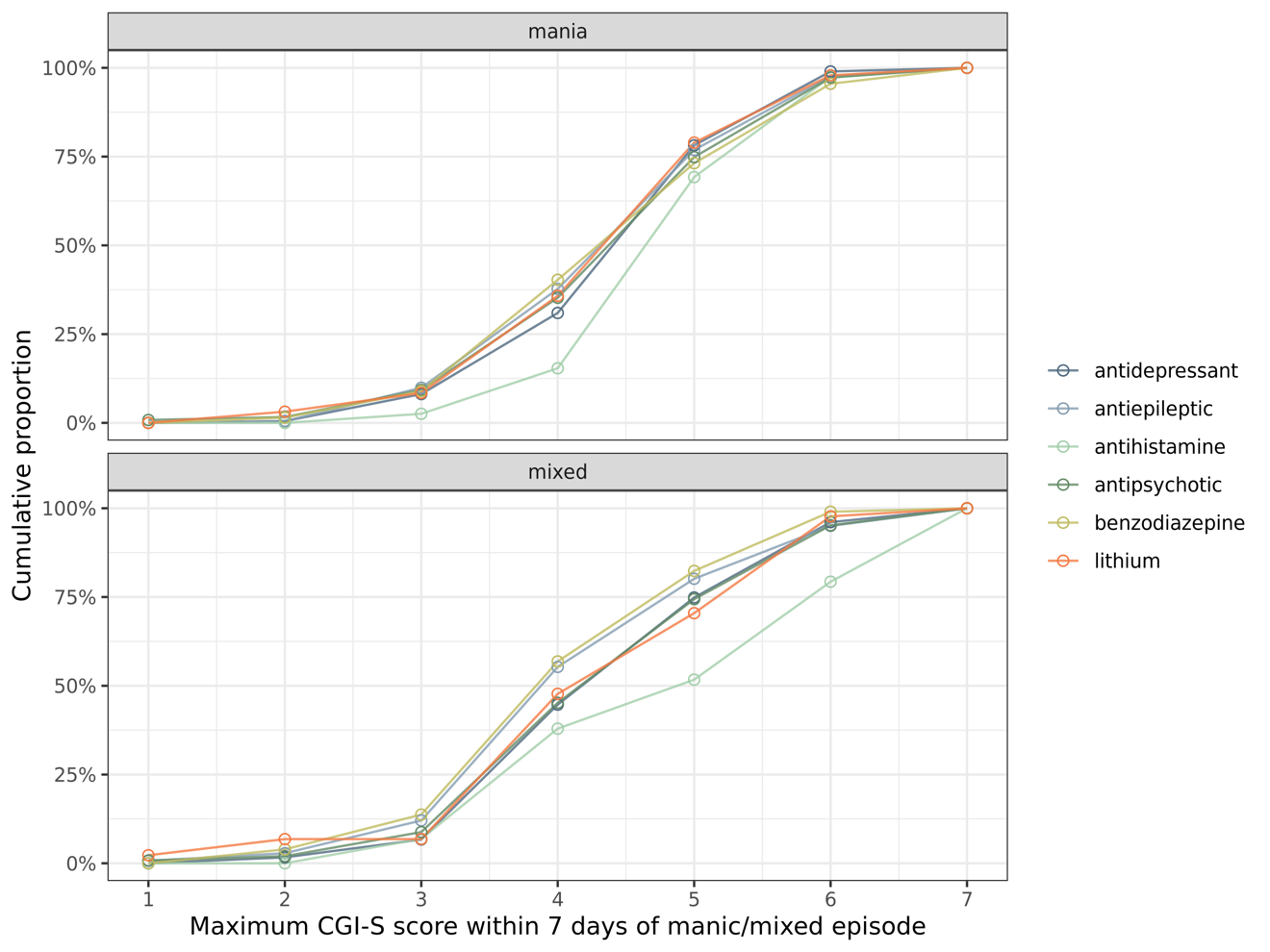

**Figure S5**: Relationship between medication prescribed within 7 days of the manic/mixed episode start date and the distribution of contemporaneous CGI-S scores. Plotted here is the overlapping subset of 9.086 episodes from 991 individuals with both medication data and CGI-S scores within 7 days of hospitalisation.

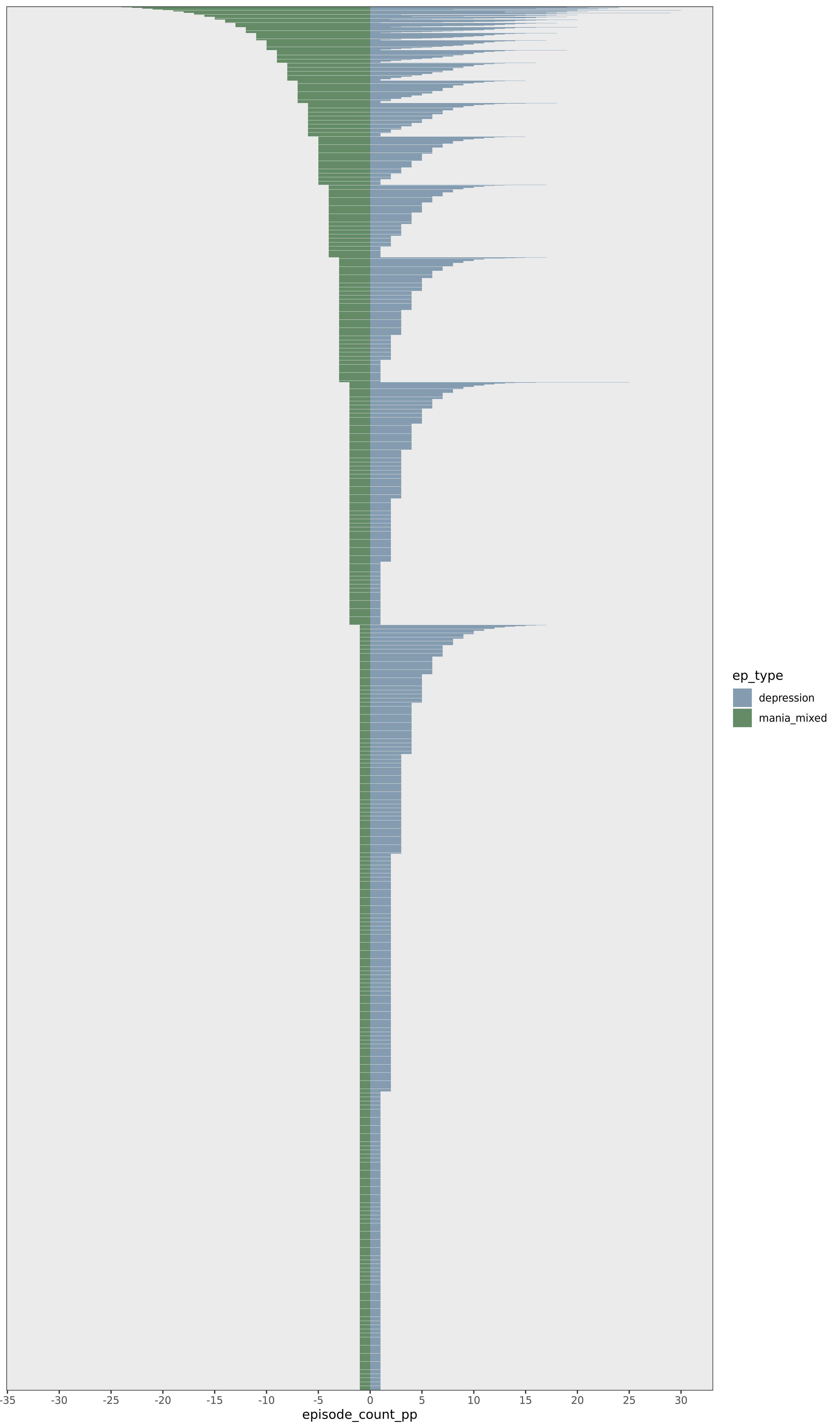

**Figure S6**: Number of manic/mixed and depressive episodes per individual identifier recorded in this dataset.

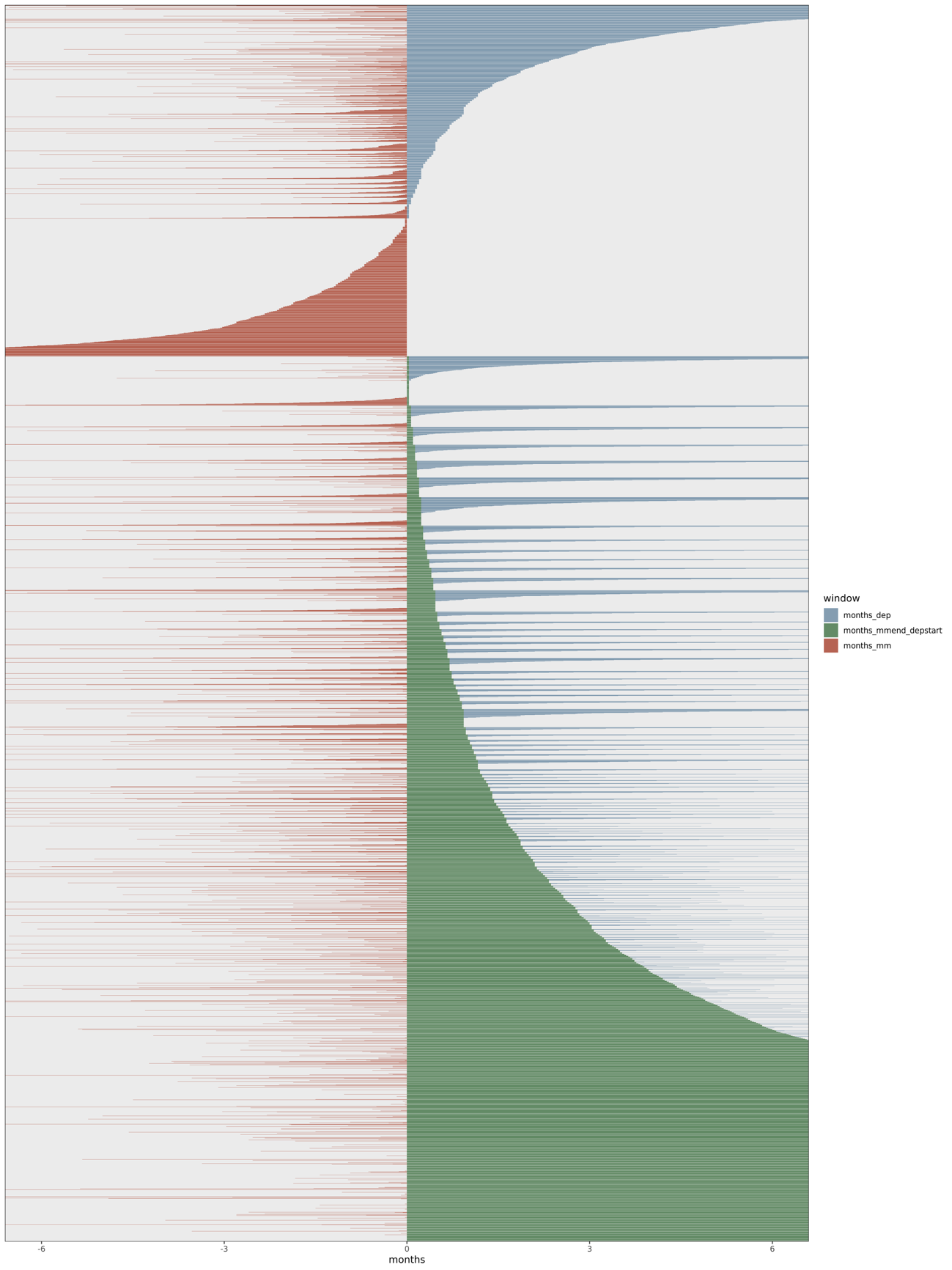

**Figure S7**: Durations (months) of each of the following phases related to the mania/mixed-to-depression transition: preceding mania/mixed episode (red), between mania/mixed and depression (green), subsequent depression episode (blue). Rows are ordered by duration between the last mania/mixed diagnosis and the next depression diagnosis, without intervening mania/mixed episodes. Lack of bars: where the apposing episodes/windows occurred on the same day. Data truncated to 6 months pre- and post- depression.

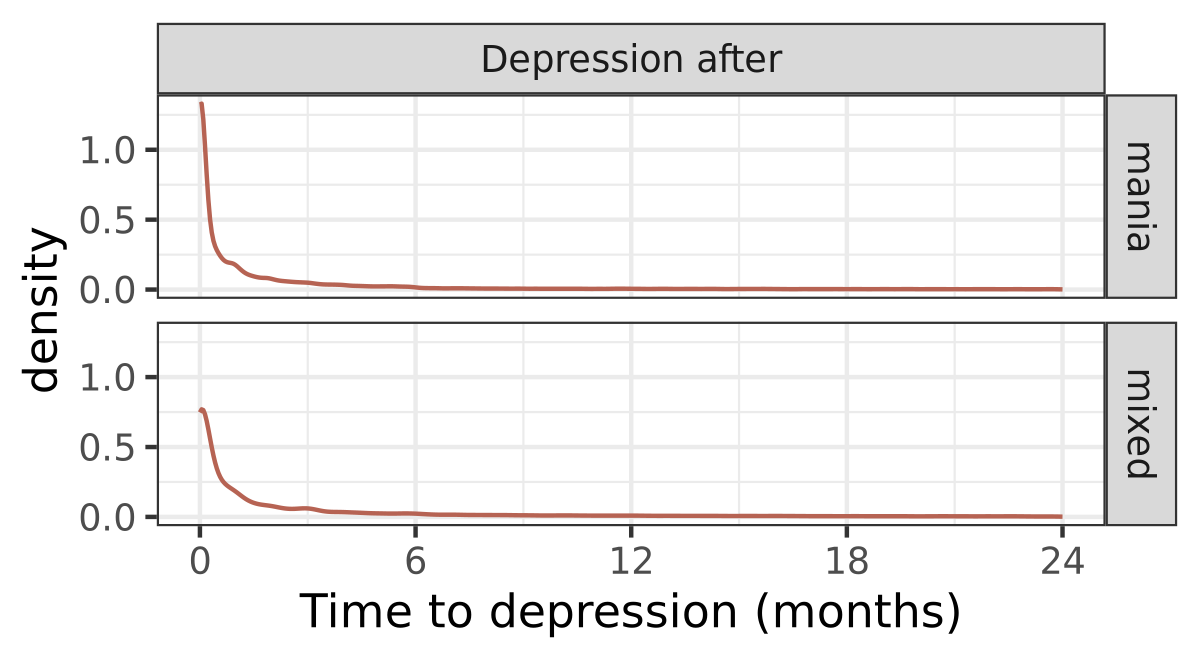

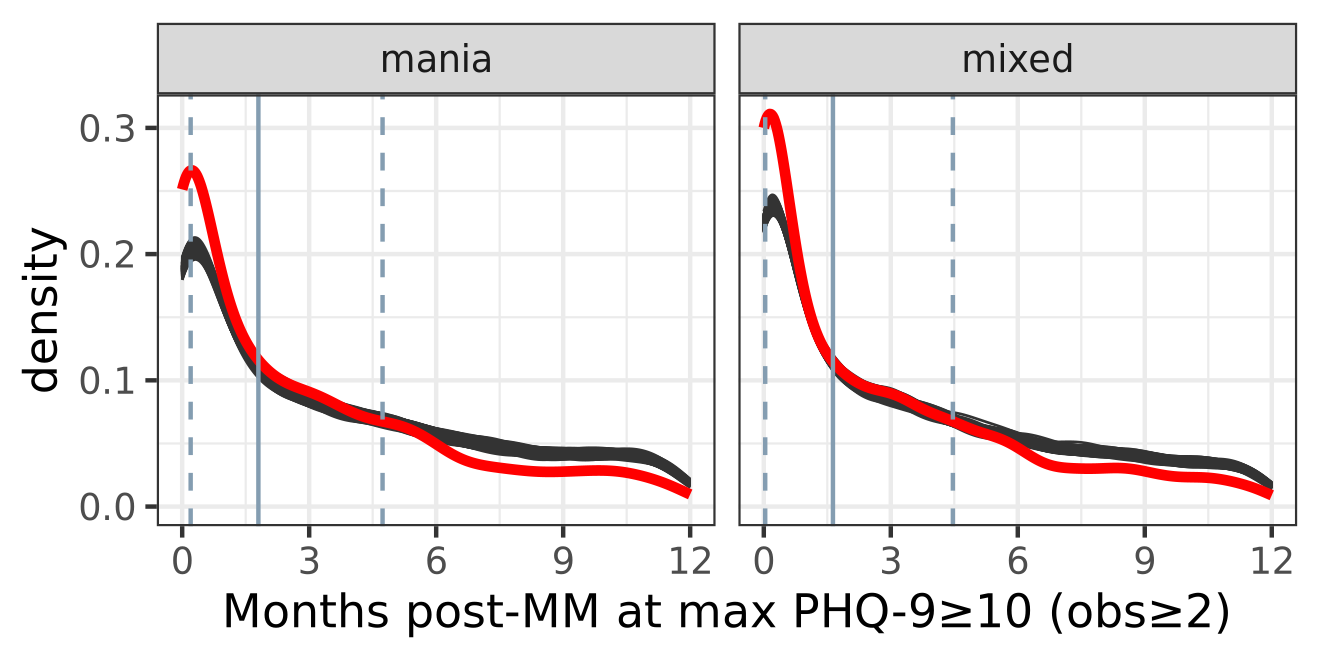

**Figure S8**: Density plots indicating peak incidence and peak severity of depression after mania/mixed episodes. (a) Density plot of time to depression after manic and mixed episodes. Data from 208,825 manic/mixed episodes from 100,776 individuals. (b) Density plot for the time to maximum severity of depression (red line), relative to n=1,000 permutations to generate an empirical null distribution (black lines) and the interquartile range of the null distribution (blue vertical lines). 11,669 episodes from 9,165 individuals.

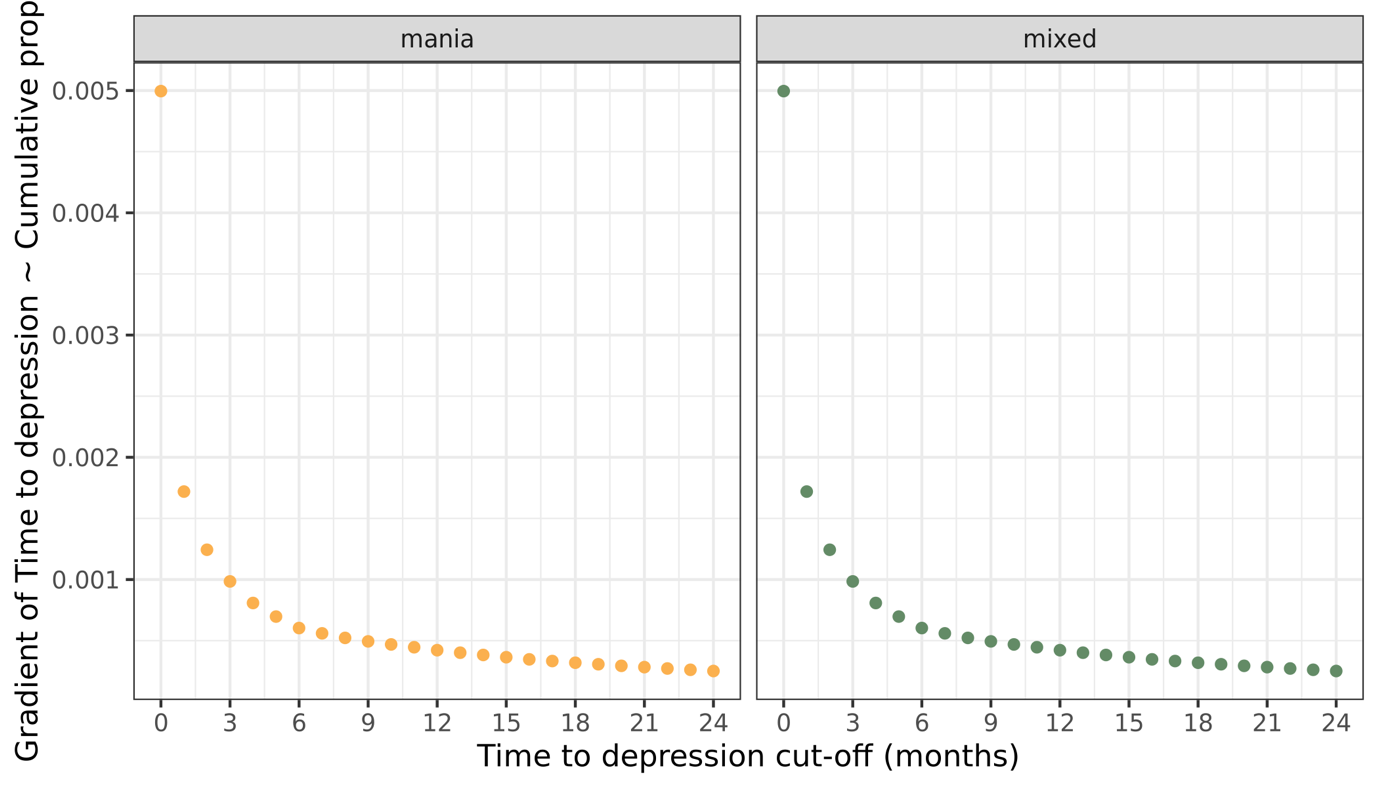

**Figure S9**: Plot of gradient (beta coefficient) of time to depression regressed against cumulative proportion of individuals as plotted in **Figure 1a**. Note that the gradient begins to stabilise around the 6 month mark.

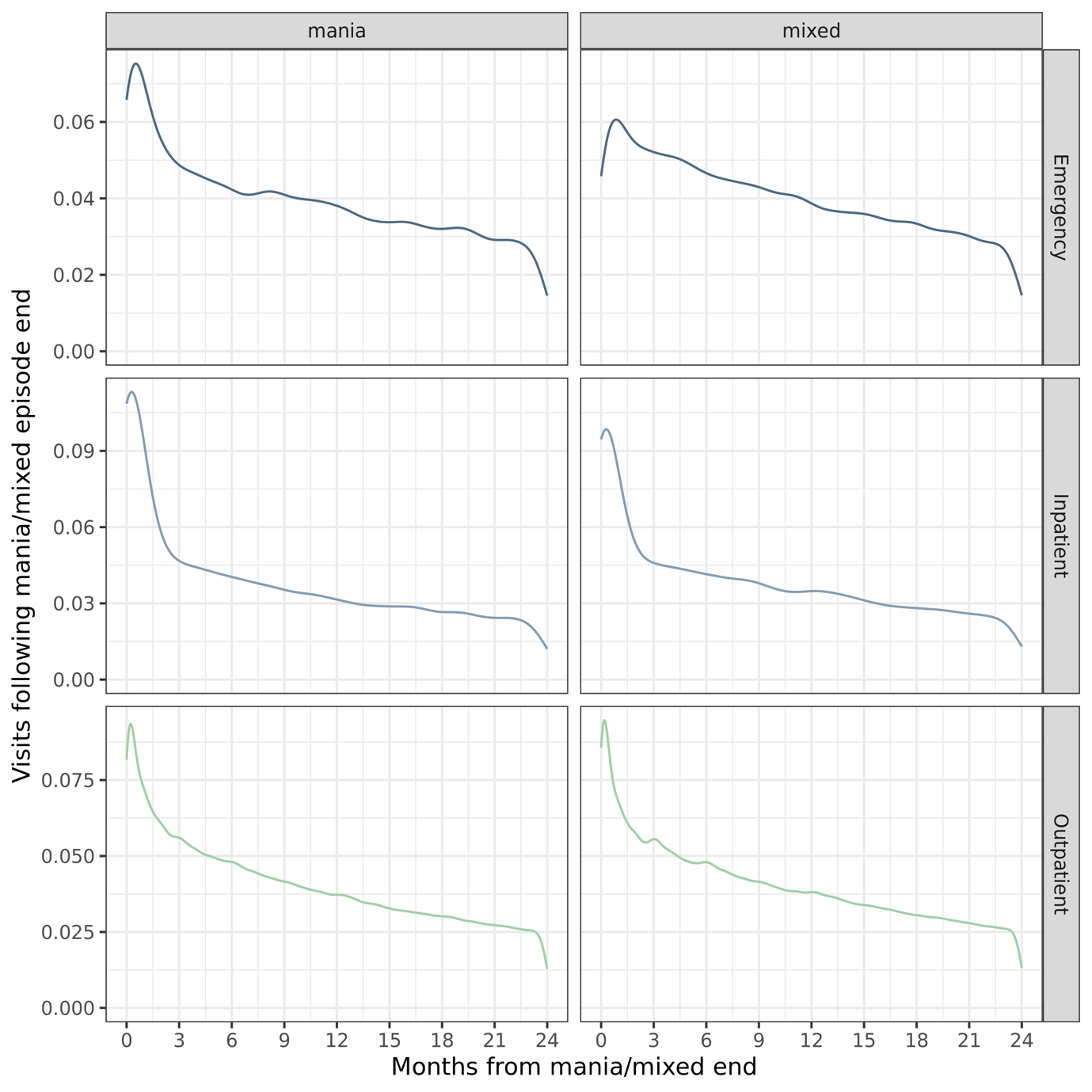

**Figure S10**: Density plot of the temporal distribution of visits following the end of manic/mixed episodes, stratified by visit type.

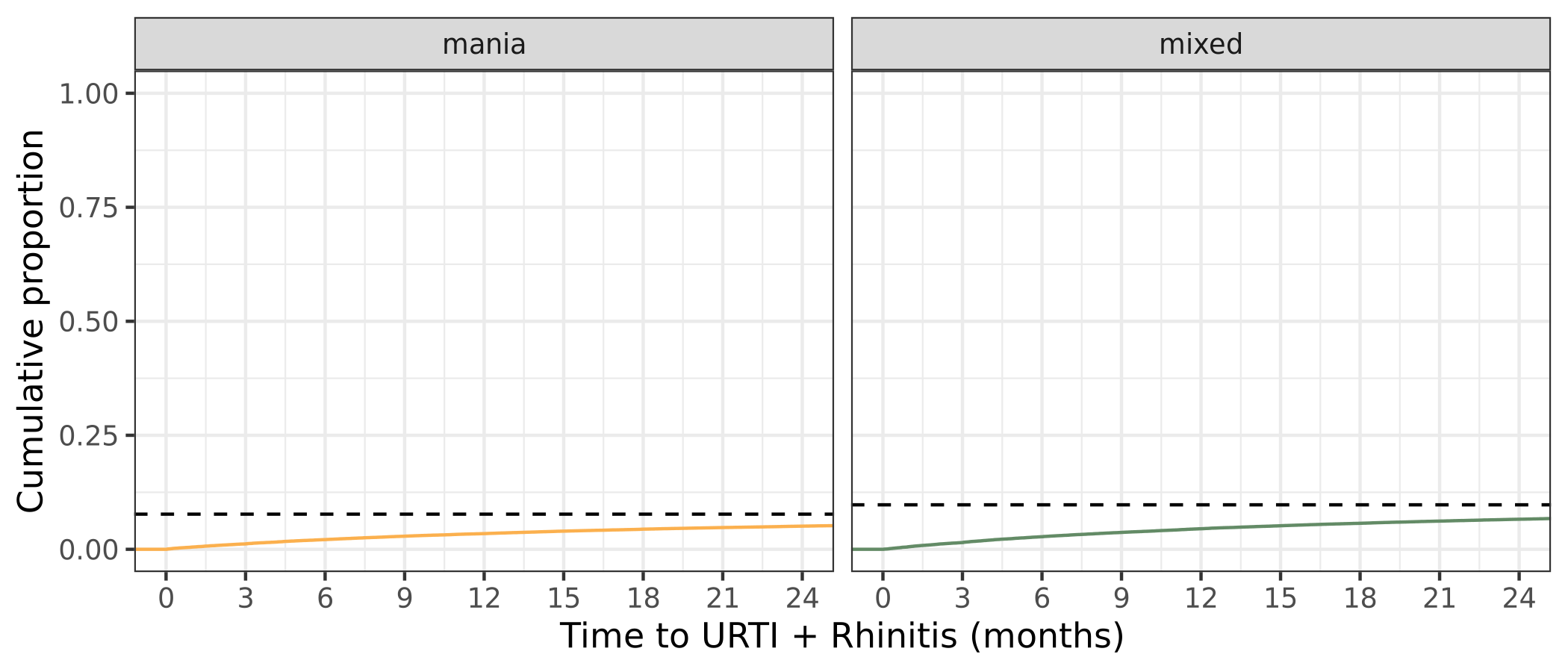

**Figure S11**: Cumulative proportional incidence of a) upper respiratory tract infections (URTI) and b) allergic rhinitis codes after manic/mixed episodes over time. Dashed line: proportion of all participants who have a ICD code for these negative control outcomes after a manic/mixed episode.

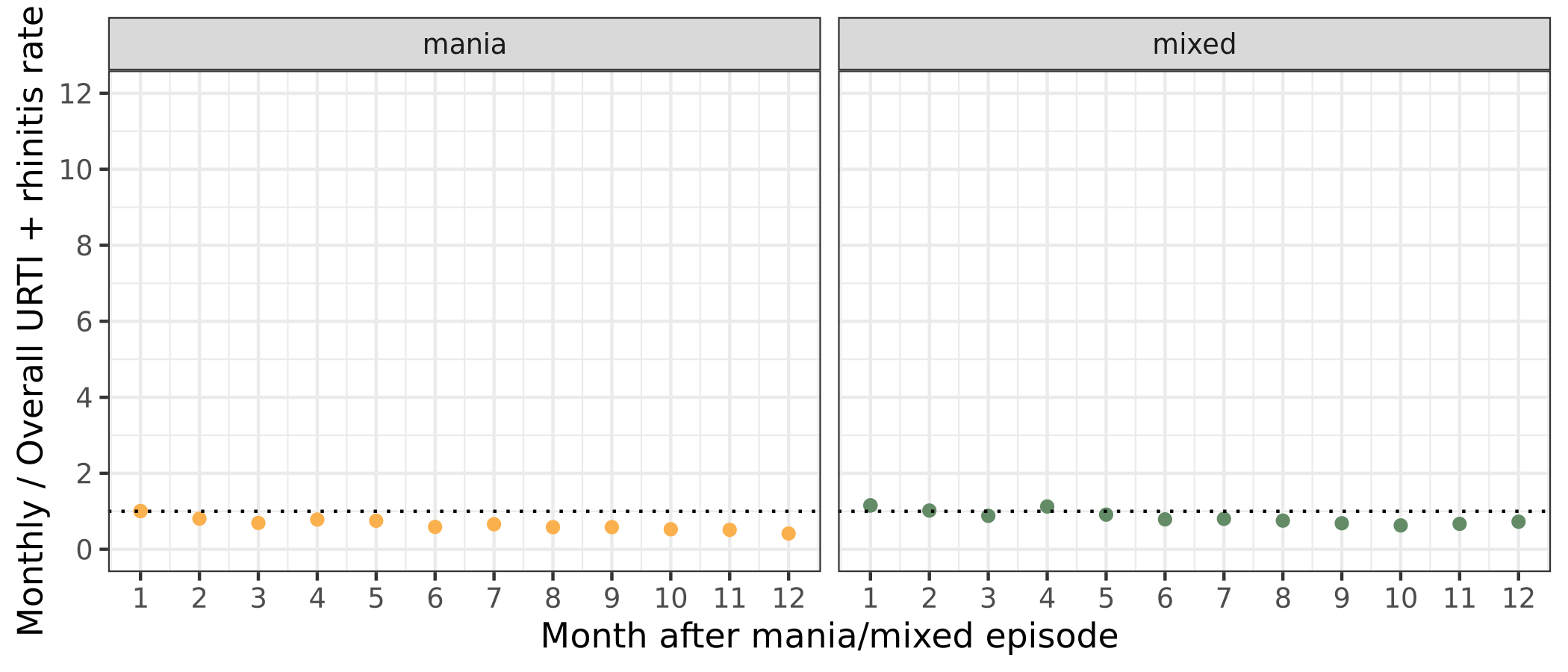

**Figure S12**: Ratio of per-month rate of a) upper respiratory tract infections (URTI) and b) allergic rhinitis diagnosis, relative to the overall rate of diagnoses in this bipolar sample. Dotted line: rate ratio=1.

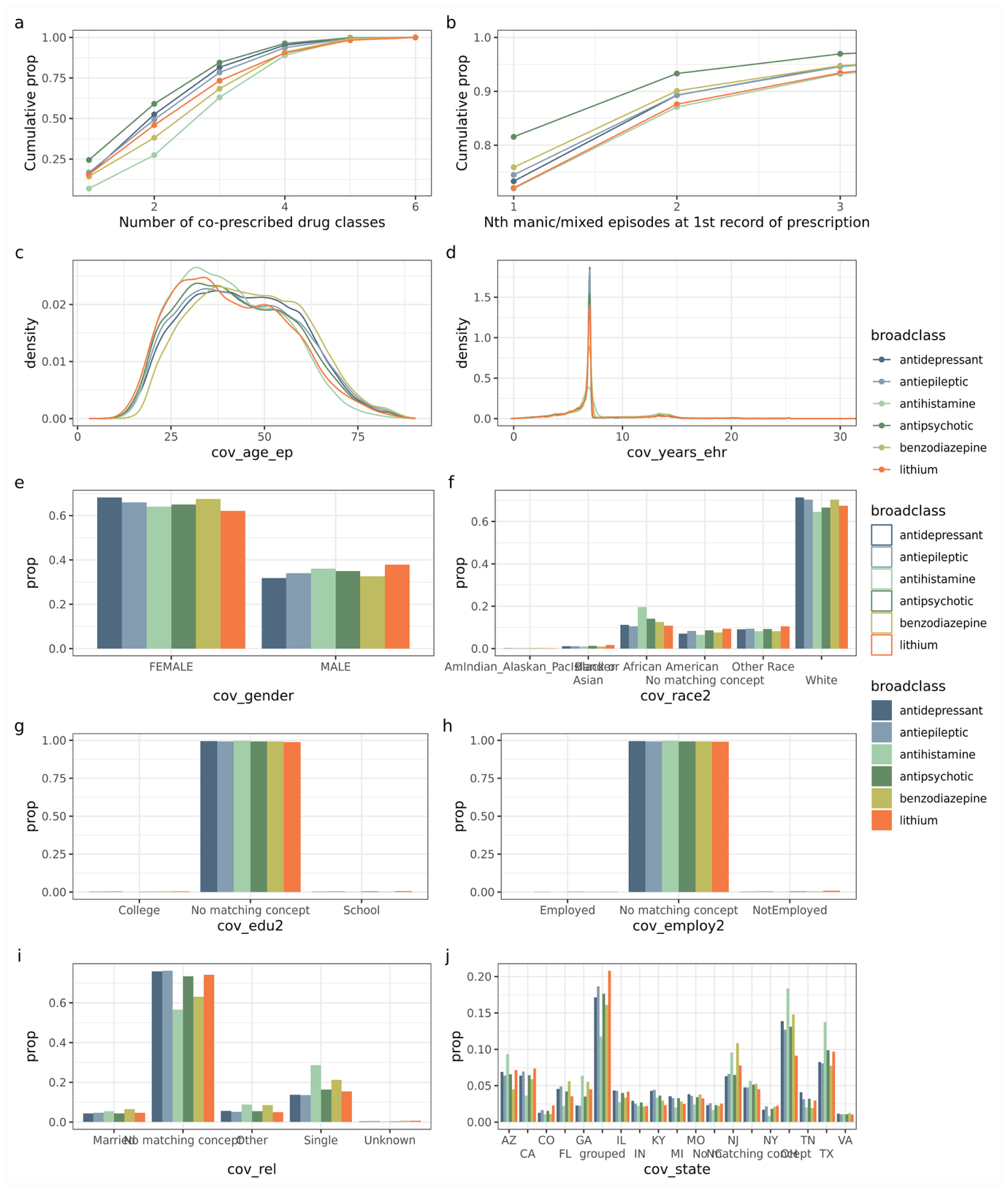
**Figure S13**: Relationships between prescribing and demographic factors and prescription of major psychotropic medication classes for manic/mixed episodes, from all episodes in this sample of individuals with bipolar disorder. a) Per drug-class cumulative proportion of manic/mixed episodes with increasing degrees of polypharmacy. b) Earliest EHR-recorded manic/mixed episode per-individual when each drug class was prescribed. Proportions are calculated within each drug class. Density plots of c) age at episode and d) years in EHR. Bar plots of categories for e) gender, f) race, g) educational attainment, h) employment status, i) relationship status, j) state.

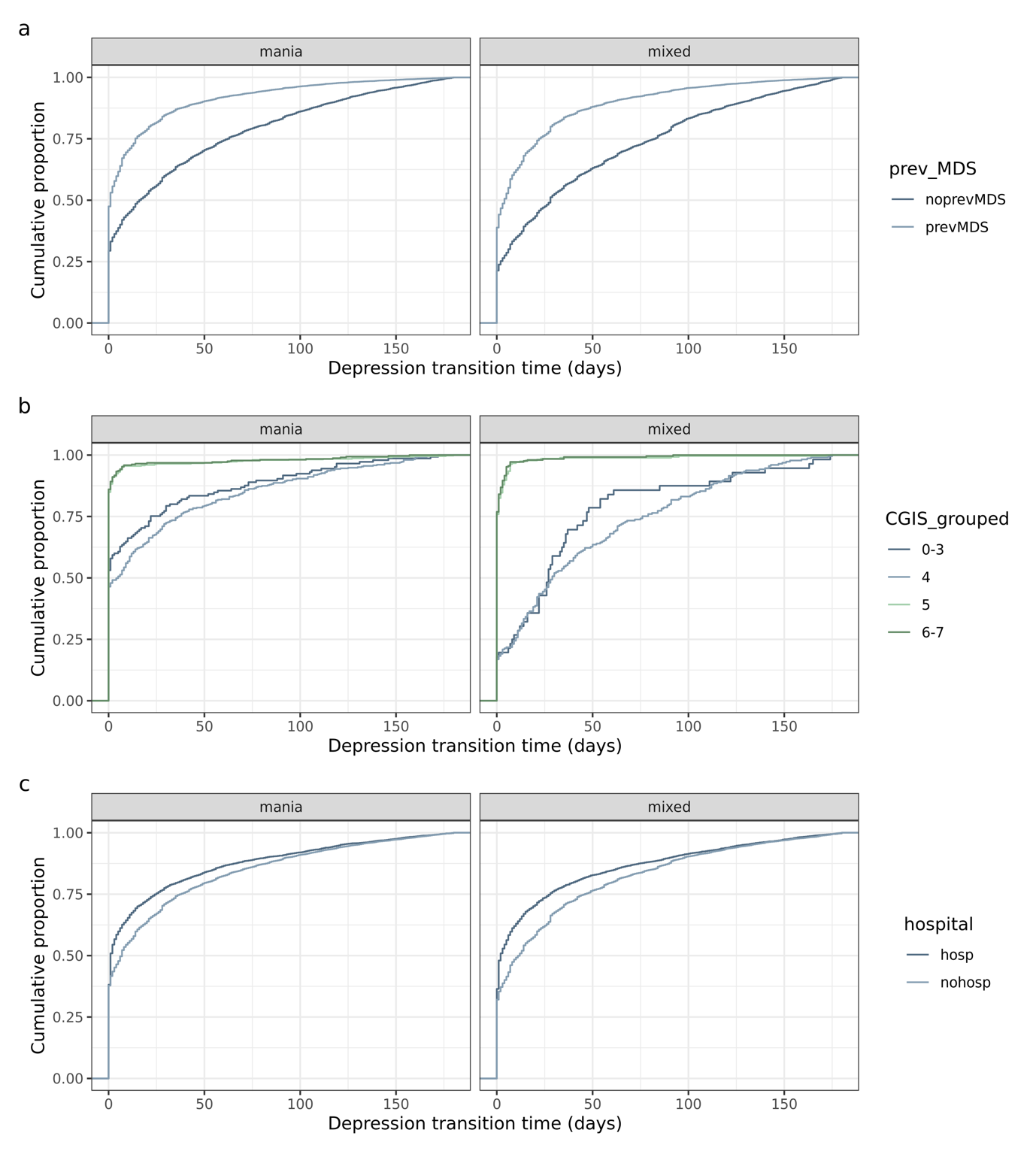

**Figure S14**: Cumulative density function plots of mania/mixed-to-depression transition time, stratified by clinical factors. a) previous short mania/mixed-to-depression transition time within 1 month (108,389 episodes from 40,082 individuals); b) CGI-S severity (2,470 episodes from 1,619 individuals); c) hospitalisation for the manic/mixed episode (104,663 episodes from 51,817 individuals). CGI-S scale has been grouped so that each group has at least 50 observations where CGI-S scale is 1 = normal, 2 = borderline, 3 = mildly ill, 4 = moderately ill, 5 = markedly ill, 6, severely ill, 7 = among the most extremely ill patients.

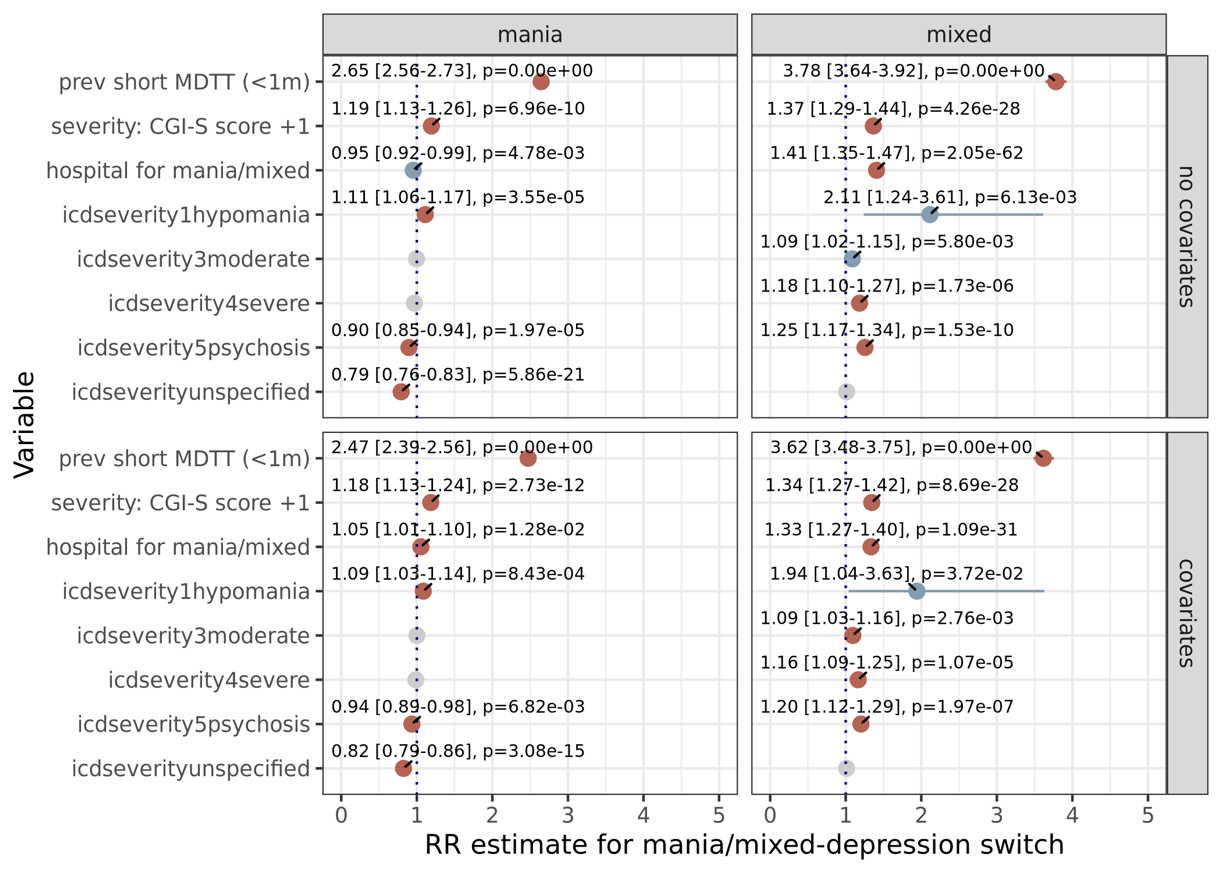

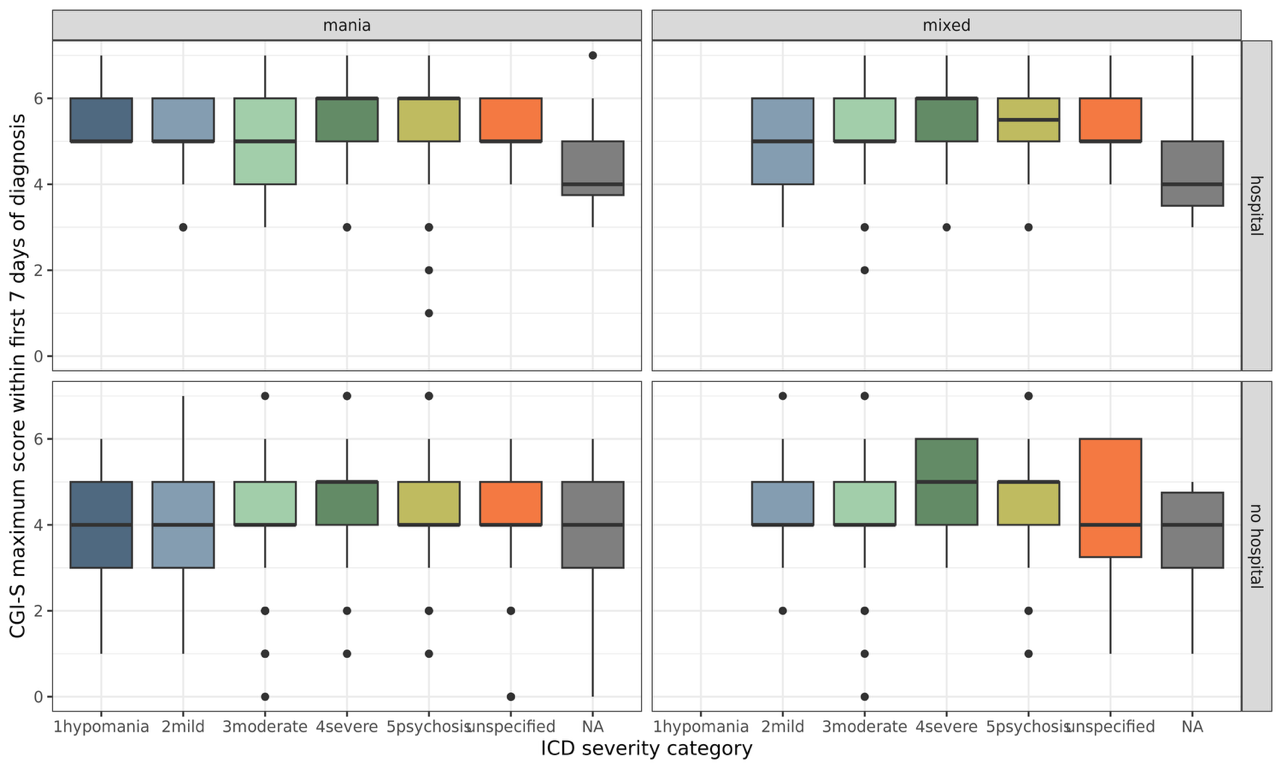

**Figure S15**: Clinical factors associated with short mania/mixed-to-depression transition time. (upper) Coefficient plot for the effect of clinical factors available at manic/mixed episode diagnosis on the risk ratio of short mania/mixed-to-depression transition time (“MDTT”) within 1 month, using models with and without covariates. Sample size: hospitalisation analysis based on 104,663 episodes from 51,817 individuals, filtering on states with a ≥1% hospitalisation rate; CGI-S analysis based on from 2,470 episodes from 1,619 individuals; previous short mania/mixed-to-depression transition time (“MDTT”) analysis was performed on 108,389 episodes from 40,082 individuals; ICD severity analysis on 200,558 episodes from 96,970 individuals. Covariates: age_episode_ + age_EHR_ + years_EHR_ + gender + state + relationship status + employment status + education status (except for the joint model with CGI-S due to model singularity issues where employment status and state were excluded). Red points indicate significance after mulitple testing correction (3 tests; p<0.017); blue indicate nominal significance (p<0.05). (lower) Relationship between ICD-coded severity and CGI-S rated severity of manic/mixed episodes indicates that CGI-S better captures severity related to hospitalisation.
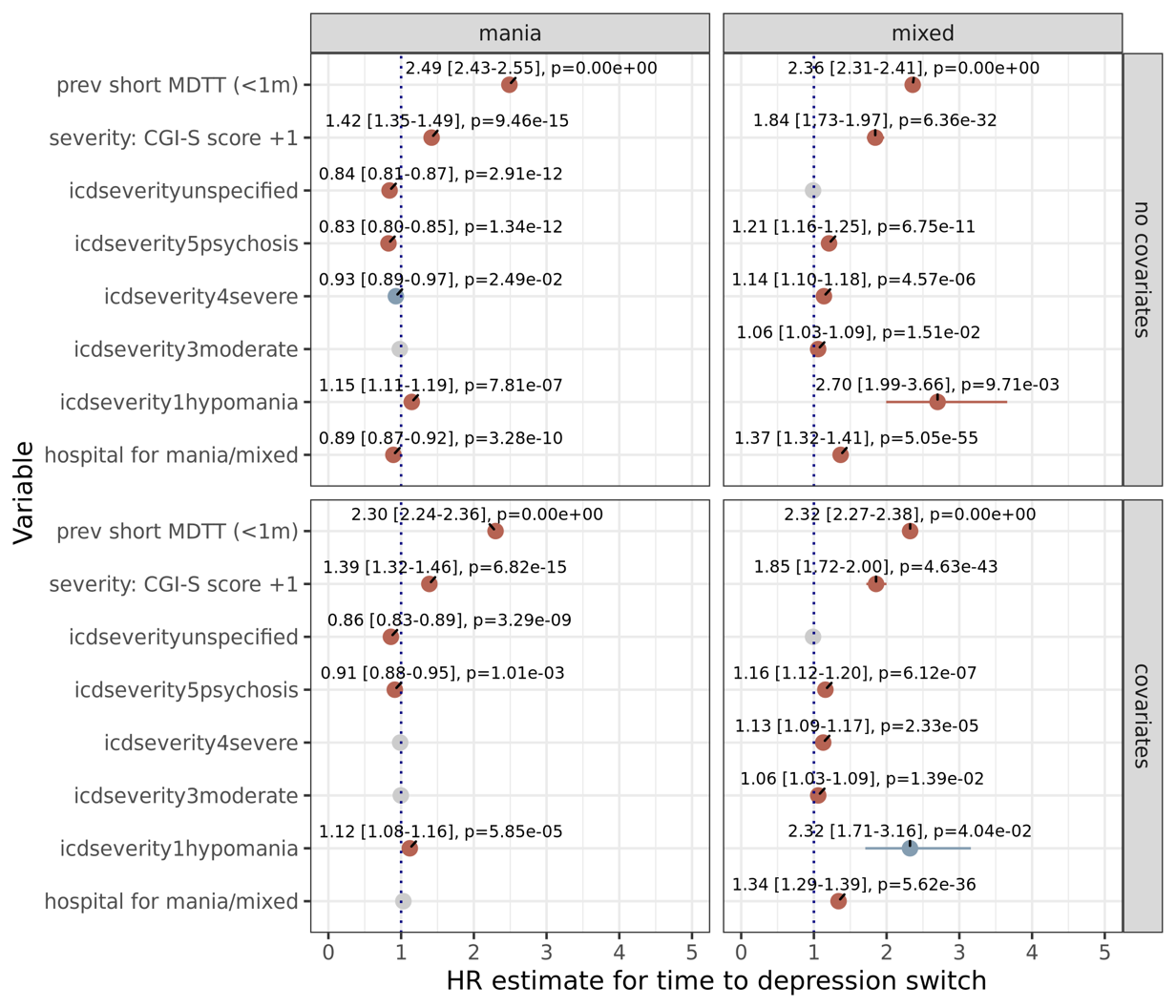

**Figure S16**: Coefficient plot for the effect of clinical factors available at manic/mixed episode diagnosis on the cause-specific hazard ratio for mania/mixed-to-depression transition time, for analyses with and without covariates, censored to a 6 month window. ICD severity coefficients are presented relative to mild severity. Sample size: hospitalisation analysis based on 104,663 episodes from 51,817 individuals, filtering on states with a ≥1% hospitalisation rate; CGI-S analysis based on from 2,470 episodes from 1,619 individuals; previous short mania/mixed-to-depression transition time (“MDTT”) analysis was performed on 108,389 episodes from 40,082 individuals; ICD severity analysis on 200,558 episodes from 96,970 individuals. Covariates: age_episode_ + age_EHR_ + years_EHR_ + gender + state + relationship status + employment status + education status (except for the joint model with CGI-S due to model singularity issues where employment status and state were excluded). Red points indicate Bonferroni-significance (8 tests; p<0.00625); blue indicate nominal significance (p<0.05).

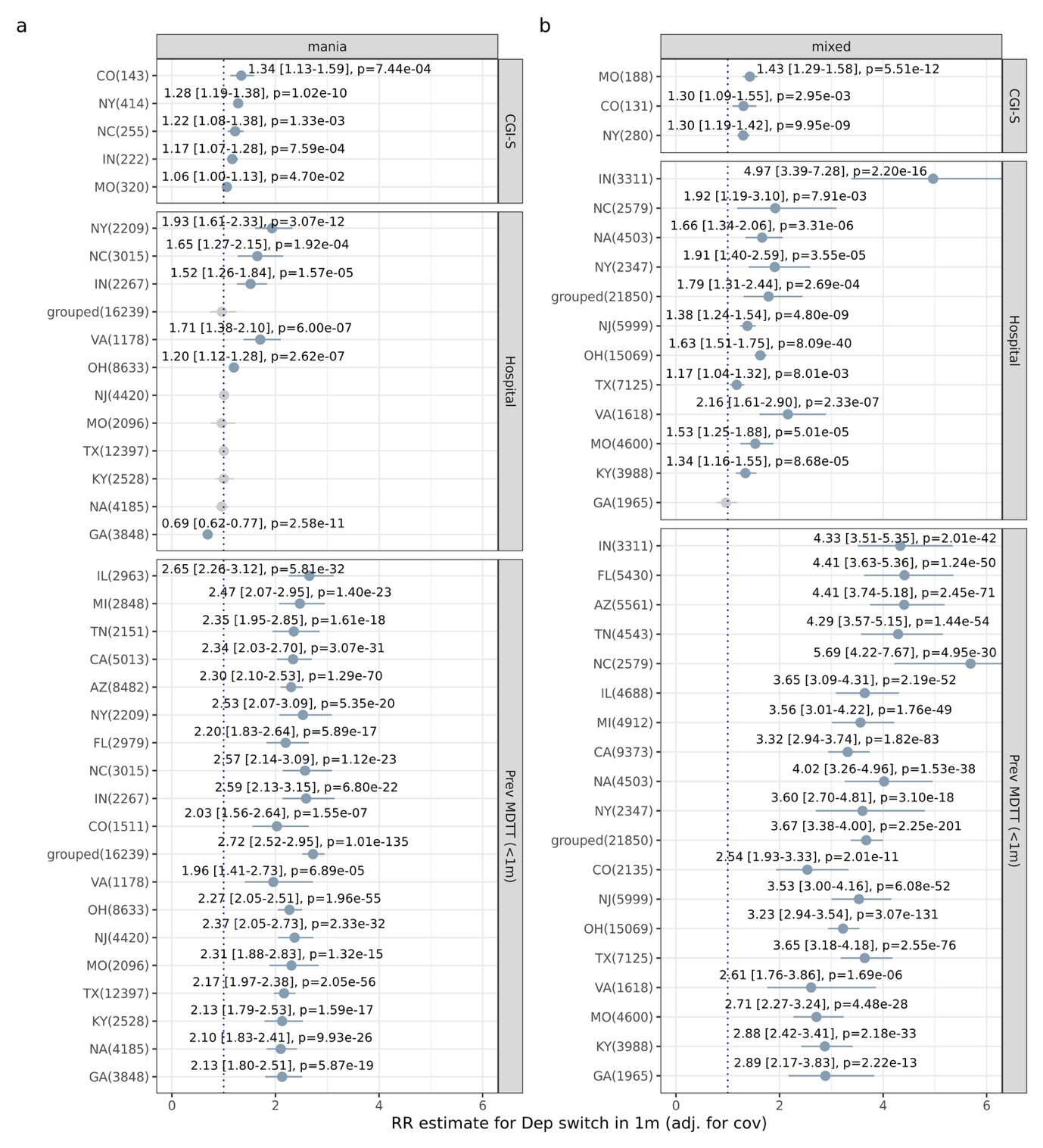

**Figure S17**: Clinical factors sensitivity analysis stratifying by state of residence. Rate ratio (±95% CI) for listed clinical factors and their relationship with short mania/mixed-to-depression transition (≤1 month; modified Poisson regression with within-individual clustered standard errors). Analyses here jointly include all listed medication classes and covariates: age_episode_ + age_EHR_ + years_inEHR_ + gender. Blue indicates p<0.05. Y-axis labels include number of episodes contributed by each state; each state had to contribute >50 episodes to be included as its own group.

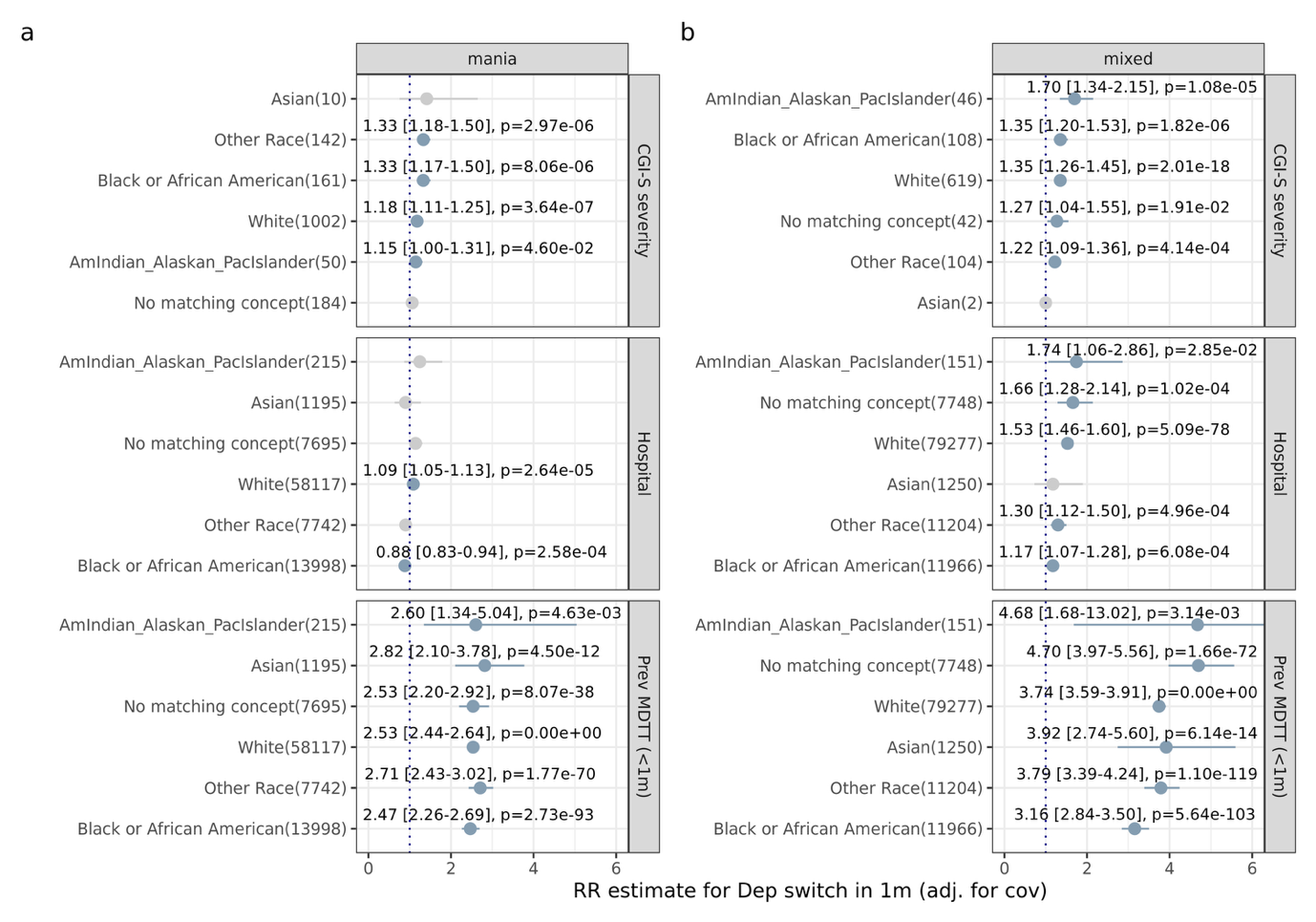

**Figure S18**: Clinical factors sensitivity analysis stratifying by recorded racial group. Rate ratio (±95% CI) for clinical factors and their relationship with short mania/mixed-to-depression transition (≤1 month; modified Poisson regression with within-individual clustered standard errors). Analyses here jointly include all listed medication classes and covariates: age_episode_ + age_EHR_ + years_inEHR_ + gender + state. Blue indicates p<0.05. Y-axis labels include number of episodes contributed by each racial group.

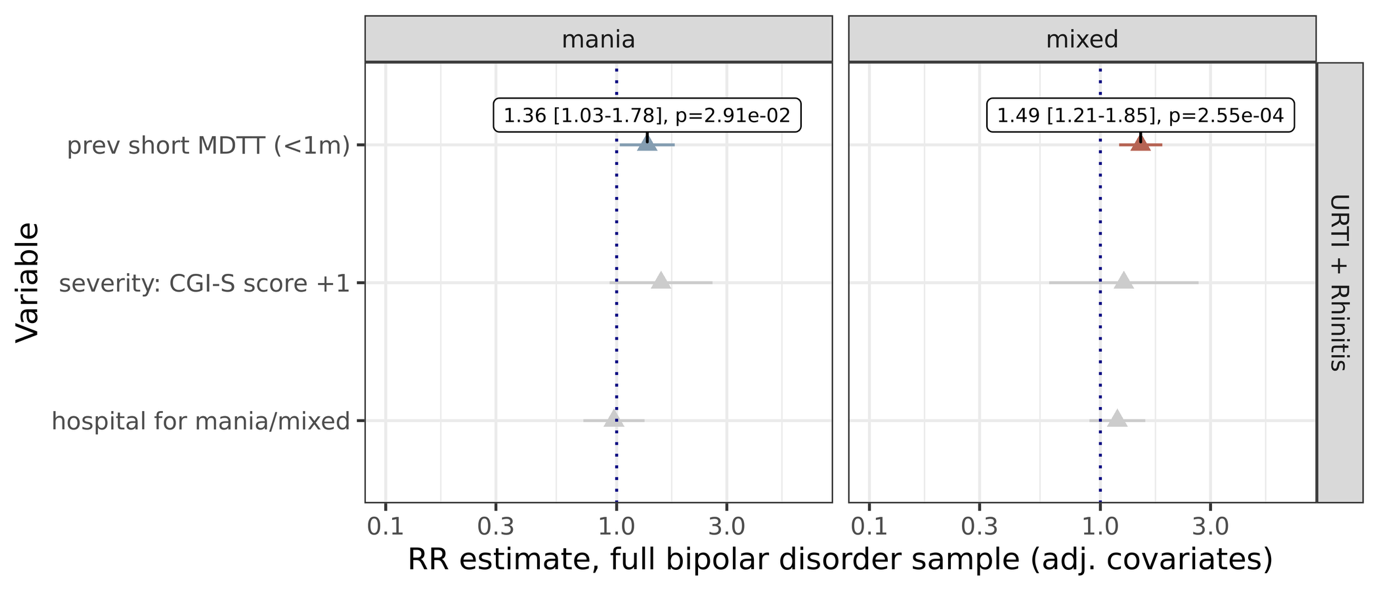

**Figure S19**: Coefficient plot for the effect of clinical factors available at manic/mixed episode diagnosis on the risk ratio of diagnosis of allergic rhinitis (upper) and upper respiratory tract infections (lower) within 1 month. Sample size: hospitalisation analysis based on 104,663 episodes from 51,817 individuals, filtering on states with a ≥1% hospitalisation rate; CGI-S analysis based on from 2,470 episodes from 1,619 individuals; previous short mania/mixed-to-depression transition time (“MDTT”) analysis was performed on 108,389 episodes from 40,082 individuals; ICD severity analysis on 200,558 episodes from 96,970 individuals. Covariates: age_episode_ + age_EHR_ + years_EHR_ + gender + state + relationship status + employment status + education status (except for the joint model with CGI-S due to model singularity issues where employment status and state were excluded). Blue points: nominal significance.

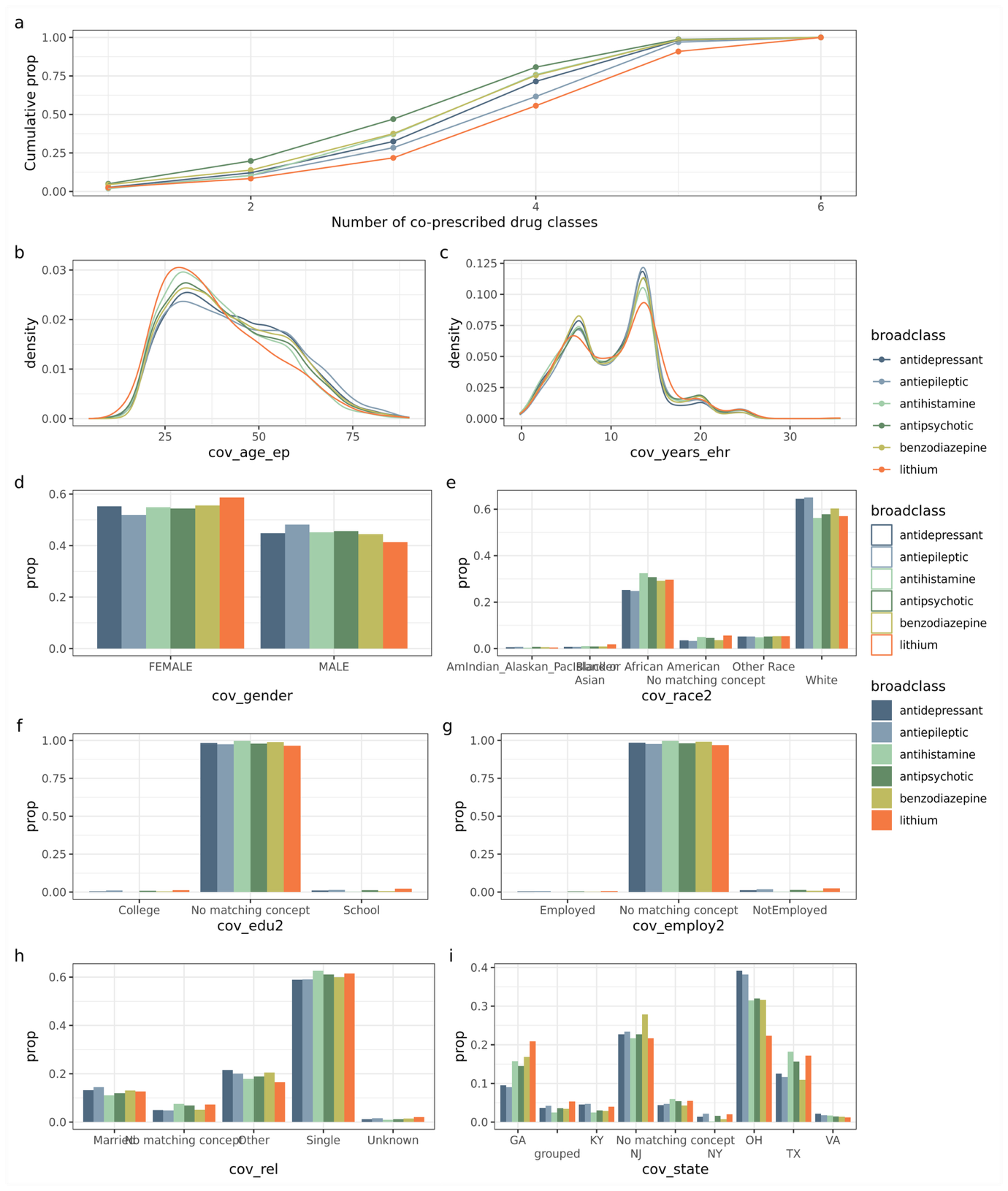

**Figure S20**: Relationships between prescribing and demographic factors and prescription of major psychotropic medication classes for hospital-associated manic/mixed episodes. a) Per drug-class cumulative proportion of manic/mixed episodes with increasing degrees of polypharmacy. Density plots of b) age at episode and c) years in EHR. Bar plots of categories for d) gender, e) race, f) educational attainment, g) employment status, h) relationship status, i) state.

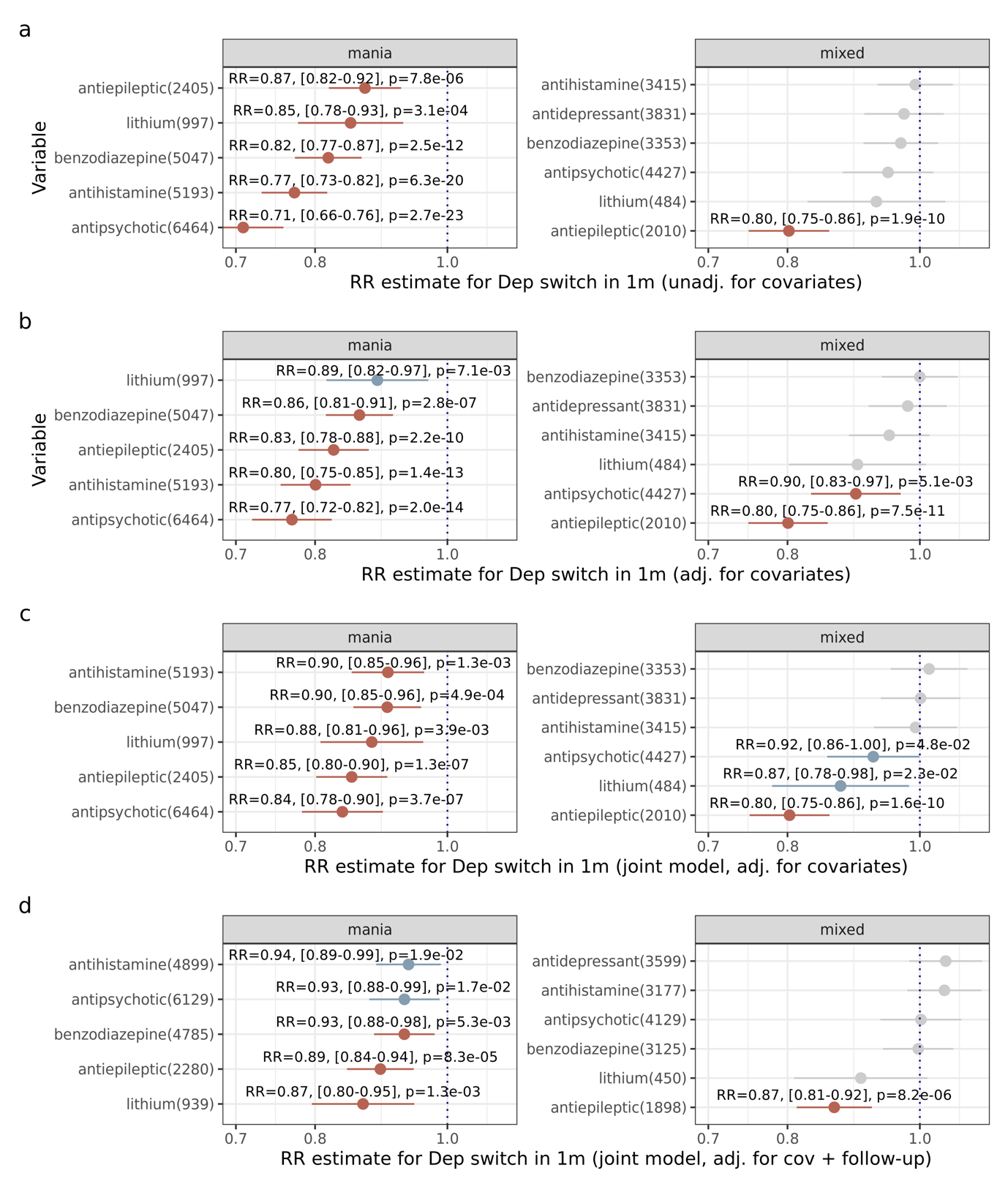

**Figure S21**: Rate ratio (±95% CI) for medication classes prescribed during hospital-associated manic/mixed episodes and their relationship with short mania/mixed-to-depression transition (≤1 month; modified Poisson regression with within-individual clustered standard errors). Results shown (a) without covariates, (b) including covariates, (c) jointly including all displayed medication in the same model including covariates, (d) sensitivity analysis adding to model (c) by adjusting for the mediator time to follow-up after hospital visit. Numbers on y-axis legend indicate sample size per drug class. Covariates are age_episode_ + age_EHR_ + years_inEHR_ + gender + education level + relationship status + race + state + hospital length of stay. Red indicates Bonferroni-significant effects; blue indicates nominally significant effects (p<0.05).

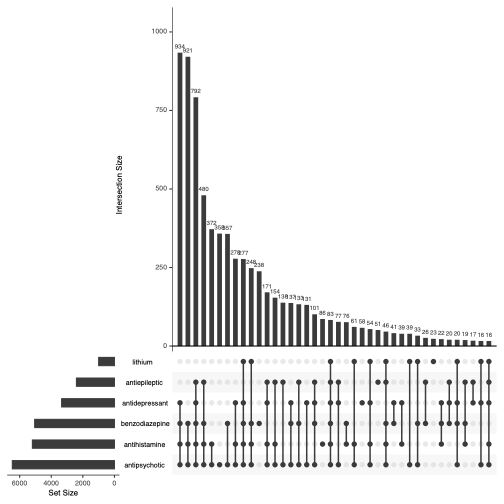

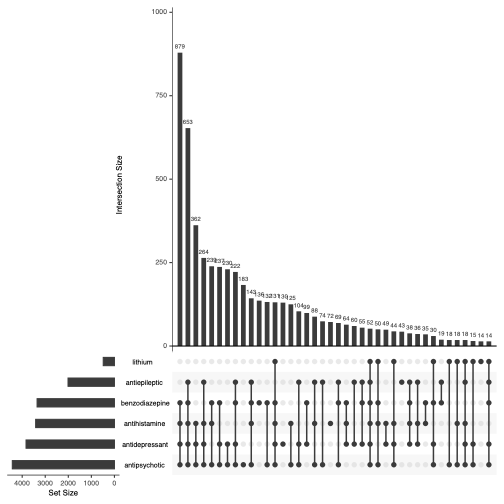

**Figure S22**: Upset plot for medications sets represented in the medication analysis, for manic (upper) and mixed (lower) episodes associated with hospitalisation.

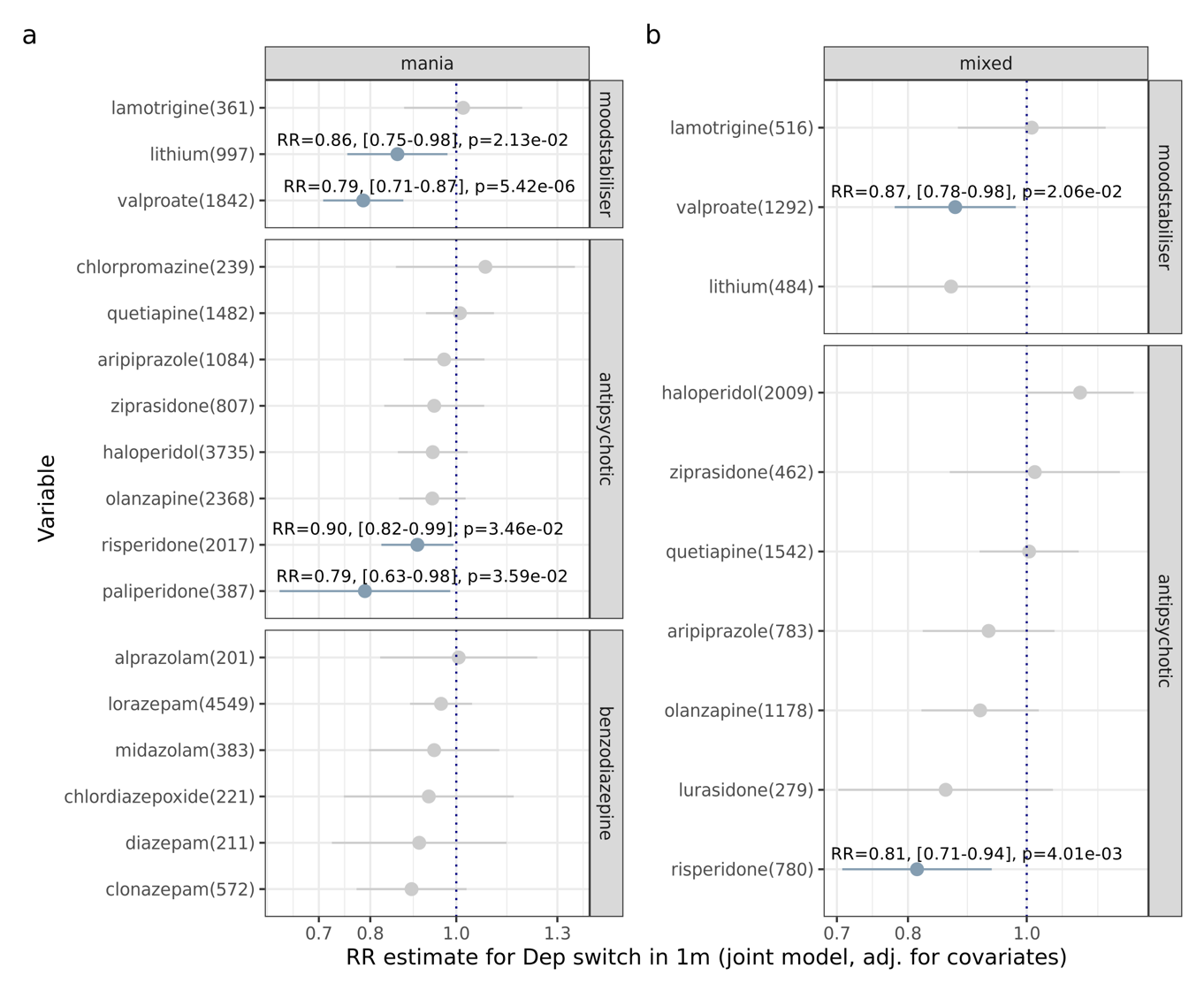

**Figure S23**: Joint analysis of individual medications belonging to the medication classes significantly associated with mania/mixed-to-depression transition time, restricting to manic/mixed episodes associated with hospitalisation, and with within-person clustered standard errors. Risk ratio (±95%CI) from modified Poisson regression. Adjusted for demographic covariates, hospital length of stay, time to follow-up and other major medication classes (antipsychotics for mixed episodes, antidepressants for manic episodes, benzodiazepines, antihistamines). Numbers on y-axis label indicate sample size per drug. Blue indicates p<0.05 (no multiple testing included as this is a sensitivity analysis). Medications had to be prescribed in >200 episodes to be included.

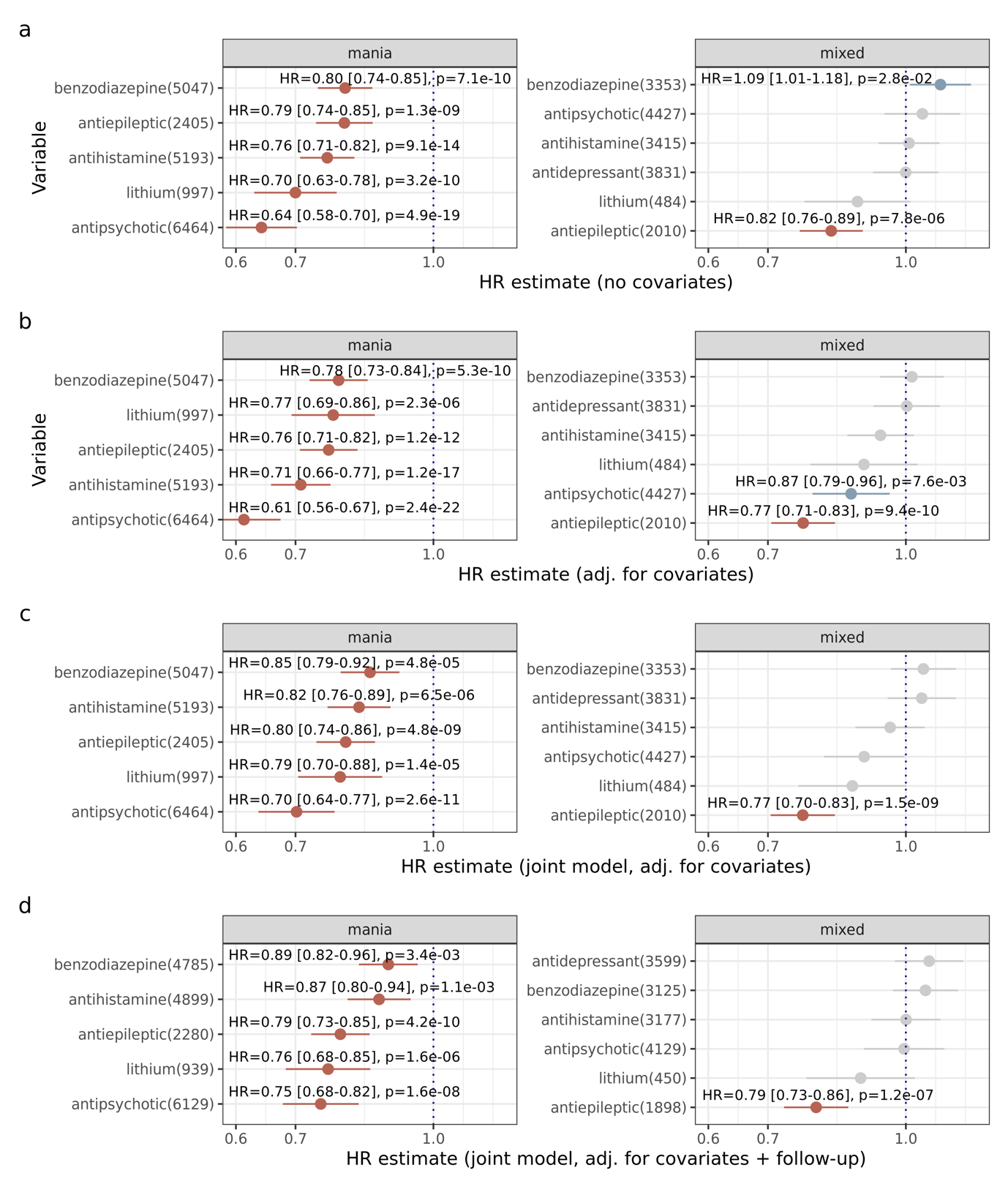

**Figure S24**: Cause-specific hazard ratio (±95% CI) from Cox regression with repeated risk intervals in medication class analyses restricting to manic/mixed episodes associated with hospitalisation, and with within-person clustered standard errors. Results shown (a) without covariates, (b) including covariates, (c) jointly including all displayed medication in the same model including covariates, (d) sensitivity analysis adding to model (c) by adjusting for the mediator time to follow-up after hospital visit. Numbers on y-axis legend indicate sample size per drug class. Model with covariates: mania/mixed-to-depression transition time ~ medications_0/1 during manic/mixed_ + age_episode_ + age_EHR_ + years_inEHR_ + gender + education level + relationship status + race + state + hospital length of stay. Red indicates Bonferroni-significant effects; blue indicates nominal significance (p<0.05).

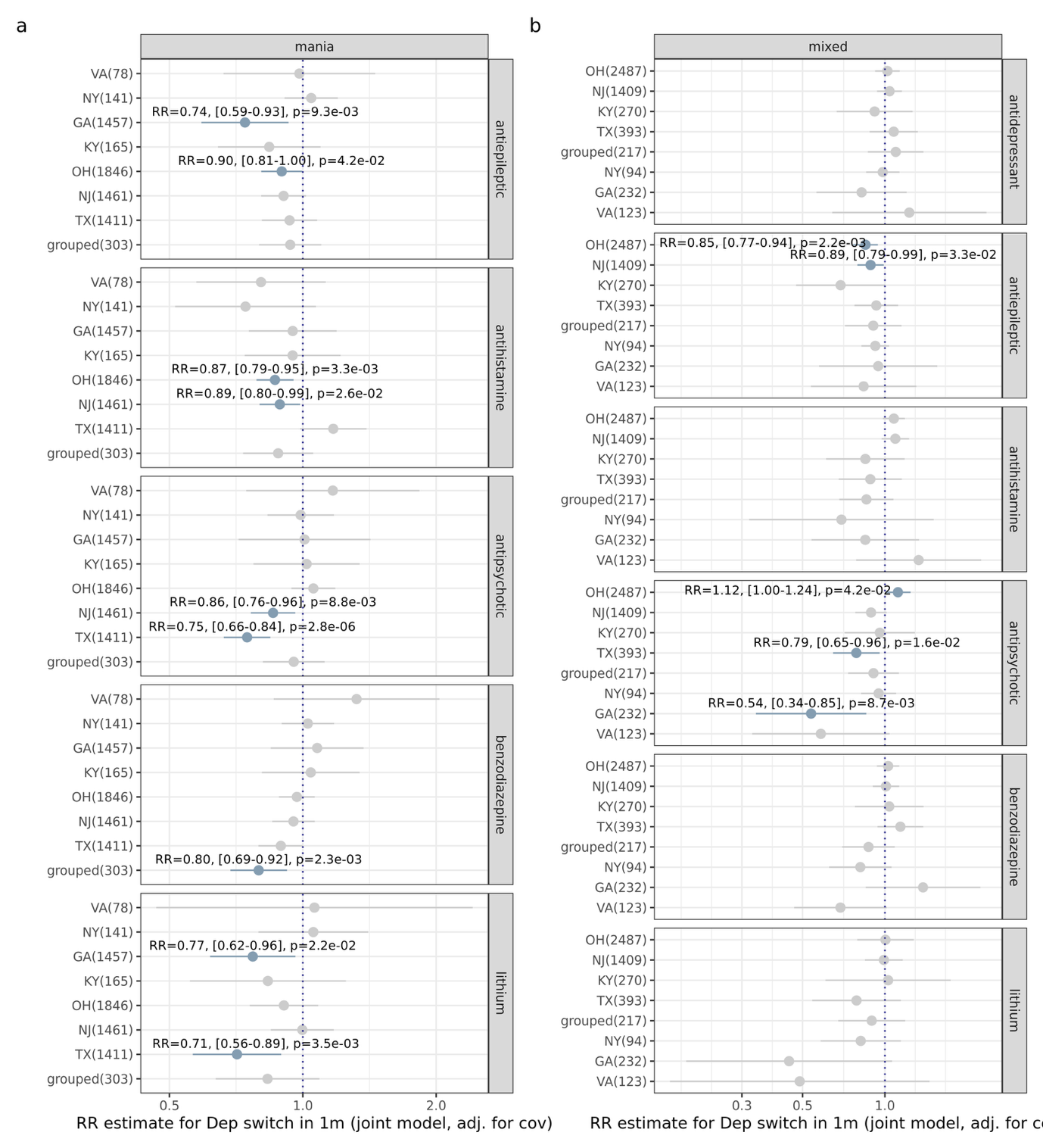

**Figure S25**: Medication sensitivity analysis stratifying by state of residence. Rate ratio (±95% CI) for medication classes prescribed during hospital-associated manic/mixed episodes and their relationship with short mania/mixed-to-depression transition (≤1 month; modified Poisson regression with within-individual clustered standard errors). Analyses here jointly include all listed medication classes and covariates: age_episode_ + age_EHR_ + years_inEHR_ + gender + hospital length of stay + time to next follow-up. Blue indicates p<0.05. Y-axis labels include number of episodes contributed by each state.

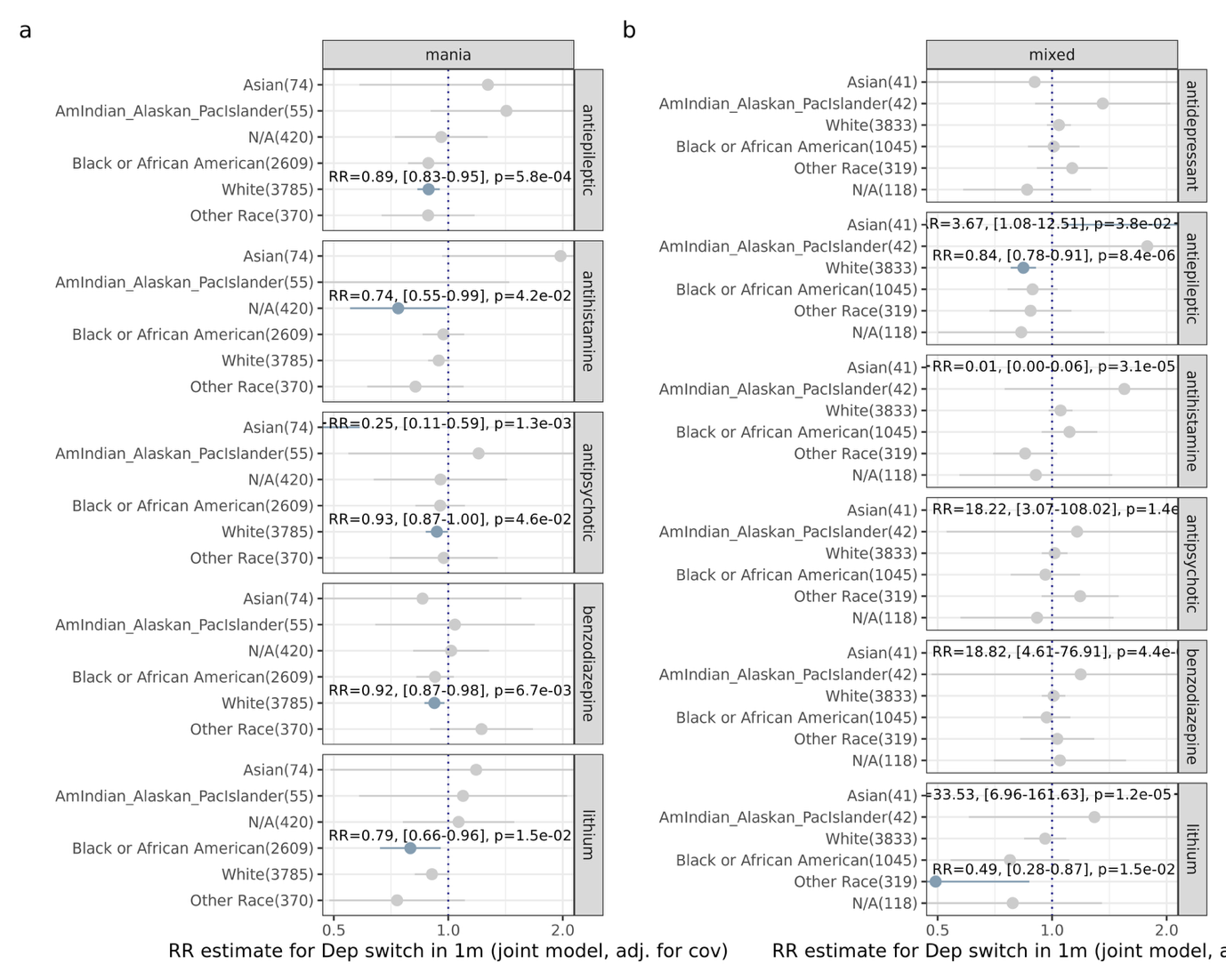

**Figure S26**: Medication sensitivity analysis stratifying by recorded racial group. Rate ratio (±95% CI) for medication classes prescribed during hospital-associated manic/mixed episodes and their relationship with short mania/mixed-to-depression transition (≤1 month; modified Poisson regression with within-individual clustered standard errors). Analyses here jointly include all listed medication classes and covariates: age_episode_ + age_EHR_ + years_inEHR_ + gender + state + hospital length of stay + time to next follow-up. Blue indicates p<0.05. Y-axis labels include number of episodes contributed by each racial group; each group had to contribute >20 episodes to be included in analysis for model stability.

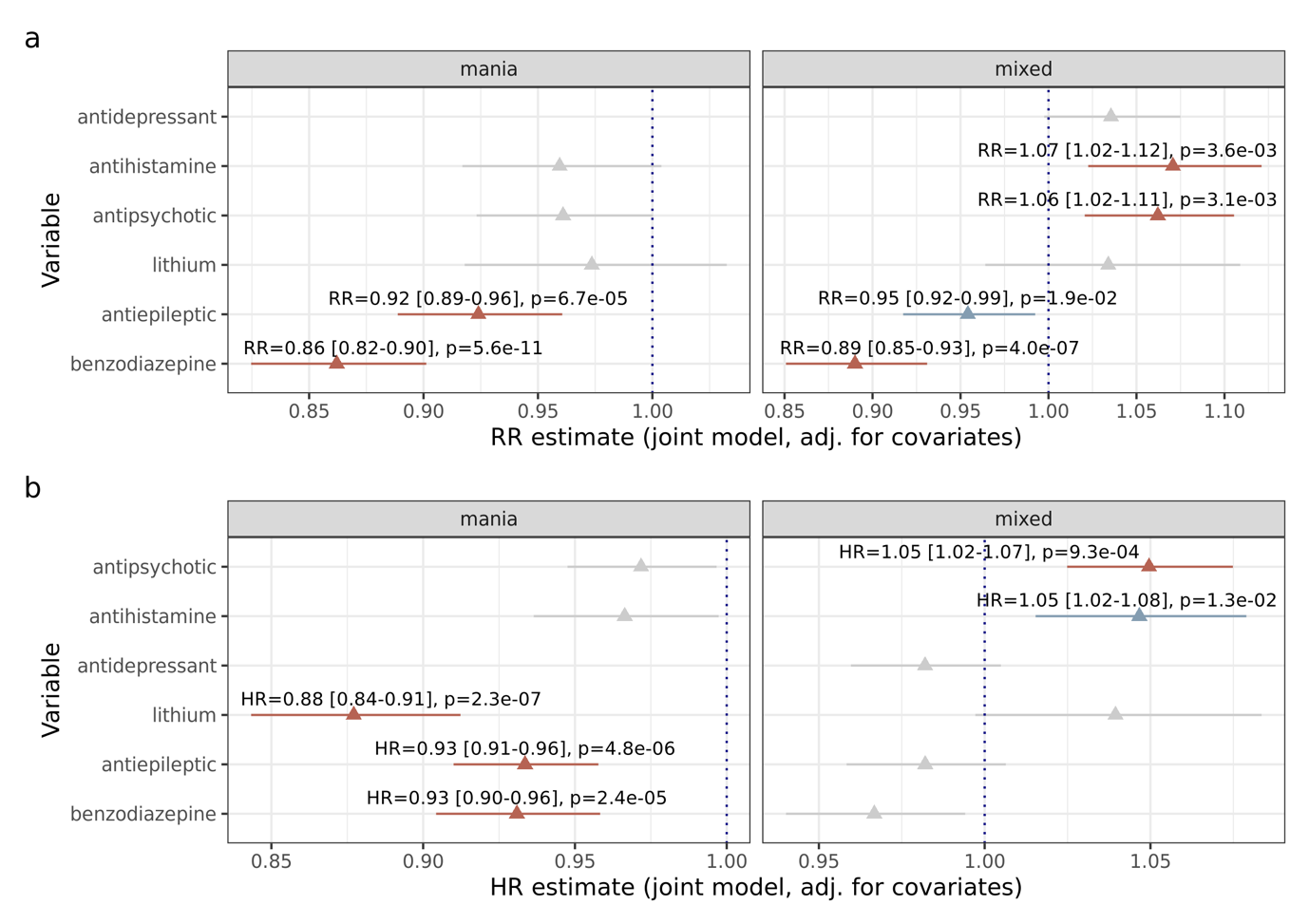
**Figure S27**: Sensitivity analysis for medications classes prescribed during any manic/mixed episode (i.e., no longer restricting to hospitalisation) and their relationship with (a) short mania/mixed-to-depression transition ≤1 month using modified Poisson regression (65,013 episodes from 36,406 individuals, filtering for states with ≥1% hospitalisation rate) and (b) time to depression using Cox regression with repeated risk intervals (, using Cox regression stratified by hospitalisation). All displayed medications modelled jointly with covariates. Medication classes are jointly modelled to account for co-prescription. Covariates: age_episode_ + age_EHR_ + years_inEHR_ + gender + education level + relationship status + race + state. Red indicates Bonferroni-significant effects; blue indicates nominally significant effects (p<0.05).

**a) Unadjusted DAG**

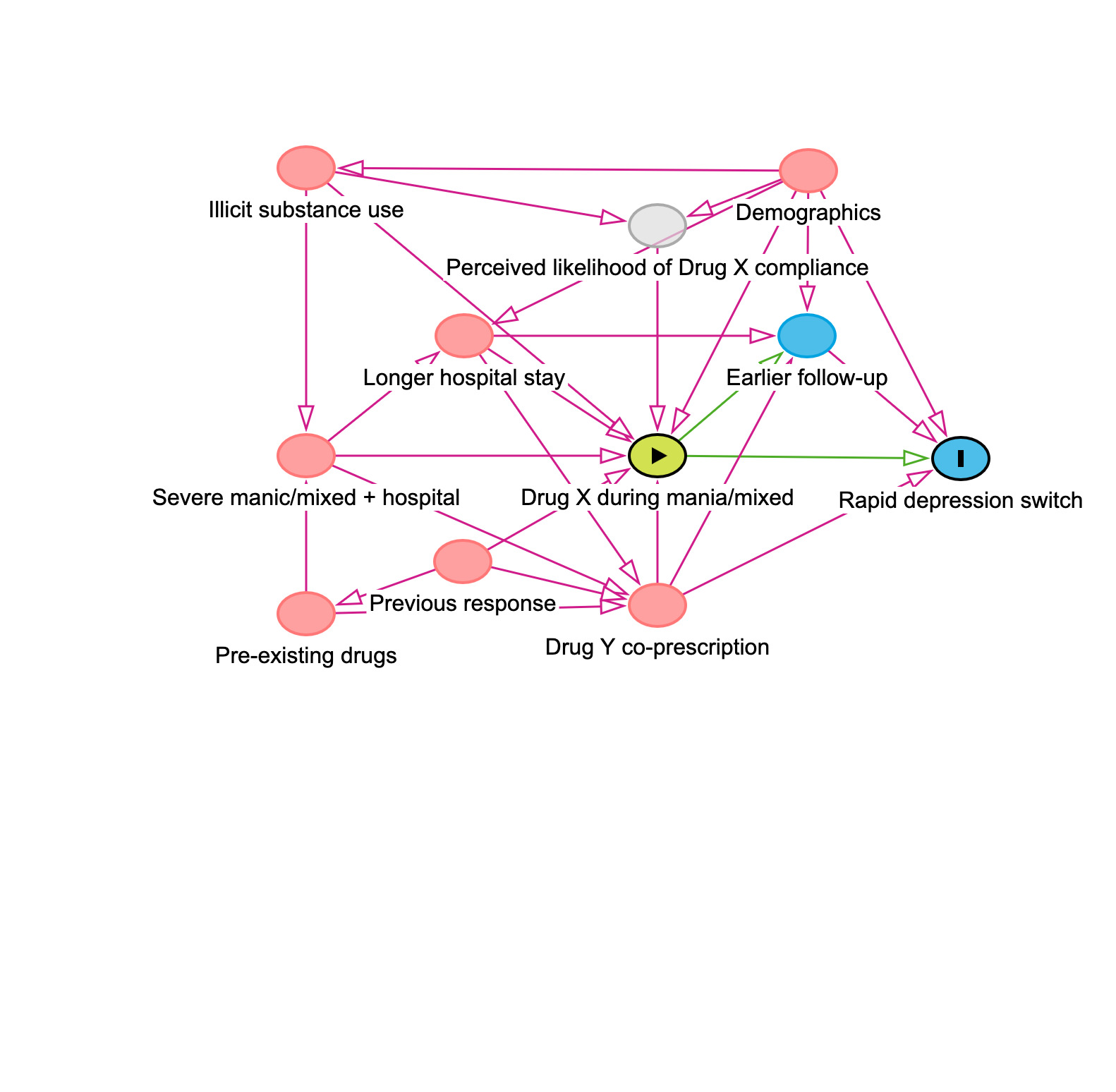

**b) Adjusted DAG**

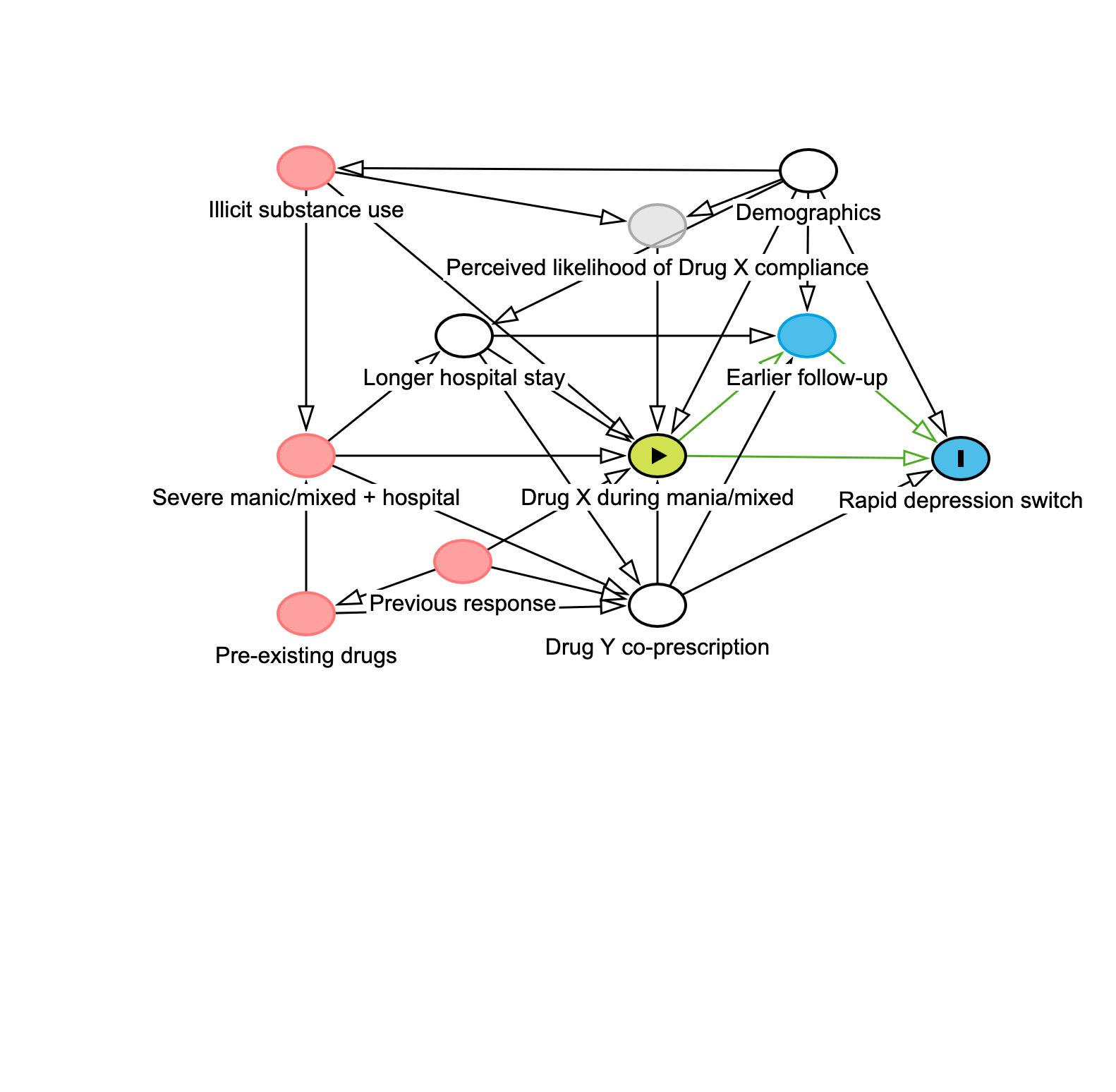

**Figure S28**: Directed acyclic graphs (DAGs) for factors influencing the relationship between prescription during manic/mixed episodes (exposure) and depression transition time (outcome). a) DAG without adjusting for covariates. b) DAG adjusting for covariates of longer hospital stay, drugY co-prescription and demographics. Pink arrows denote open biasing paths (i.e., unadjusted confounding). Green arrows denote open causal paths. Black arrows denote blocked or non-causal paths. Green oval indicates the exposure, blue oval with “I” indicates outcome, empty blue oval indicates mediator. Pink ovals indicate potential confounders. Grey oval indicates unmeasured confounder. White oval indicates confounder that has been adjusted for. Upper: unadjusted model. Lower: model adjusted for hospital length of stay, drug co-prescription and demographic variables.

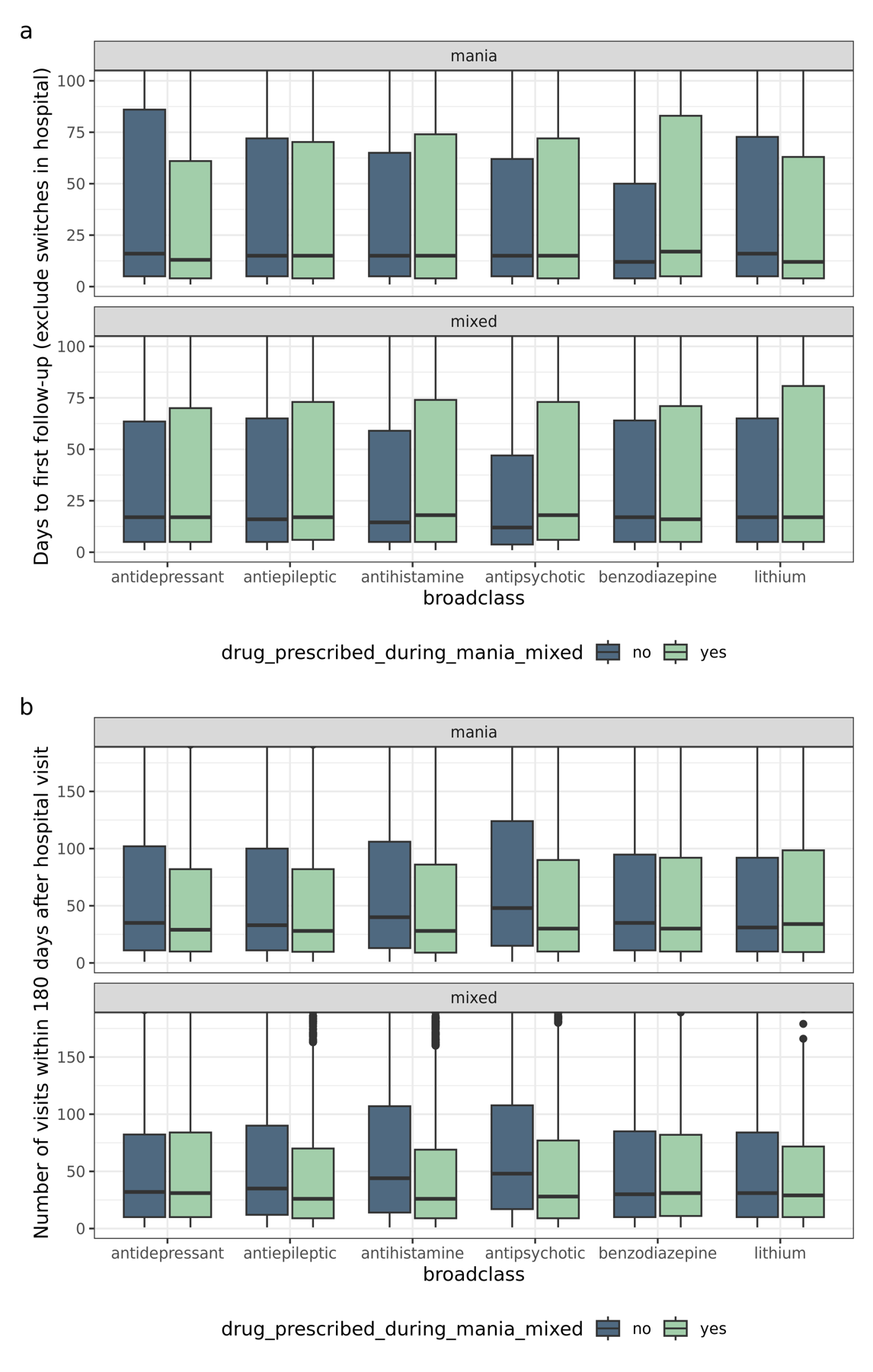

**Figure S29**: Boxplot for the distribution of (a) days from hospital visit end to the next visit and (b) total number of visits within 180 days from hospital visit end. Stratified by the presence or absence of prescription of each broad drug class within the first 7 days of the hospital-associated manic/mixed episode. Note that all the “no” group includes every episode where the drug is not prescribed. For panel a), episodes that switch during the hospital visit (and so were assigned 0 days to follow-up) were excluded.

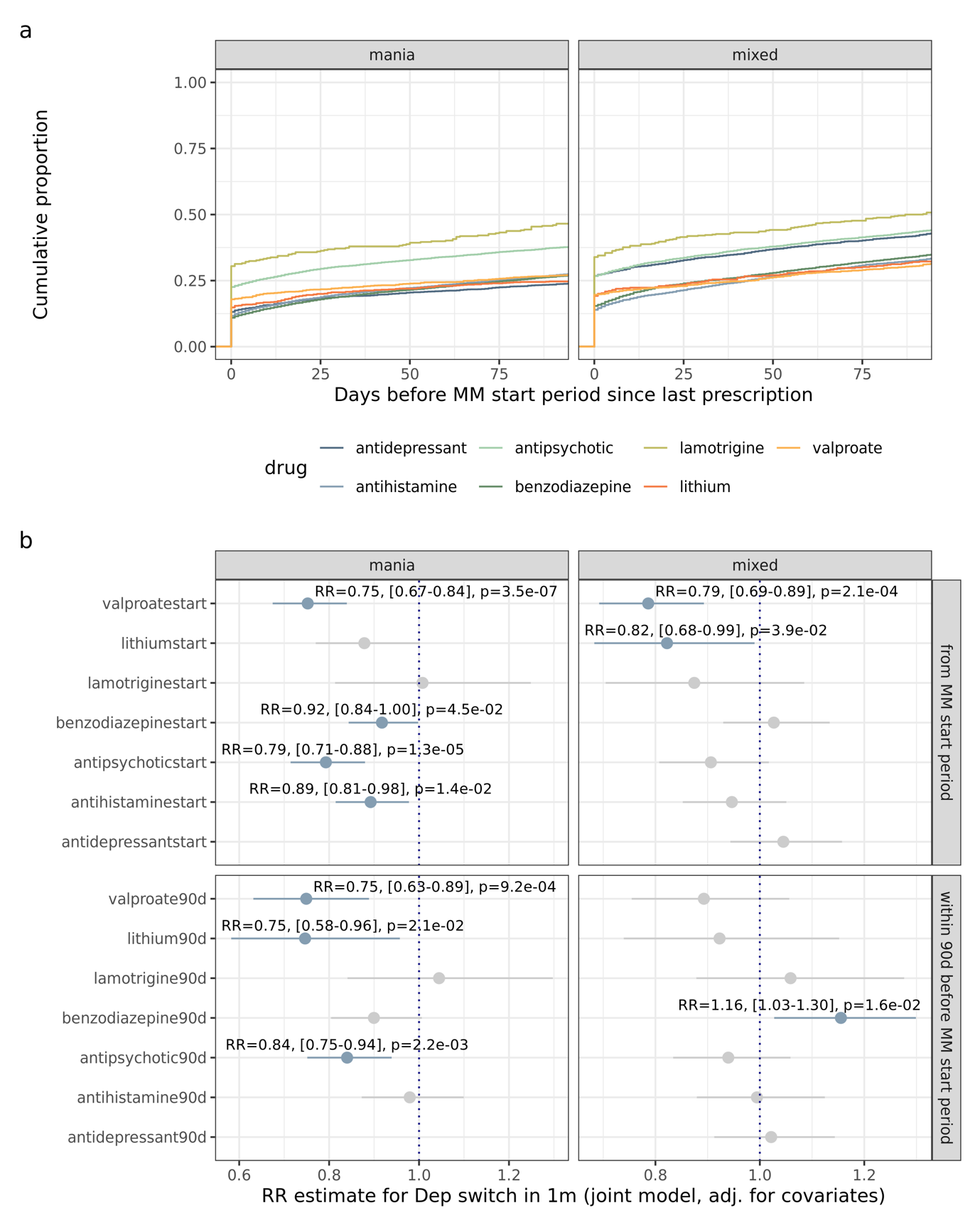

**Figure S30**: Sensitivity analysis exploring recency of last prescription on the medication results. Drugs aggregated at the class level except for mood stabilisers due to delays in effect related to titration. a) Cumulative proportion the per drug/class for the distribution of days since the last prescription. Plot limits are restricted to the 90 days before the manic/mixed start period (defined as 7 days before the first manic/mixed diagnosis code, for consistency with the medication analysis). b) Medications prescribed during hospital-associated manic/mixed episodes and their relationship with short mania/mixed-to-depression transition (≤1 month; modified Poisson regression). Results are stratified by whether there are records of a prescription within the preceding 90 days, in both cases comparing to the group which did not receive the relevant medication. All displayed medications modelled jointly with covariates. Blue indicates p<0.05 (no multiple testing was employed in this sensitivity analysis).

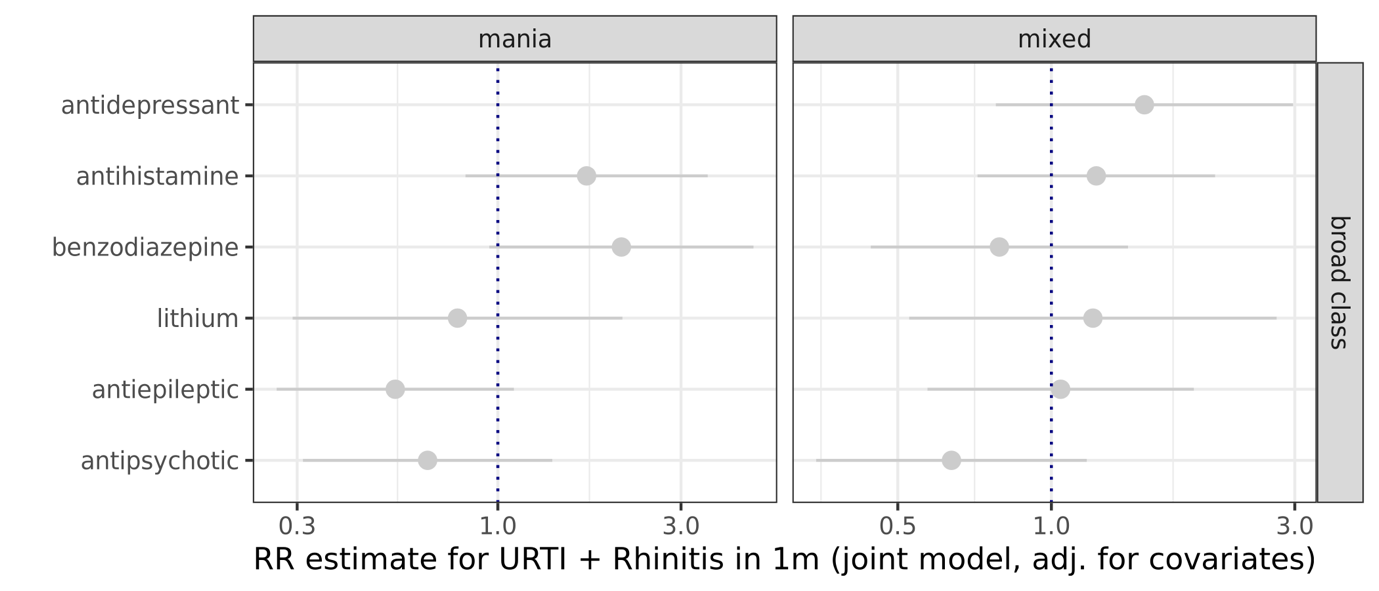

**Figure S31**: Medications classes prescribed during hospital-associated manic/mixed episodes and their relationship with a composite negative control outcomes of upper respiratory tract infections (URTIs) and allergic rhinitis within 1 month of mania end (modified Poisson regression). All displayed medications modelled jointly with covariates.

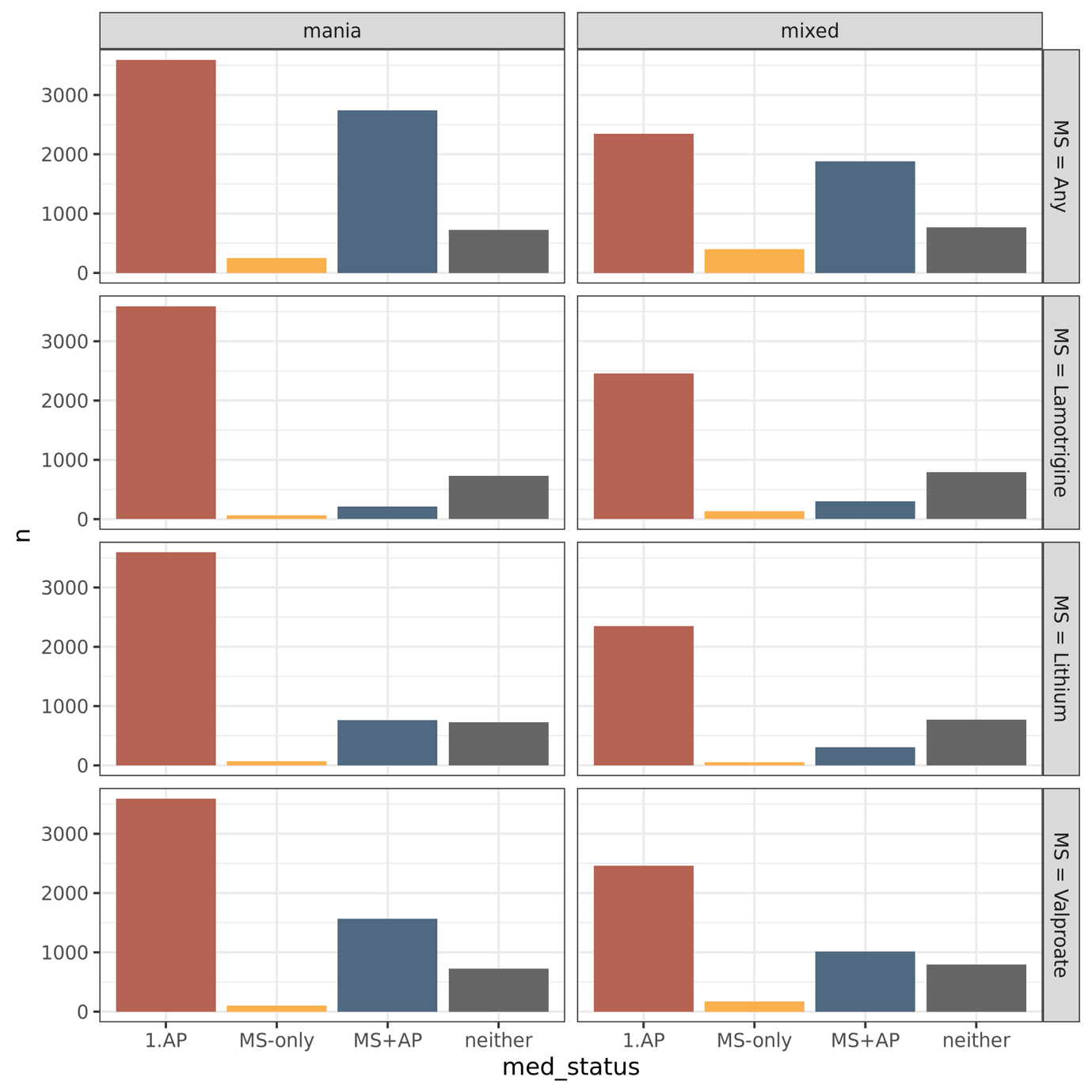

**Figure S32:** Barplot of the number of individuals in each category for the mood stabiliser/antipsychotic combination analysis in **Figure 4**. Individuals with a prescription of lamotrigine, lithium or valproate are not included in analyses of the other mood stabilisers (except for the “any” mood stabiliser analysis).

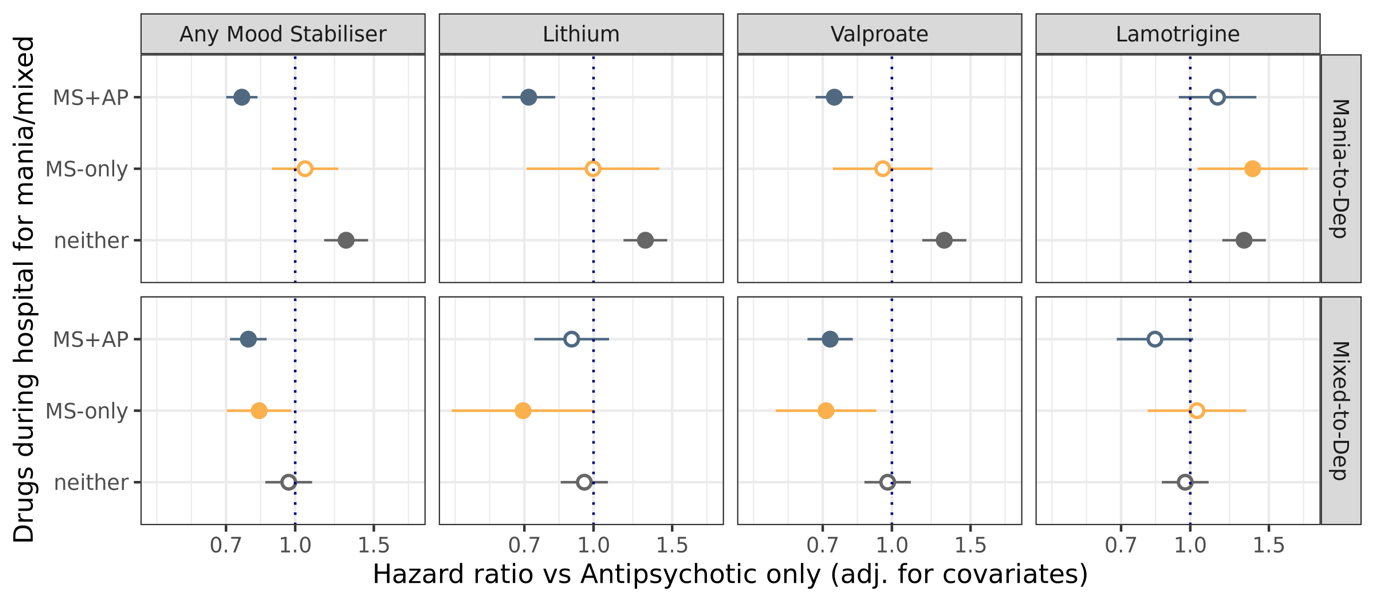

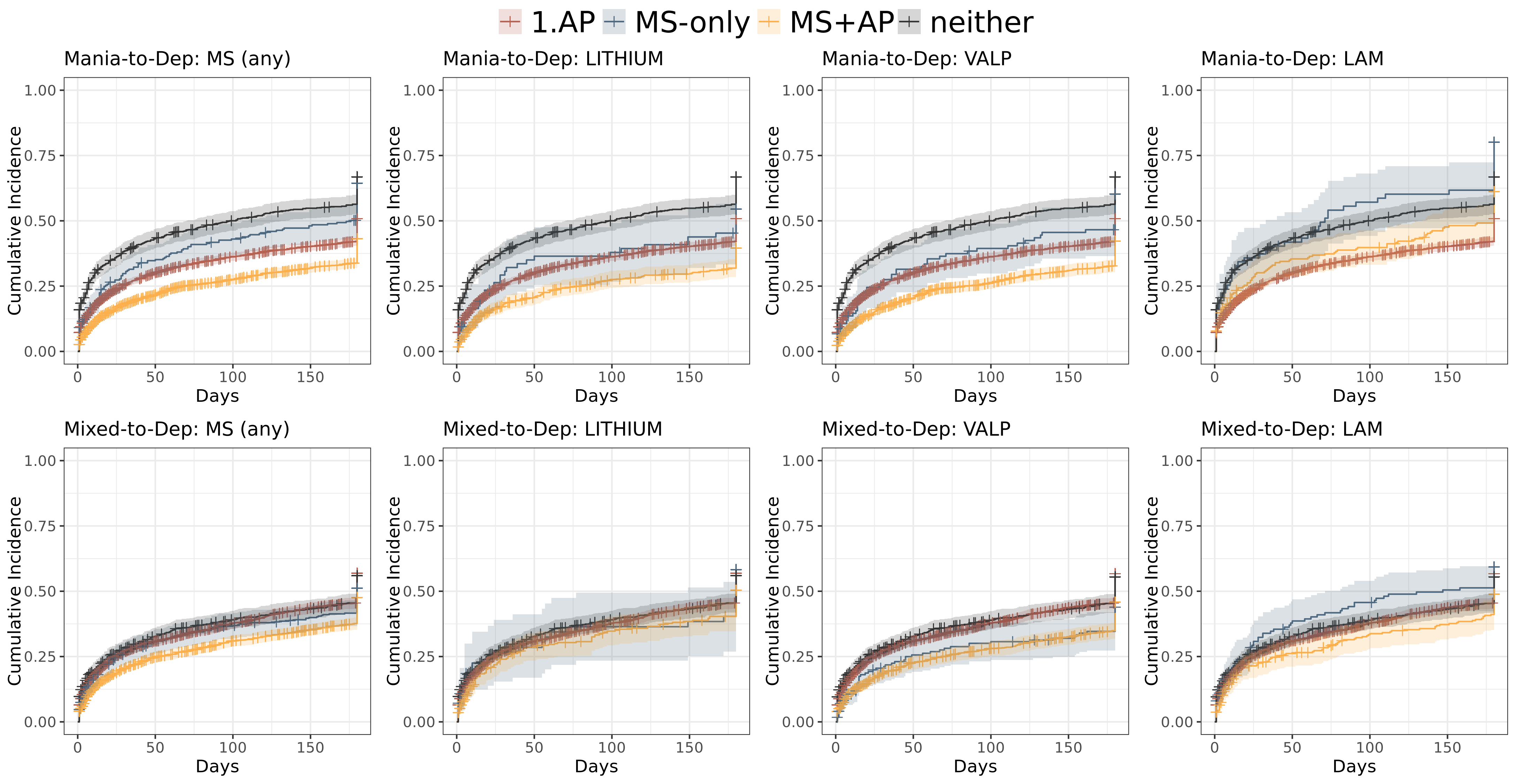

**Figure S33:** Mood stabiliser and antipsychotic combination effects on mania/mixed-to-depression transition. (upper) Cause-specific hazard analysis. Prescriptions had to occur within 7 days of manic/mixed episode start. Mood stabilisers are here considered aggregated as a class, then individually (each of lithium, valproate and lamotrigine). Filled points indicate p<0.05. “MS”: mood stabiliser; “AP”: antipsychotic; “MS+AP”: combination therapy. Source data: **Table S10**. (lower) Cumulative incidence plots per group.

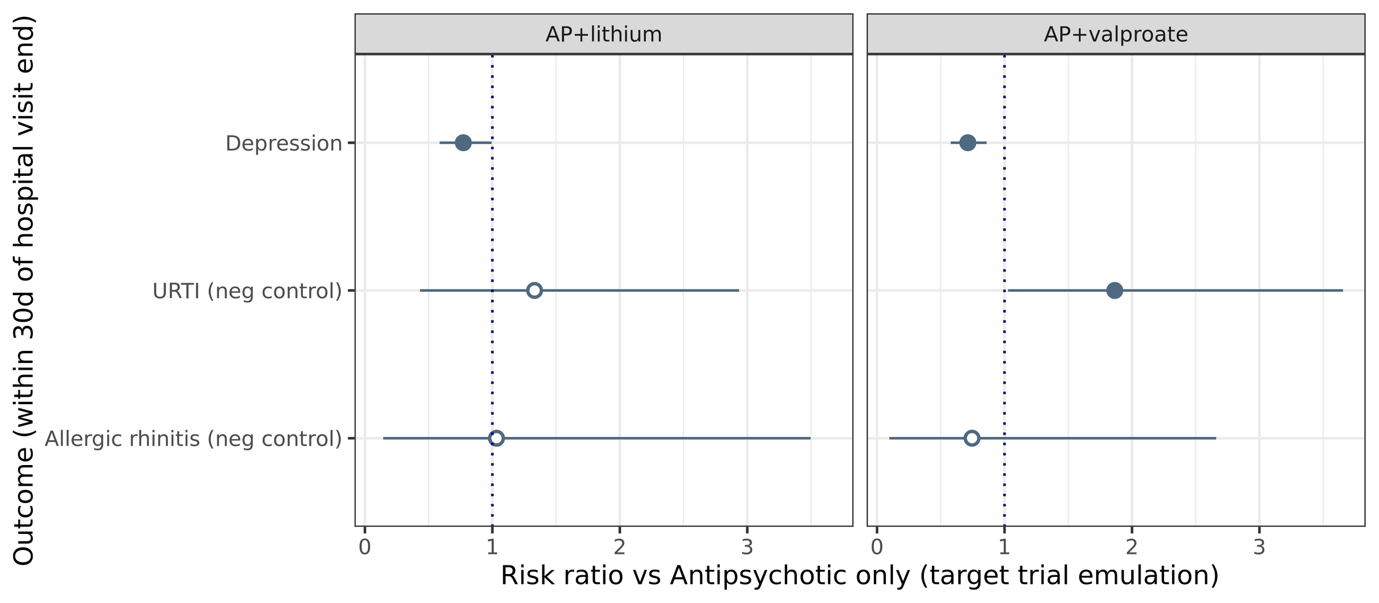

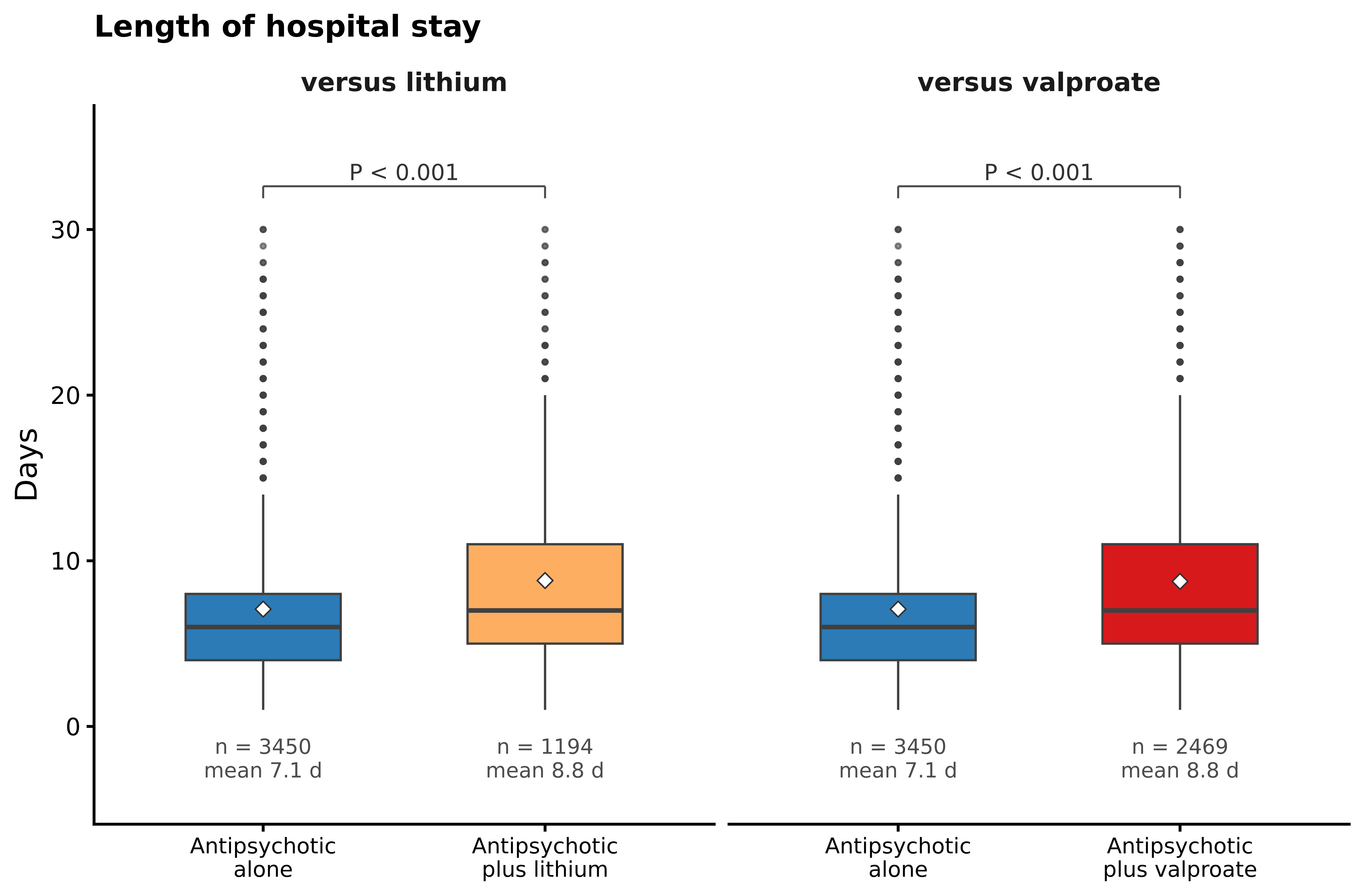

**Figure S34**: Target trial emulation results. (upper) Relative risks from target trial emulation analysis comparing lithium+antipsychotic and valproate+antipsychotic to antipsychotic-alone for depression (primary outcome) and two negative control outcomes within 30 days of hospital visit end. Filled points denote p<0.05. AP: antipsychotic; URTI: upper respiratory tract infection. (lower) Boxplots comparing length of hospital stay between the exposure arms.

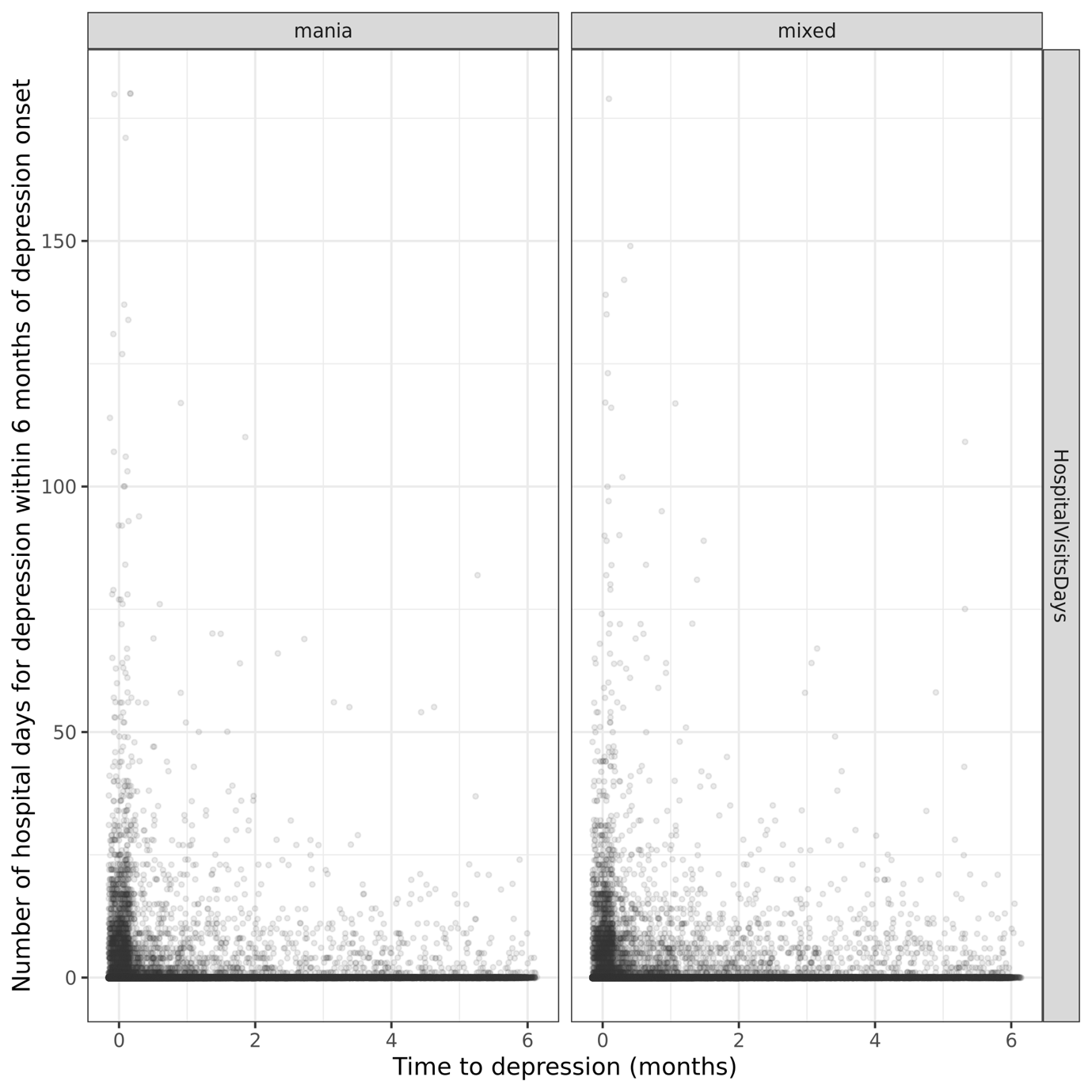

**Figure S35**: Scatterplot of depression-related hospital days and mania/mixed-to-depression transition time. Data from 101,840 episodes where depression has onset ≤6 months from the end of the manic/mixed episode.

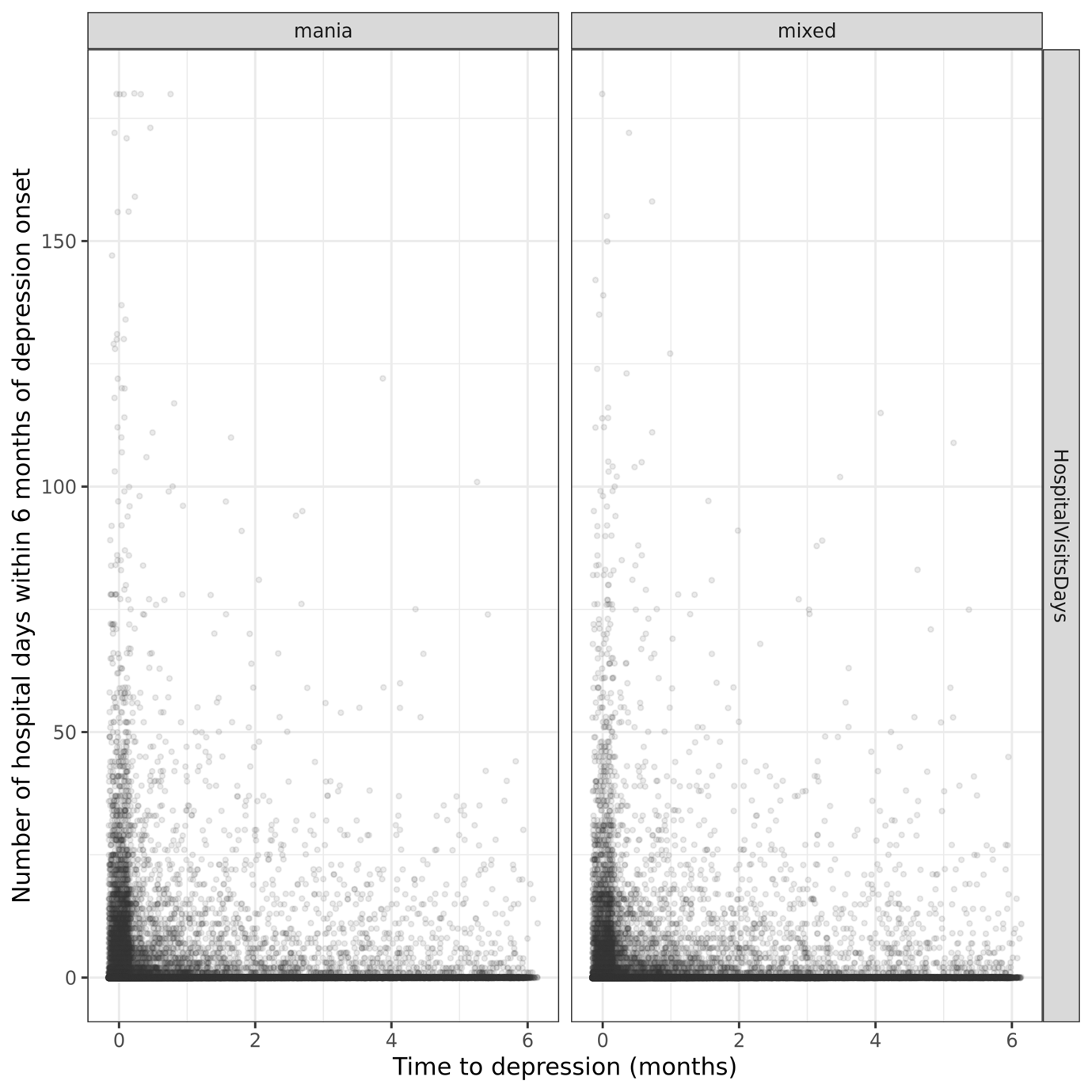

**Figure S36**: Scatterplot of all-cause hospital days and mania/mixed-to-depression transition time. Data from 101,840 episodes where depression has onset ≤6 months from the end of the manic/mixed episode.

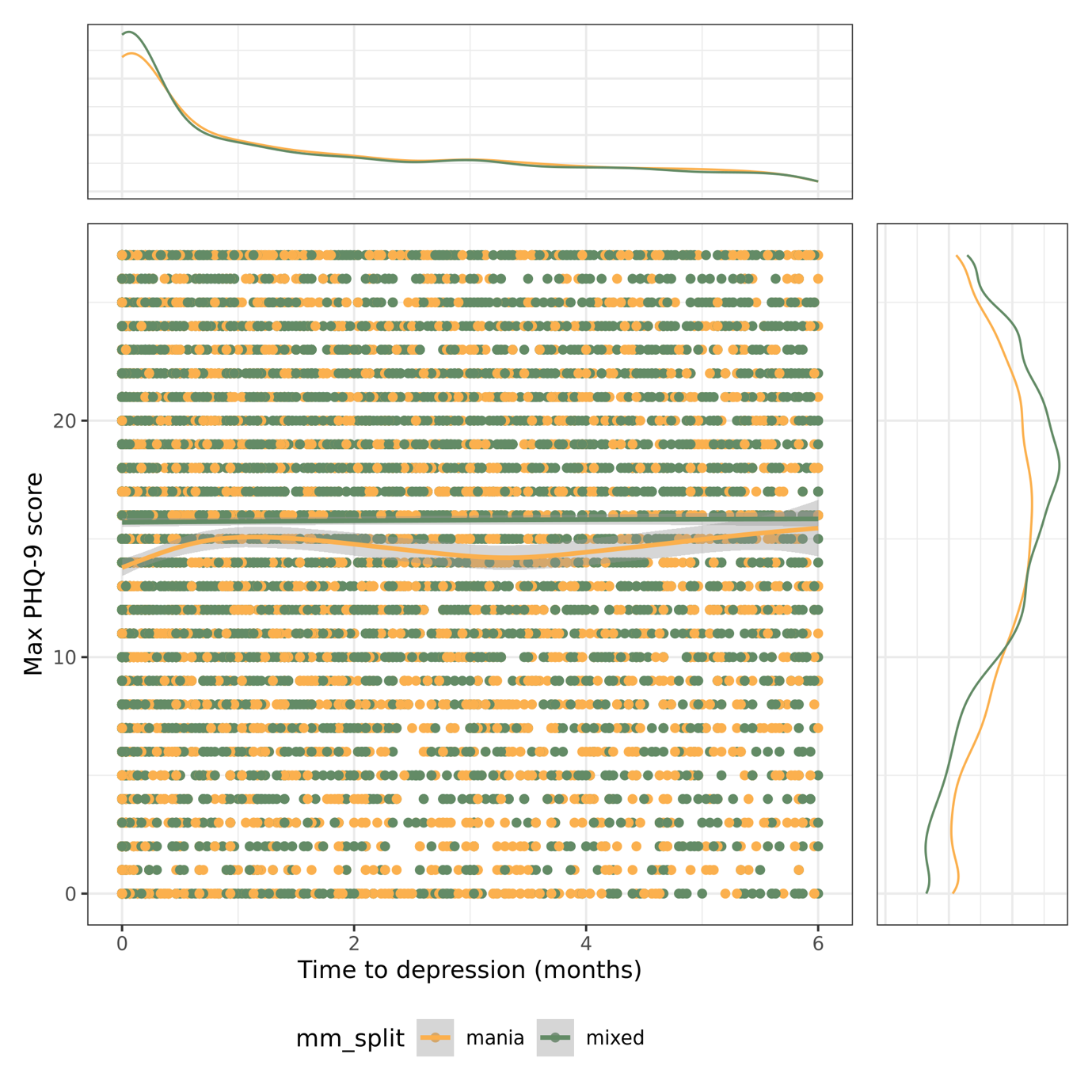

**Figure S37**: Scatterplot of time to depression and maximum PHQ-9 score. Episodes only included if there were >=2 PHQ-9 scores of which at least one occurred within 6 months of the end of the manic/mixed episode, and the others could occur up to 12 months following a manic/mixed episode. Results from 28,448 episodes.

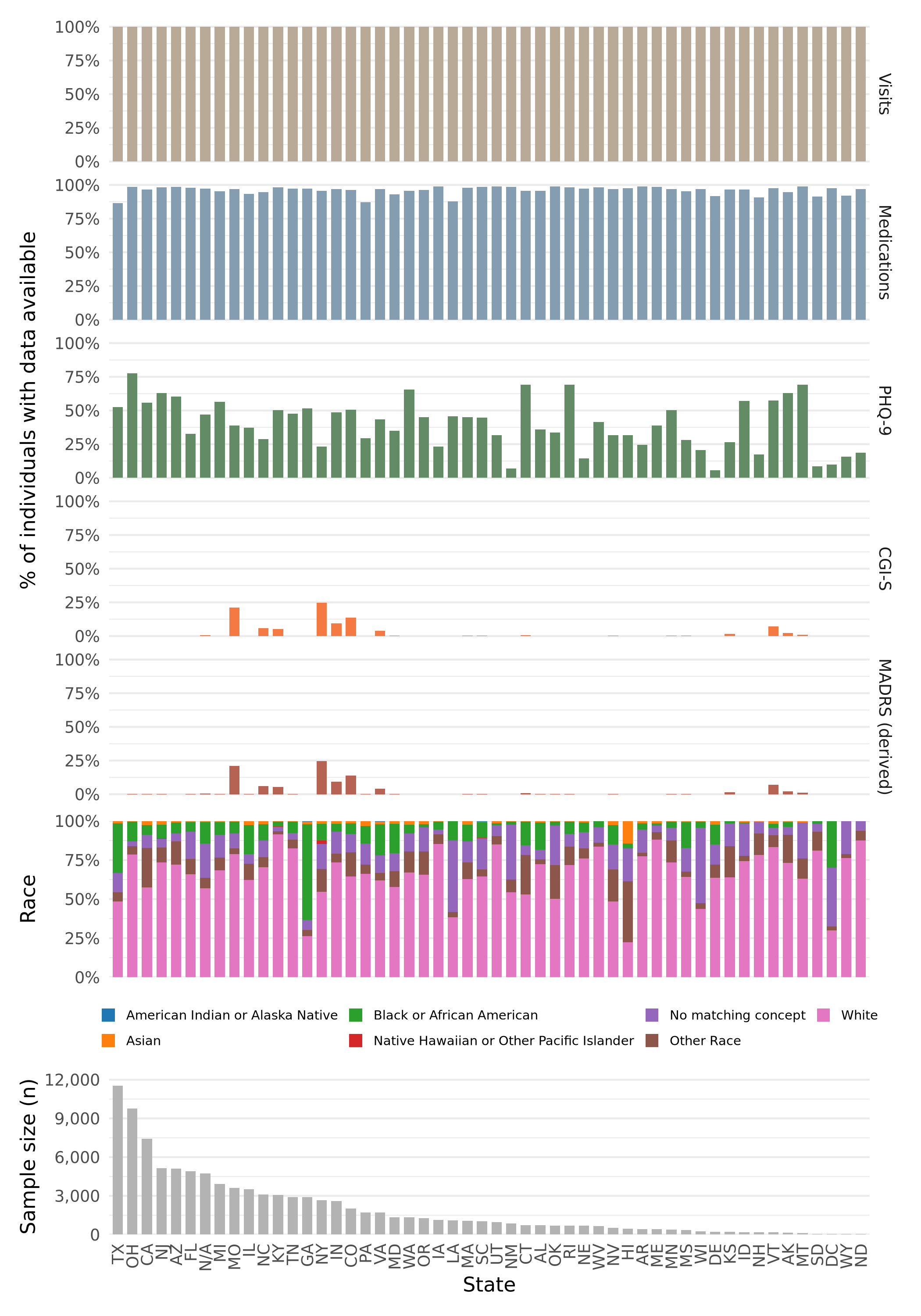

**Figure S38**: Percentage of individuals in this bipolar sample with medication and clinical scale data availability on a per-state basis.
